# Predicting the course of Covid-19 and other epidemic and endemic disease

**DOI:** 10.1101/2021.12.26.21268419

**Authors:** Ana Cascon, William F. Shadwick

## Abstract

The Gompertz Function is an accurate model for epidemics from Cholera in 1853 to Spanish Flu in 1918 and Ebola in 2014. It also describes the acute phase of annual outbreaks of endemic influenza and in all of these instances it has significant predictive power.

For Covid-19, we show that the Gompertz Function provides accurate forecasts not just for cases and deaths but, independently, for hospitalisations, intensive care admissions and other medical requirements. In particular Gompertz Function projections of healthcare requirements have been reliable enough to allow planning for: hospital admissions,intensive care admissions,ventilator usage, peak loads and duration.

Analysis of data from the ‘Spanish Flu’ pandemic and the endemic influenza cycle reveals alternating periods of Gompertz Function growth and linear growth in cumulative cases or deaths. Linear growth means the Reproduction Number is equal to 1 which in turn indicates endemicity.

The same pattern has been observed with Covid-19. All the initial outbreaks ended in linear growth. Each new outbreak has been preceded by a period of linear growth and has ended with a transition from Gompertz Function growth to linear growth. This suggests that each of these outbreak cycles ended with a transition to endemicity for the current dominant strain and that the normal seasonal respiratory virus periods will continue to see new outbreaks. It remains to be seen if widespread vaccination will disrupt this cyclicality.

Because both Gompertz Function Growth and linear growth are accurately predictable, the forecasting problem is reduced to identifying the transition between these modes and to improving the performance in the early Gompertz Function growth phase where its predictive power is lowest.

The dynamics of the Gompertz Function are determined by the Gumbel probability distribution. This is an exceptional distribution with respect to the geometry determined by the affine group on the line which is the key to the role of the Gumbel distribution as an Extreme Value Theory attractor. We show that this, together with the empirically observed asymmetry in epidemic data, makes the Gompertz Function growth essentially inevitable in epidemic models which agree with observations.

## 1 Overview and Outline

### 1.1 Overview

Galileo showed by experimental observations that, in the absence of friction, projectiles follow a parabolic trajectory. Only some decades later did Newton’s laws of motion explain *why* that was the case.

In the interim, the parabolic trajectory was what would now be called a ‘phenomenological model’–a mathematical formulation of a process that describes it without explaining why, or by what mechanism, it takes place. If we can make reasonable observations of points on the projectile’s trajectory, we can approximate the parameters that determine the parabola. The model can then make accurate predictions about the future on the basis of our observations of the past.

We show that major epidemics in the past from Cholera in 1853 to Spanish Flu in 1918 and Ebola in 2014 are all described very well by a 3-parameter phenomenological model–the Gompertz Function.

So it is not surprising that the Covid-19 epidemic also followed the Gompertz Function Model.

We demonstrate through numerous examples that this model has significant short to medium term predictive power. The error between the predictions made by the Gompertz Function fit at a given time and the subsequent out of sample data remains small, first only for a few days and then for progressively longer and longer periods.

Most importantly, this means that the model can be used to make increasingly accurate forecasts of healthcare requirements as a Covid-19 outbreak (or other epidemic) progresses. Such forecasts are essential to address the fear that hospitals will be overwhelmed–which has been a major preoccupation almost everywhere.

In every location for which we have obtained data, the predictions of the Gompertz Function Model could have been used in planning for: hospital admissions and intensive care admissions as well as such things as ventilator requirements, timing and size of peak loads and duration of an outbreak.

The Gompertz Function Model is based on observed data and the model parameters can be calculated very rapidly by a simple transparent process (non-linear regression) for which there are numerous ‘off the shelf’ implementations.

Fortunately, given how little we know about how coronaviruses are transmitted, [1] using the Gompertz Function Model to forecast hospital demand doesn’t require models of infections, cases or the proportion of cases that will require hospitalisation. It is based simply on counting hospital and intensive care admissions.

By analysing a multi-year record of daily influenza cases in Portugal, we have shown the annual influenza cycle there is described to very good accuracy by alternating periods of Gompertz Function growth and periods of *linear growth* (where the Reproduction Number must be approximately equal to 1).

These cycles of Gompertz Function and linear growth provide a natural way of separating epidemic waves (as in the ‘Spanish Flu’) and of observing the transition from epidemic to endemic phases of disease.

Alternating Gompertz Function growth and linear growth is exactly what has been observed in Covid-19. The initial outbreaks began with Gompertz Function growth and then switched to linear growth. Subsequent outbreaks in 2020 followed the normal seasonal pattern of influenza [9] and other common respiratory illnesses. As yet, none of the seasonal ‘slots’ have been missed out–so we should be alert for repetitions of this pattern. After large scale vaccination programs there have been some outbreaks in periods other than the normal seasonal ones. In each case they followed the same alternating Gompertz Function growth-linear growth regime.

This Extended Gompertz Function Model of alternating Gompertz Function and linear growth phases provides an important mechanism for the early observation of new outbreaks and the ability to accurately forecast resulting demands on the healthcare system: identify transition from a linear growth regime to Gompertz Function growth, then use the Gompertz Function Model to provide forecasts.^1^

For a Gompertz Function, the speed of infection peaks with 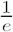 or approximately 37% of the susceptible population infected and declines steadily thereafter. This marked asymmetry with rapid growth and slow decay is observed to very good agreement in all of our data. It is also consistent with recent generalisations of compartmental models that remove the unrealistic assumption of a perfectly homogeneous susceptible population.

The dynamics of the Gompertz Function are determined by the Gumbel probability distribution.This is an exceptional distribution with respect to the geometry determined by the affine group on the line which is the key to the Gumbel distribution’s role as an Extreme Value Theory attractor. We show that this, together with the empirically observed asymmetry in epidemic data, makes Gompertz Function growth an essentially inevitable feature in epidemic models which agree with observations.

### 1.2 Outline of Contents by Section

In Section 2 we discuss the purpose and predictive power of models and review the performance of the Gompertz Function model in a wide variety of infectious disease outbreaks prior to 2020 and the Covid-19 epidemic.

Next we illustrate the Gompertz Function Model in the initial Covid-19 outbreaks. Our focus is on healthcare and the predictive power to make forecasts of practical use for planning.

In Section 3 we discuss the mathematical properties of the model. We show how these can be used to infer features of disease dynamics, including the reproduction number and herd immunity thresholds.

In Section 4 we use a multi-year data set of Influenza cases in Portugal as a guide to extend the epidemic model to include epidemic-endemic cycles where Gompertz Function growth alternates with linear growth. We also show that linear growth periods separated the waves of the ‘Spanish Flu’ in England.

Finally we demonstrate the same sequence of Gompertz Function Growth and Linear Growth in Covid-19 outbreaks.

In Section 5 we discuss a number of important aspects of the fits of Gompertz Functions to data.

Section 6 is a brief note on the relation between the Gompertz Function Model and compartmental models.

In Section 7 we discuss the use of the Gompertz Function Model in planning for new outbreaks of Covid-19 and as well as for seasonal Influenza and future pandemics.

Appendix A provides a brief account of the geometry behind the properties of the Gumbel probability distribution that underlies the Gompertz function and the way Gumbel distribution’s role as an Extreme Value Theory attractor helps to explain the ubiquity of Gompertz Functions in observations of epidemic data.

Appendix B contains links to our Covid-19 data sources.

## 2 Predicting the Course of An Epidemic

### 2.1 Mathematical Models

The purpose of mathematical models is to allow us to see into the future. To predict things such as the number of people who will be admitted to hospital with Covid-19 in the next two weeks or the trajectory of a Mars lander, for example.

The distance we can see and the accuracy of our vision may vary greatly, as it does in these two cases, but predicting the future is the fundamental goal of mathematical models of processes in the real world. In our case, we will focus on making short term predictions (a week to several weeks) with a ‘reasonable’ degree of accuracy (say within 10 −20%), which is a very different goal from the models which are used to create scenarios outlining a wide range of possible futures contingent on different events or policy decisions.^2^

Early in 2020, people feared that the Covid-19 epidemic was growing exponentially. This was a model that predicted a catastrophic future with explosive growth in the numbers of cases and deaths.

But Michael Levitt observed [12] that cumulative cases in the Covid-19 outbreak seemed instead to be growing according to a Gompertz Function. This is a simple 3-parameter phenomenological model for epidemic growth in which the number of cases at time *t* is:

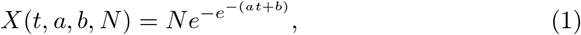

where *N, a* and *b* are parameters to be calculated^3^ from the reports of daily case numbers. By contrast with exponential growth where the doubling time is constant, in Gompertz Function growth *the time required for cases to double increases rapidly*, damping the epidemic more and more as it progresses.

And this is what was subsequently seen as the Covid-19 epidemic spread. A very asymmetric growth pattern with a rapid rise and a much slower fall off after the peak.

This is apparent in the Covid-19 deaths reported daily in Italy in the spring of 2020, shown in Figure 1. Because of the large degree of variability in the daily data and to illustrate the trend in the data we have included a rolling 7-Day average.^4^ This shows the high degree of asymmetry in the daily deaths which rise much more quickly than they fall.^5^

**Figure 1:**
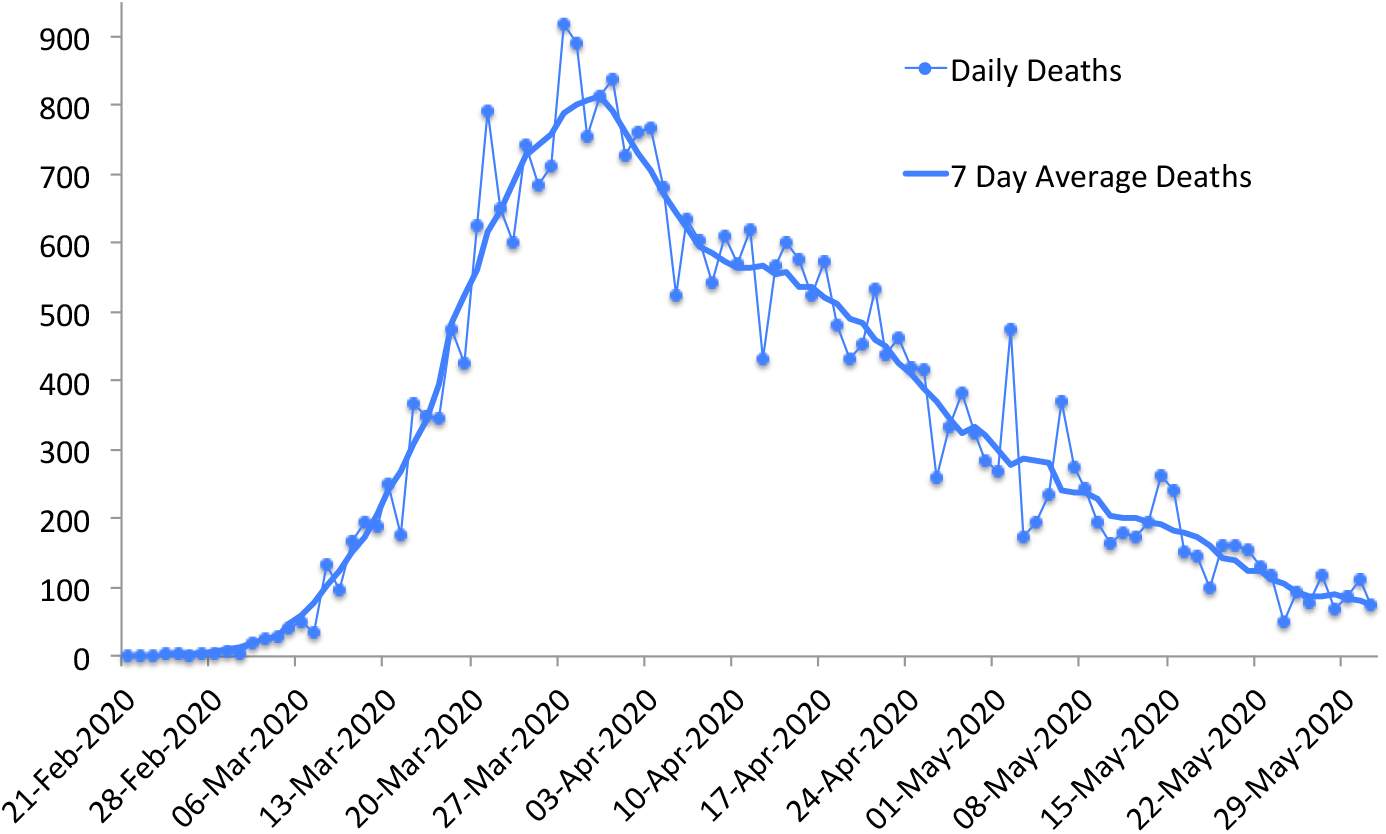
Daily Covid-19 Deaths in Italy 28 Feb to 31 May 2020

The cumulative deaths for the same period^6^ are shown in Figure 2. Figure 3 shows how well the Gompertz Function Model fits the cumulative Covid-19 Deaths data.

**Figure 2:**
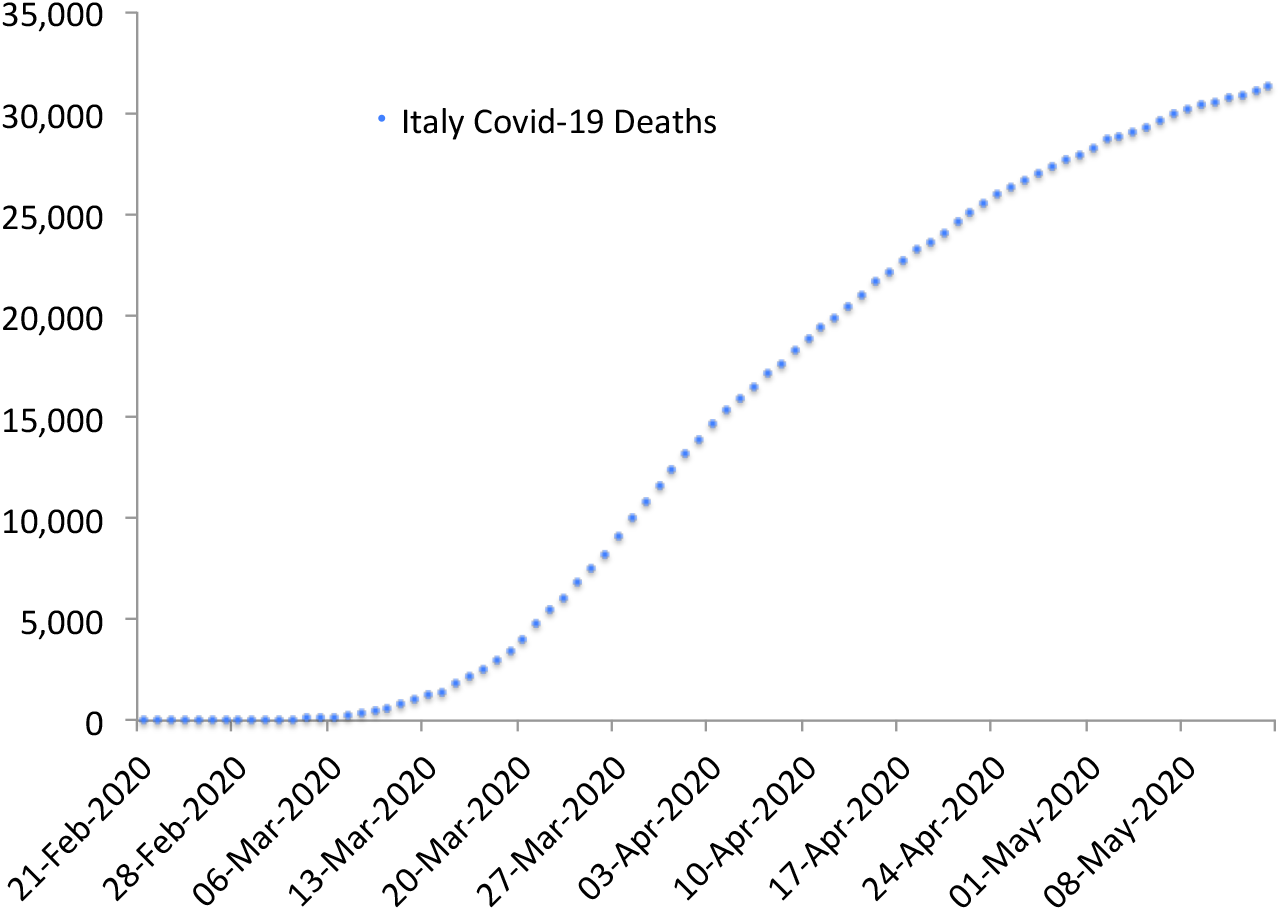
Cumulative Covid-19 Deaths in Italy 28 Feb to 14 May 2020

**Figure 3:**
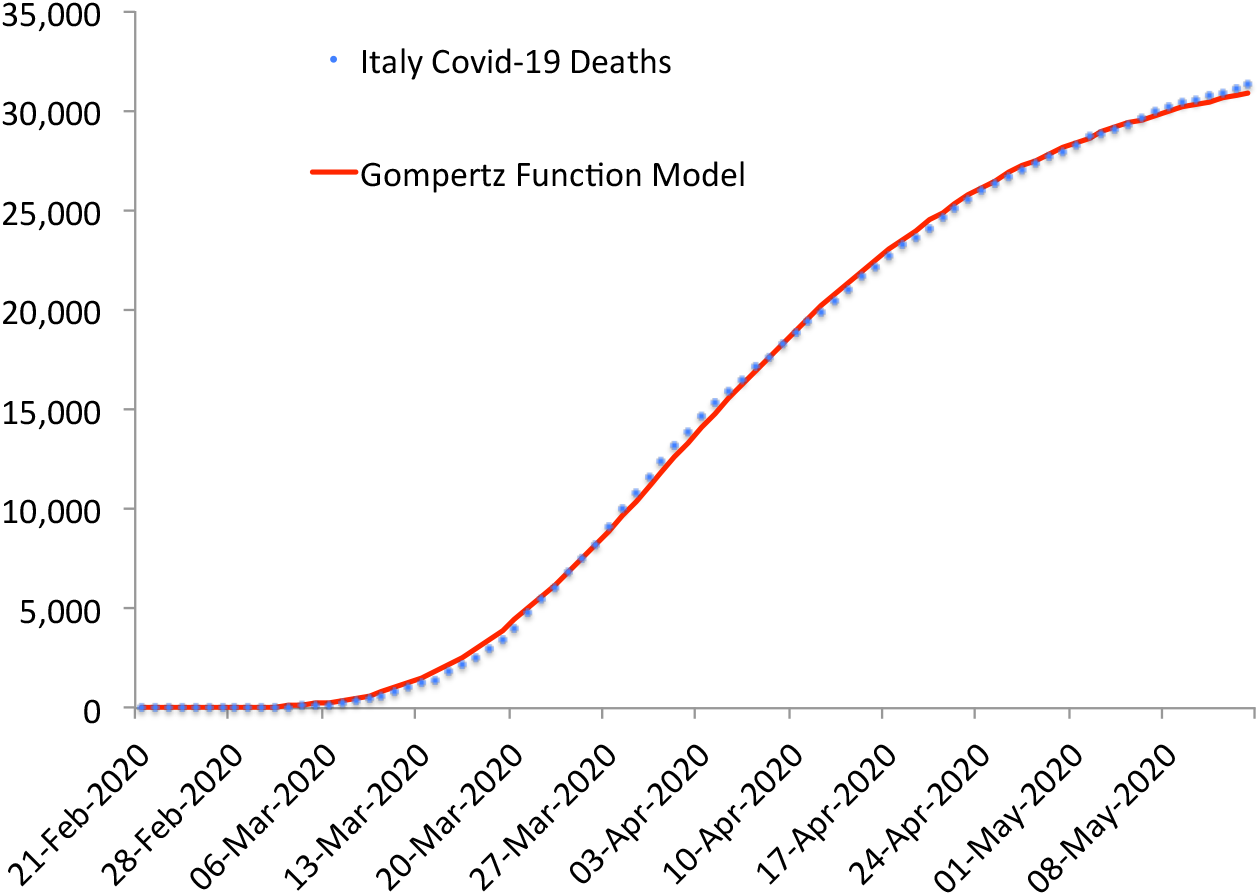
Gompertz Function fit to Cumulative Covid-19 Deaths in Italy 28 Feb to 14 May 2020

The daily differences between Gompertz Function values are shown together with the 7-Day average of daily deaths in Figure 4.

**Figure 4:**
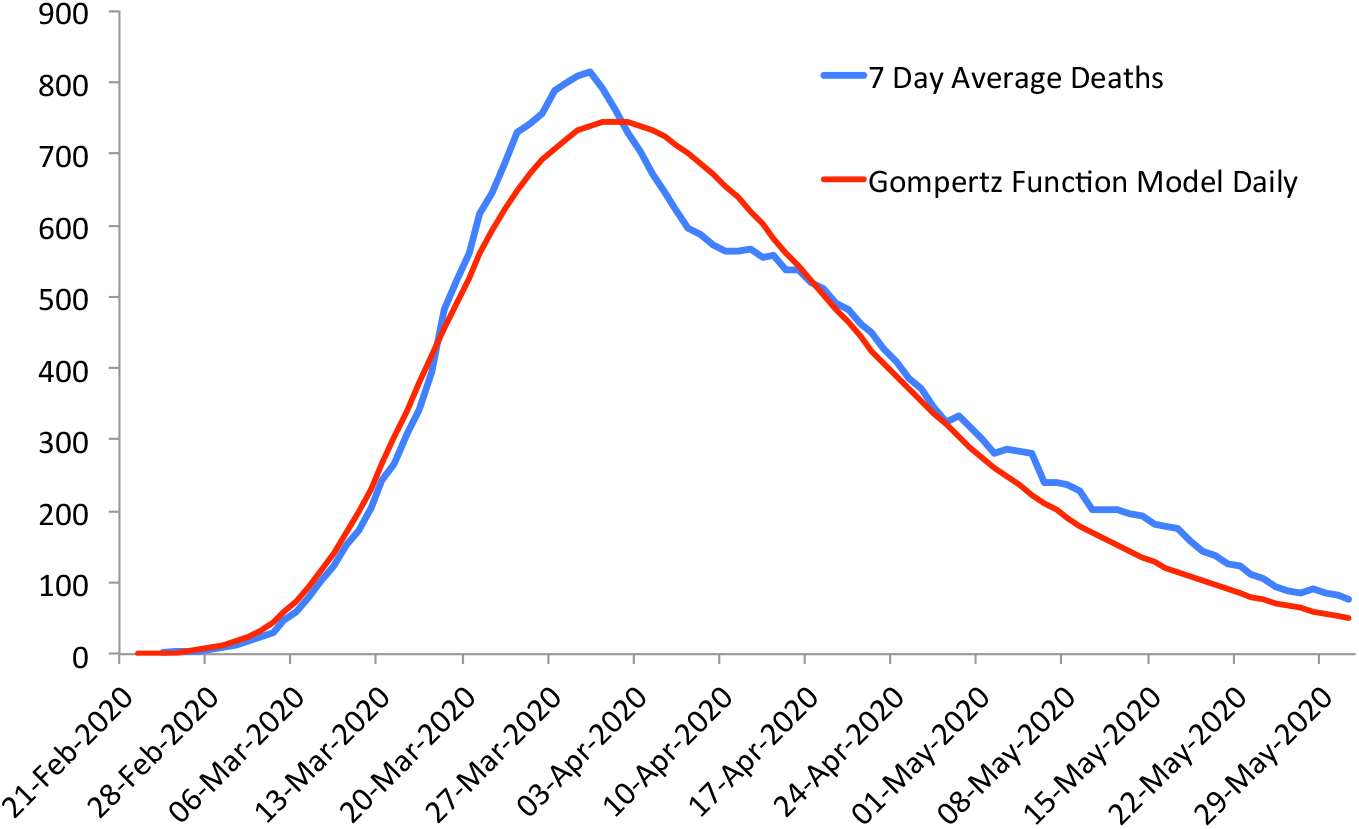
Gompertz Function Model (peak on 1 April) and 7 Day Average (peak on 30 March) of Daily Covid-19 Deaths in Italy 28 Feb to 31 May 2020

Levitt and his co-authors [13] showed subsequently that in hundreds of countries and states worldwide the Gompertz Function Models obtained by fitting observed data could be used to predict final case and death levels as the epidemic continued and more data became available.

This should not have come as a surprise. It is precisely what the historical record told us to expect, as we show in the next section.

### 2.2 What Data Tell Us in Examples of Infectious Disease Outbreaks Prior to 2020

In this section we show that the Gompertz Function Model is a generically good description of epidemic data from a wide variety of epidemics over the past 170 years.

In each case we display the ‘best fit’ of a Gompertz Function Model to the cumulative epidemic data at the end of the outbreak, where the best fit is determined by non-linear regression using a standard Python routine.^7^,^8^

It is important to note that the quality of the Gompertz Function fits shown in all the examples here implies that the strong asymmetry we noted in Italy’s Covid-19 deaths data is, to a good approximation, a regular feature of epidemic growth.

Figures 5-11 show the course of epidemic outbreaks as varied as: Cholera in Aalborg Denmark in 1853,’Spanish Flu’ in Prussia and San Francisco in 1918, SARS CoV-1 in Hong Kong in 2003, Ebola in Sierra Leone in 2014 and Zika in Antioquia, Colombia, 2016. All of them evolved according to Gompertz Functions.

**Figure 5:**
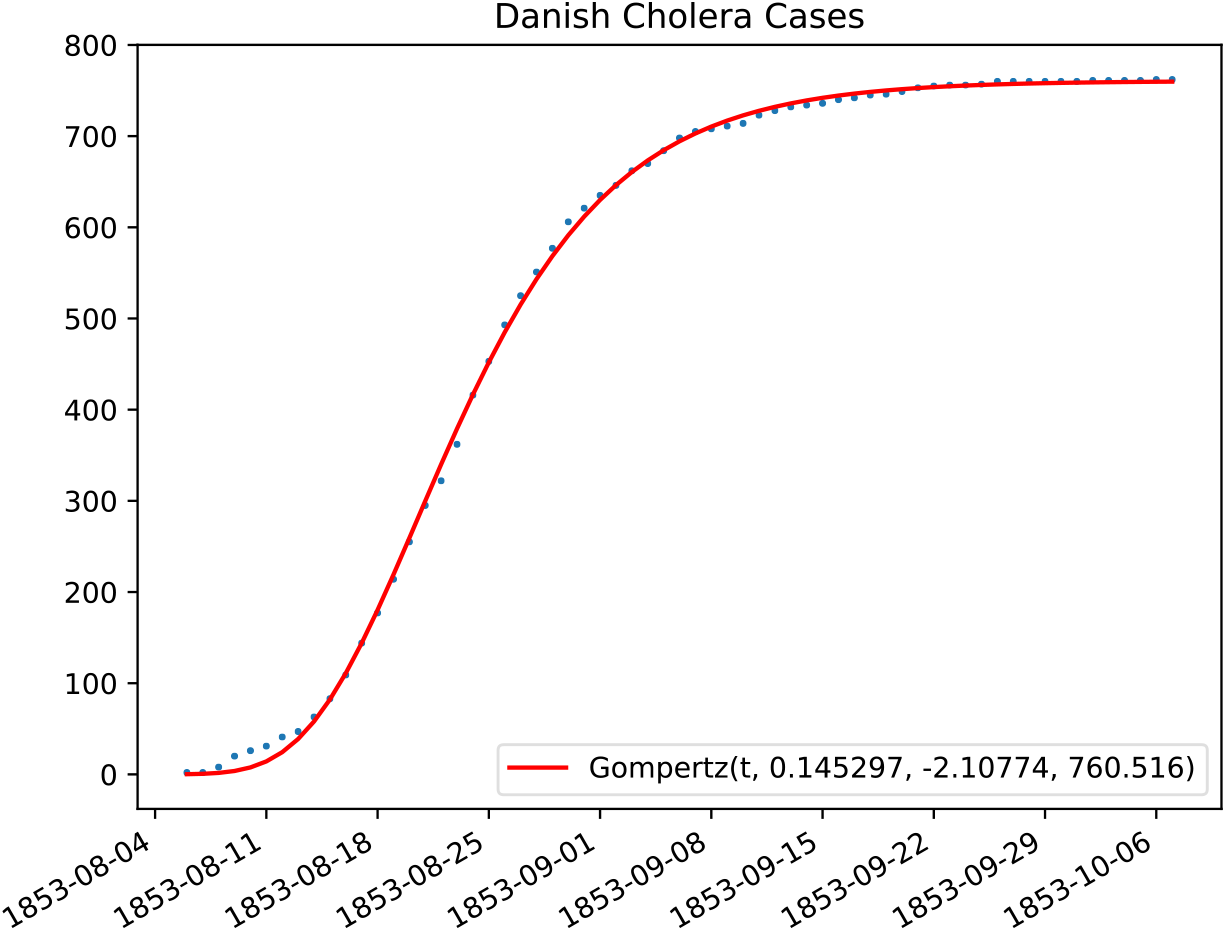
Aalborg Denmark Cholera Cases 1853

**Figure 6:**
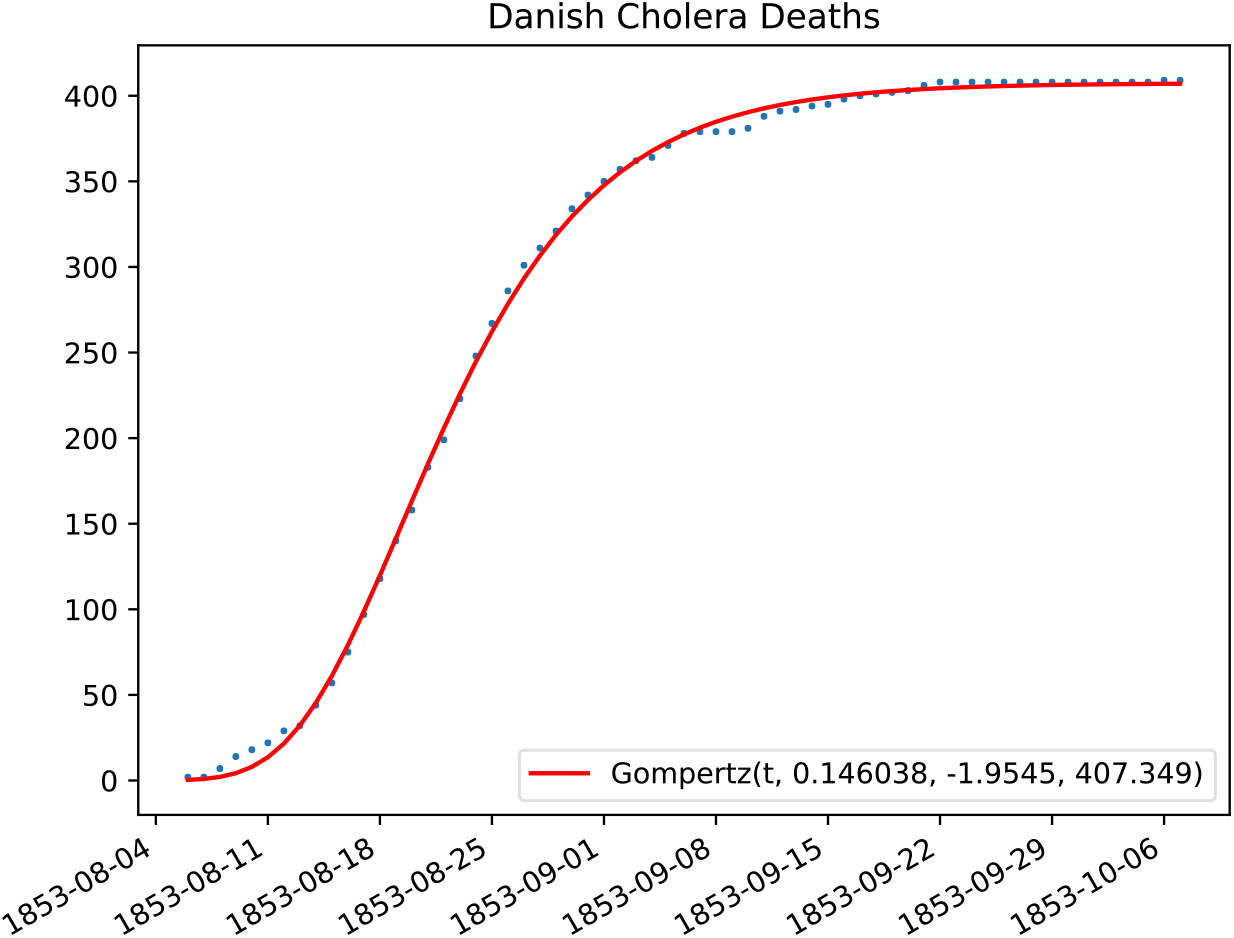
Aalborg Denmark Cholera Deaths 1853

**Figure 7:**
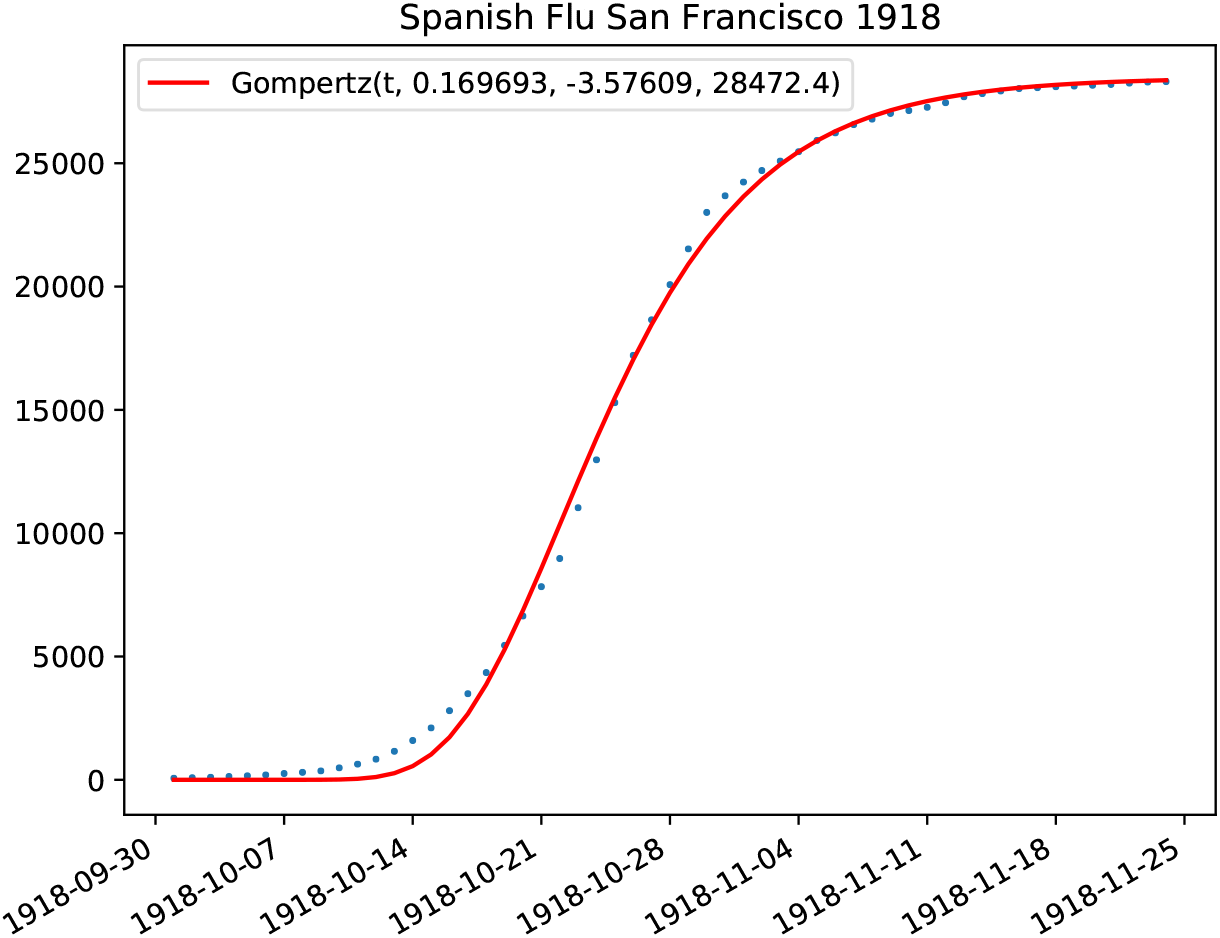
‘Spanish Flu’ Cases San Francisco 1918

**Figure 8:**
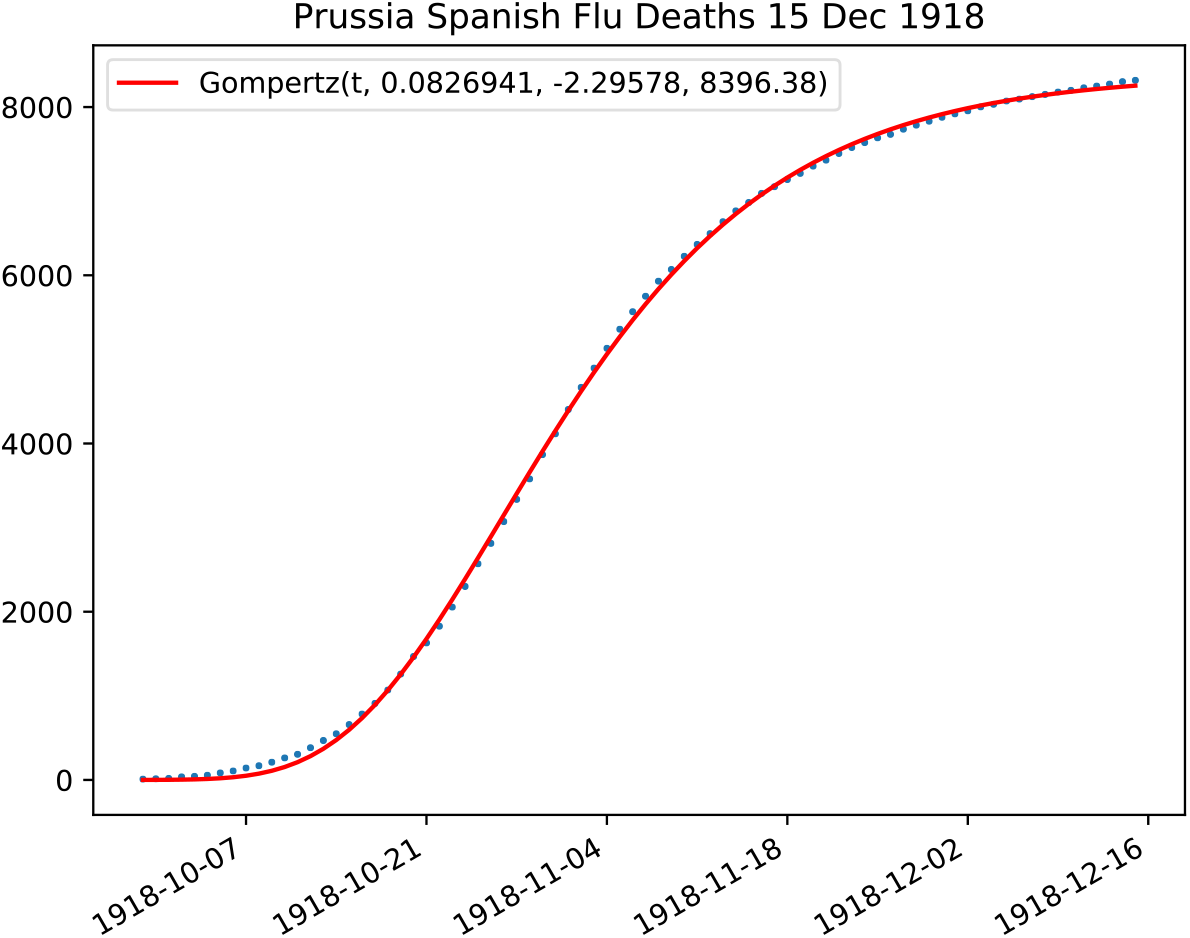
‘Spanish Flu’ Deaths Prussia 1918

**Figure 9:**
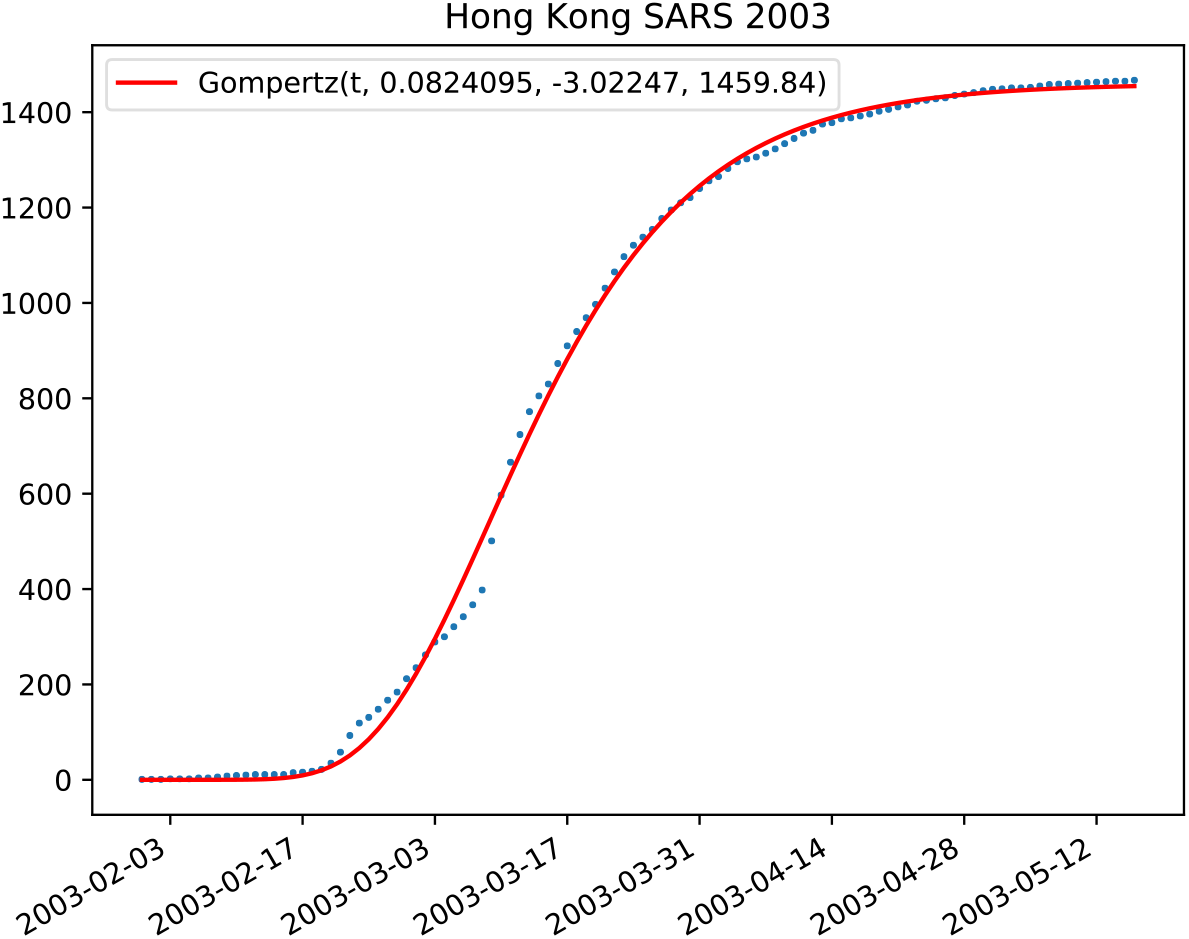
Hong Kong SARS Cases 2003

**Figure 10:**
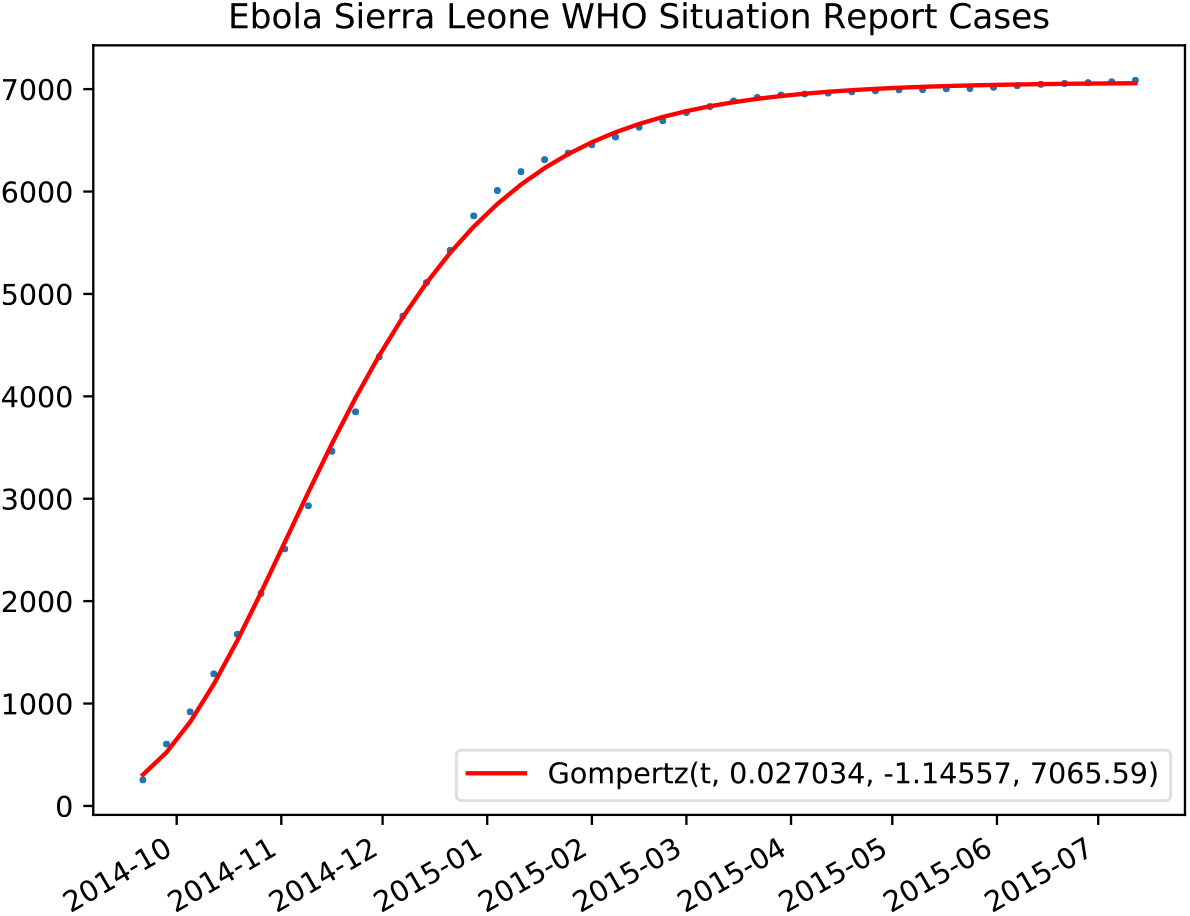
Ebola Cases Sierra Leone 2014-2015

**Figure 11:**
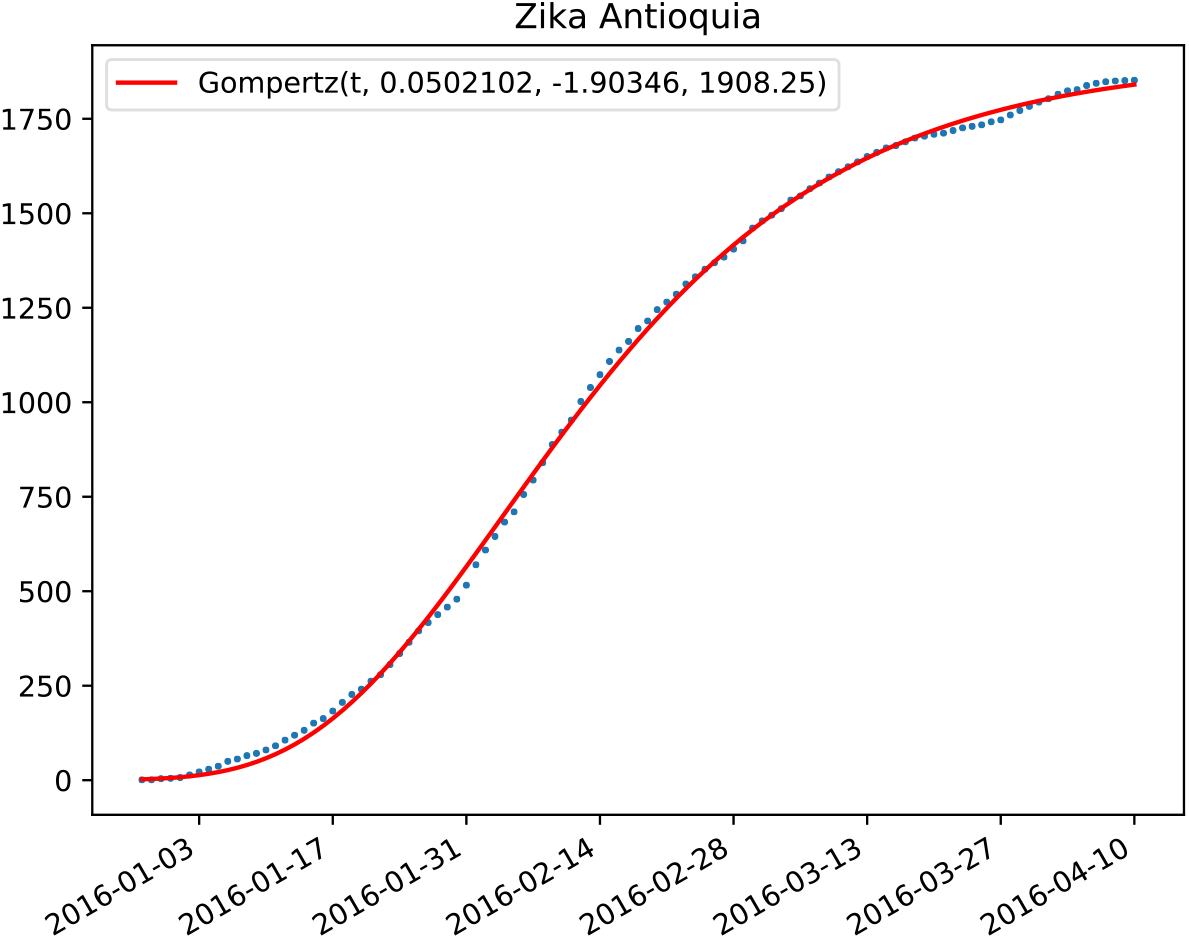
Zika Cases, Antioquia Colombia 2016

**Figure 12:**
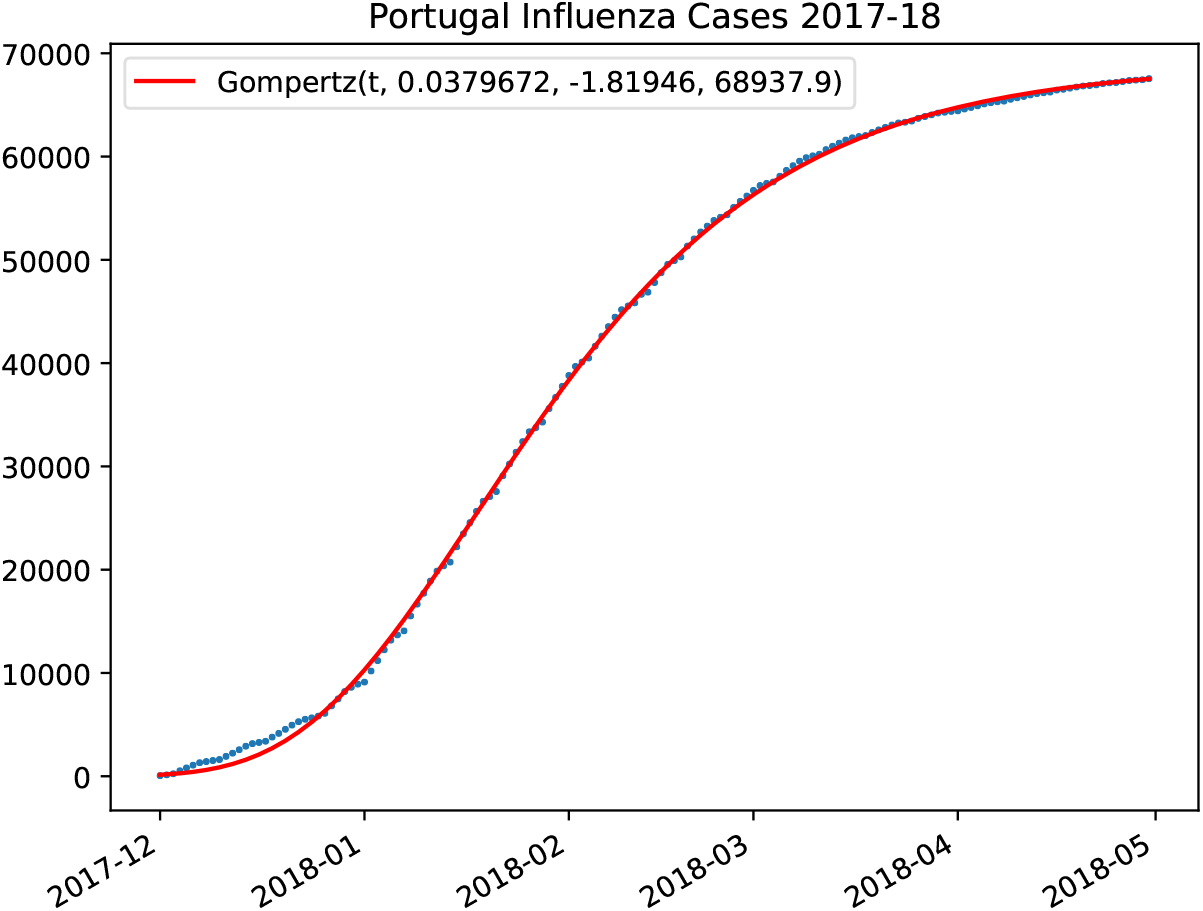
Influenza Cases in Portugal from 1 Dec 2017 to 30 Apr 2018

**Figure 13:**
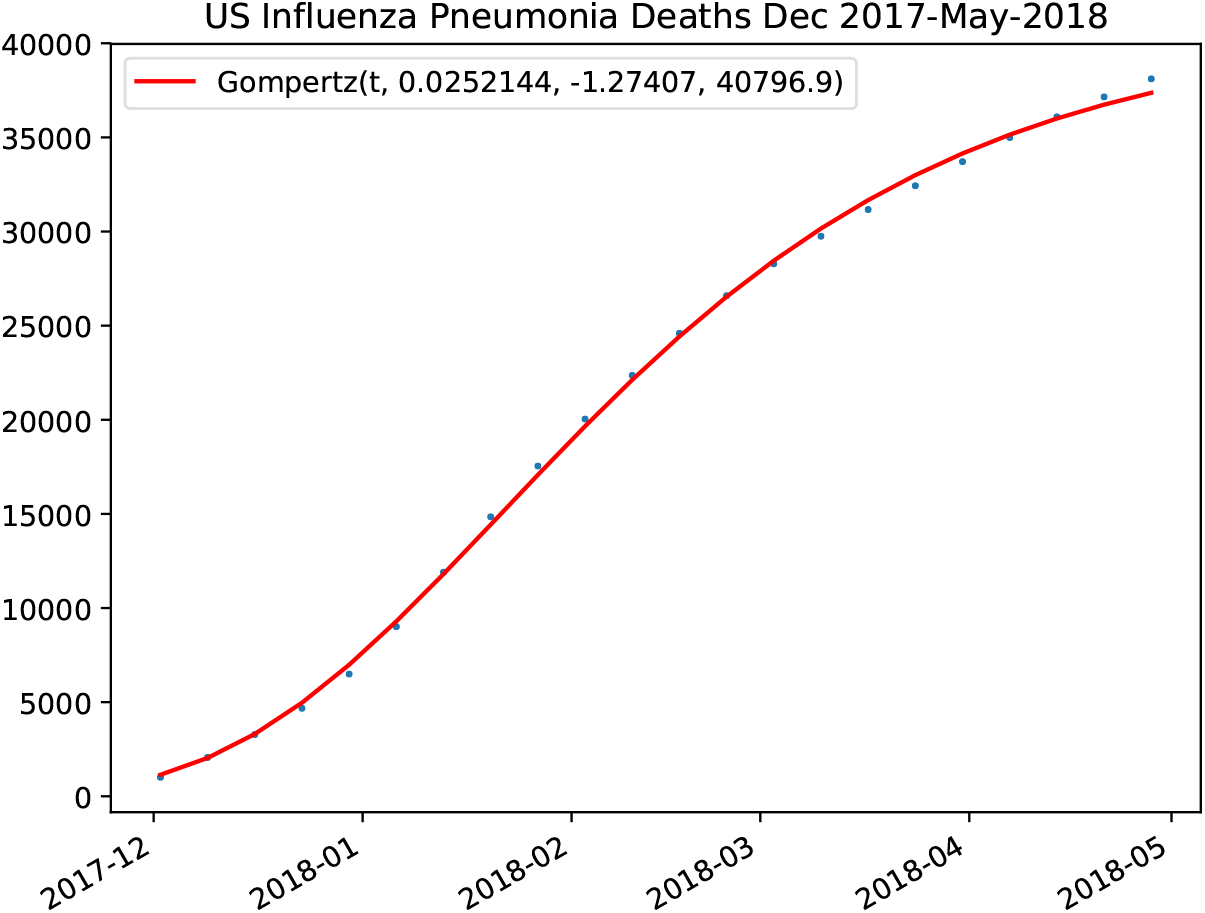
Influenza-Pneumonia Deaths USA Dec 2017-May 2018

In addition, Figures fig:PortugalInfluenza17-13 show that the following examples of outbreaks of endemic disease: Influenza Cases in Portugal and Influenza-Pneumonia Deaths in U.S.A., both in 2017-18,^9^ all evolve to follow Gompertz Functions.

From Denmark in 1853, a time when even the bacterial origin of Cholera was still to be discovered, through the ‘Spanish Flu’–the greatest pandemic of modern times–to major bacterial and viral epidemics prior to Covid-19, the data shows us that the Gompertz Function has been an accurate description of the outcome of *all* of these outbreaks.

### 2.3 Predictive Power of the Gompertz Function Model in Examples Prior to 2020

The agreement between the cumulative cases or deaths and the Gompertz Function Model in all of these examples is remarkable. In itself it allows us to make deductions about the dynamics of the outbreak, based on those of the Gompertz Function (which we take up in Section 3).

In these examples, the final outcome is, to a very good approximation, a Gompertz Function. But the important question for us is the extent to which the Gompertz Function Model allows us to predict the future, that is to use observations of the data up to the current time to make accurate predictions about what we will see in the next days or weeks.

To assess the predictive power of the Gompertz Function model we need to know if the best Gompertz Function fit at time *t* allows us to predict numbers for some time *t* + *τ* into the future with a reasonable degree of accuracy. For concreteness we’ll use an error of *±*10% as our desired level of accuracy and measure the time *τ* for which a fit remains within that error.

We illustrate this process in Figure14 for Cholera in Denmark in 1853, in Figure 15 for Ebola in Sierra Leone in 2014-15 and in Figure 16 for the Portuguese 2017-18 Influenza season.

**Figure 14:**
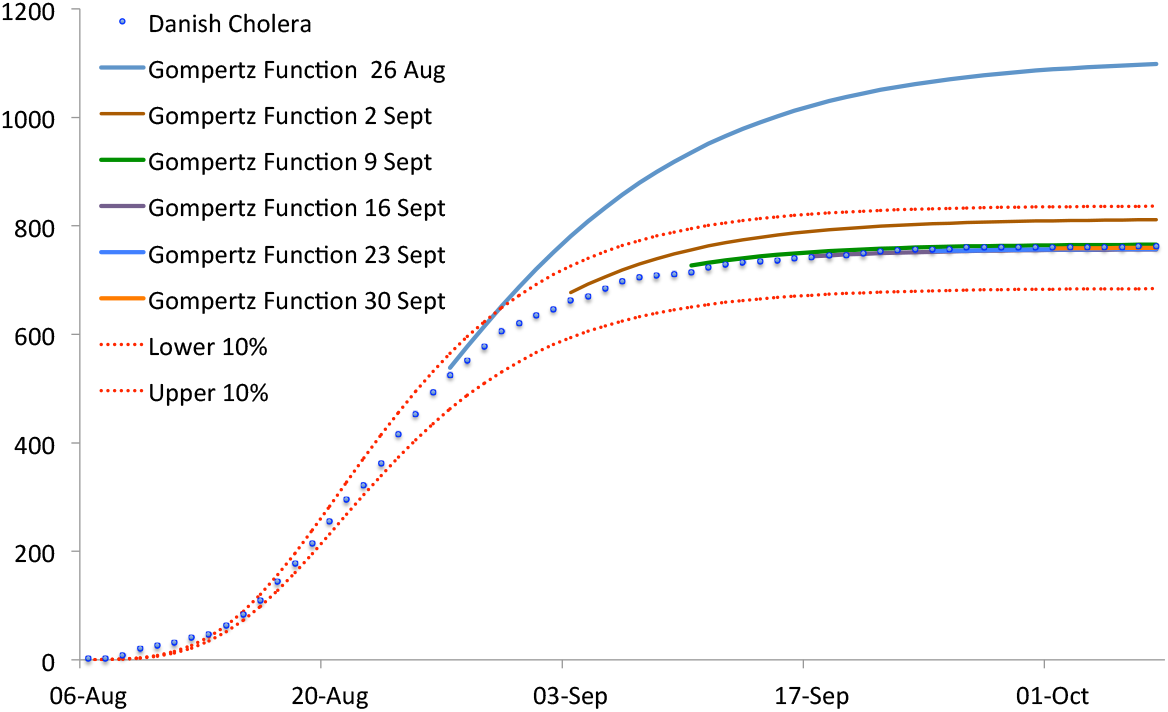
Sequence of weekly updates of Gompertz Function fits to Danish Cholera Cases

**Figure 15:**
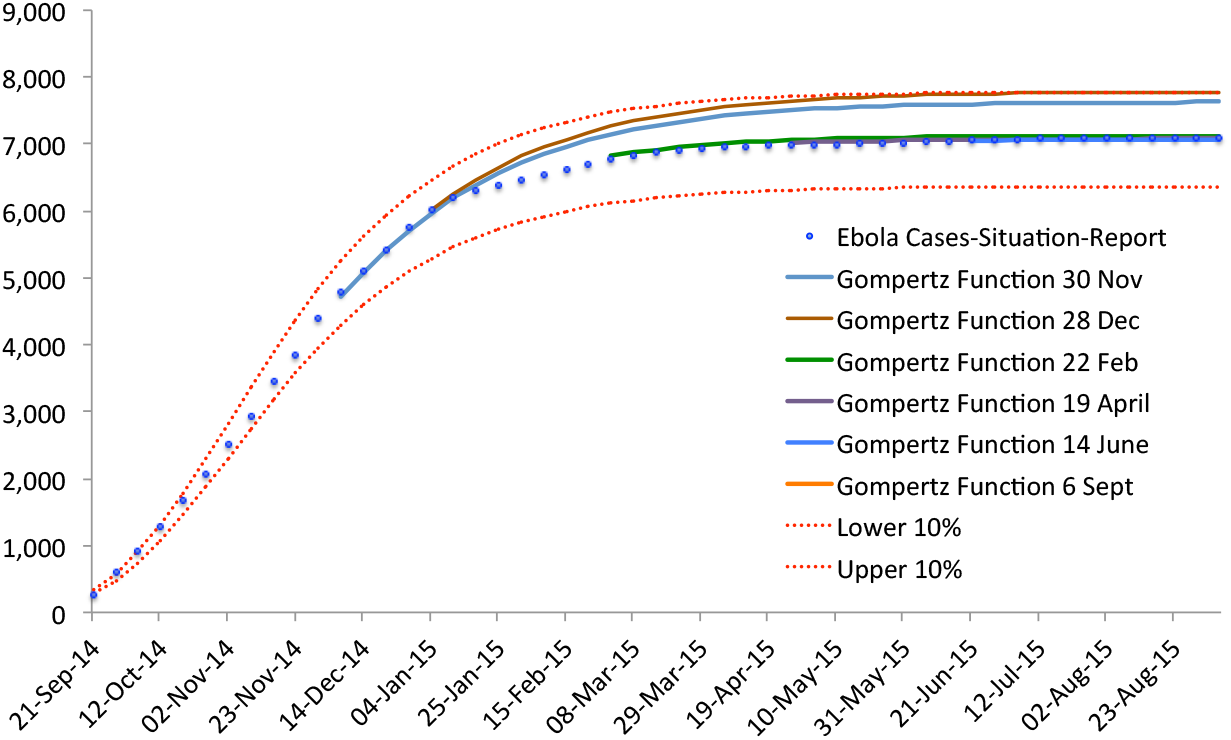
Sample of 4-weekly updates of Gompertz Function fits to weekly Ebola Case data, Sierra Leone 2014-15

**Figure 16:**
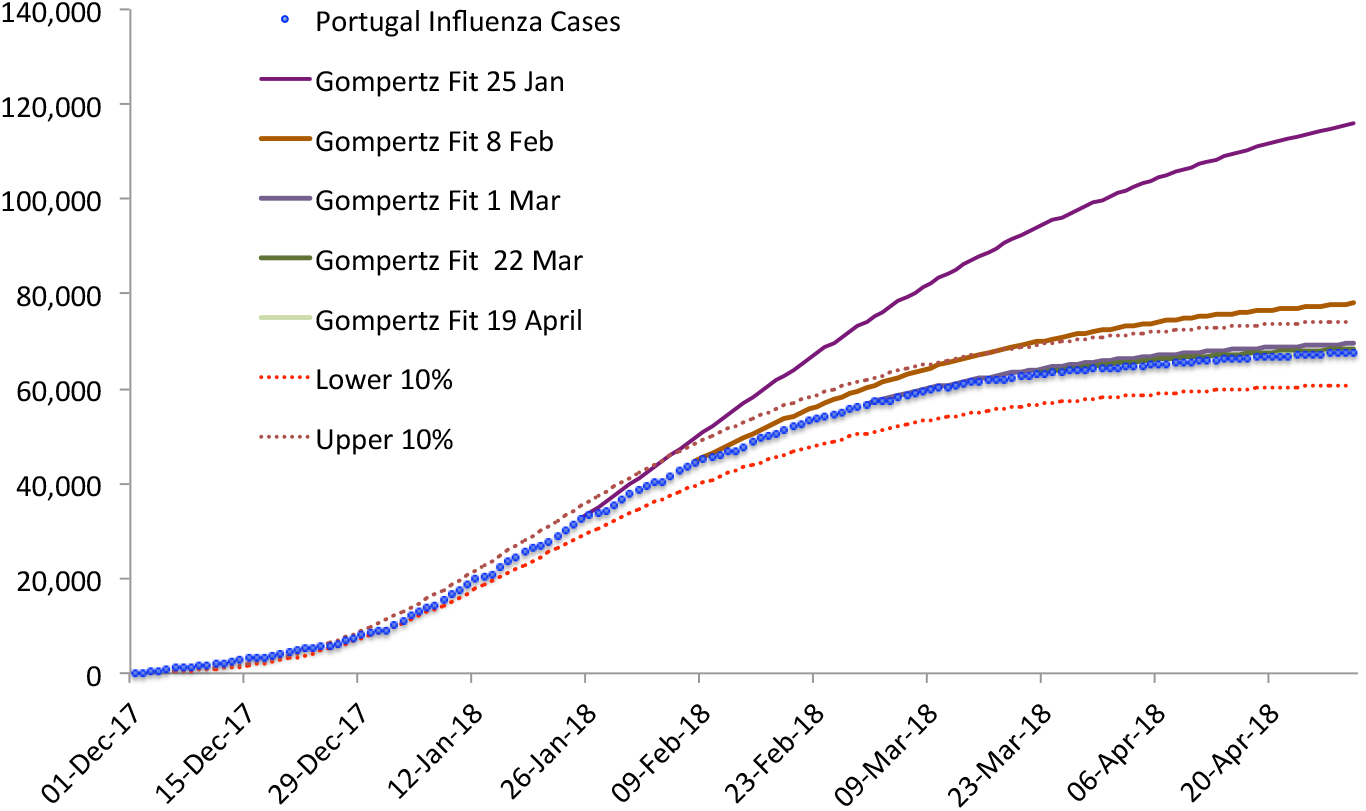
Sequence of weekly updates of Gompertz Function fits to Portuguese Influenza Cases

To simplify the figures, only the out of sample part of the Gompertz Function is graphed. Error bands of *±*10% around the data show how quickly a high level of accuracy is achieved.

To judge the quality of successive predictions, we want to measure the gap between the predicted value *X*(*t*) and the observed value *X*_*obs*_(*t*) as a percentage of *X*(*t*). In other words we want to judge the extent to which the observation differed from the prediction, as a percentage of the prediction.^10^ We will use the absolute value of the error to simplify the graphs. So we display the error *ϵ* in % defined as

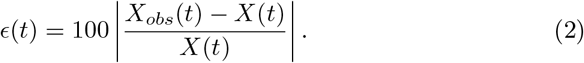

#### 2.3.1 Danish Cholera Cases

The Danish Cholera epidemic evolved very rapidly. The case data begin on 6 August and by 6 October 1853, less than 9 weeks later, the epidemic was over.

The first Gompertz Function fit on 26 August, as Figure14 shows, soon proved over-pessimistic. Its out of sample error was less than 10% for only 3 days. Nevertheless even this early fit would have been useful. The error remained less than 20% for 12 days.

Figure 17 shows how quickly the predictive power of subsequent fits increases. The fit made one week later, on 2 September, was never out by more than 6%. The remainder of the weekly update fits had errors of less than 1% for the duration of the epidemic.

**Figure 17:**
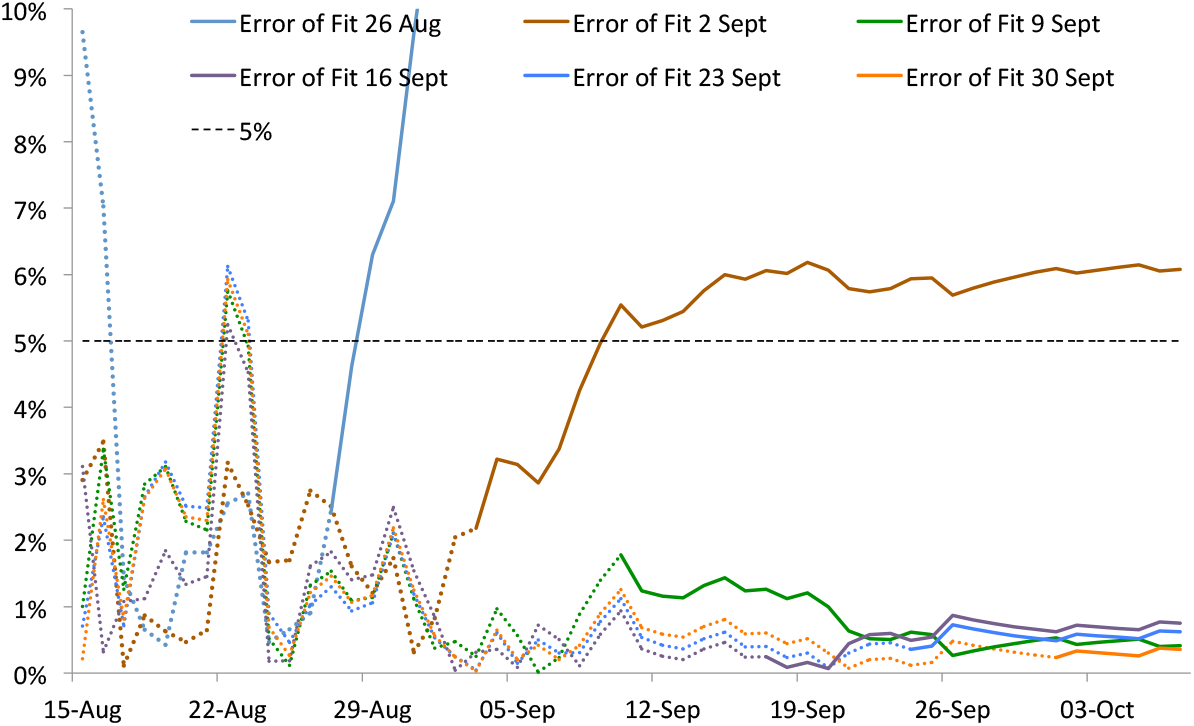
Evolution of the in and out of sample errors in the Gompertz Function fits to Danish Cholera Cases.

#### 2.3.2 Ebola Sierra Leone

We have WHO Situation Reports with weekly updates on Ebola cases in Sierra Leone for 52 weeks from 21 September 2014 to 13 September 2015.

The first fit (not shown) was done with six weeks of data (i.e. with only 6 data points) on 26 October. Its out of sample error was less than 10% for only 1 week but remained below 15% for 2 weeks. Subsequent fits gained predictive power rapidly. For the fit made two weeks later the error was less than 10% for 2 weeks and below 15% for 3 weeks

The error from the first fit shown in Figure18 (November 16 2014) with only 9 data points was never out by more than 11%. Subsequent updates to the Gompertz Function fits were done every 4 weeks. Their out of sample errors all remained below 10% for the duration of the outbreak.

**Figure 18:**
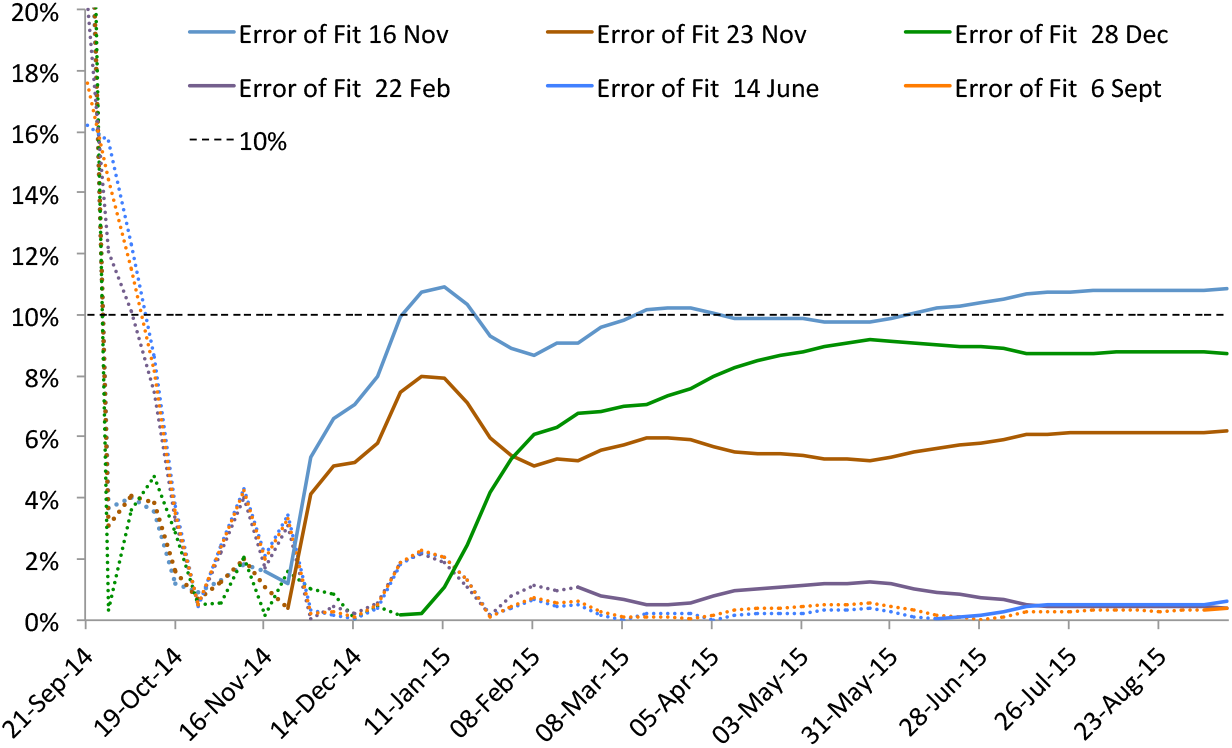
Evolution of the in and out of sample errors in the Gompertz Function fits to Ebola Cases.

**Figure 19:**
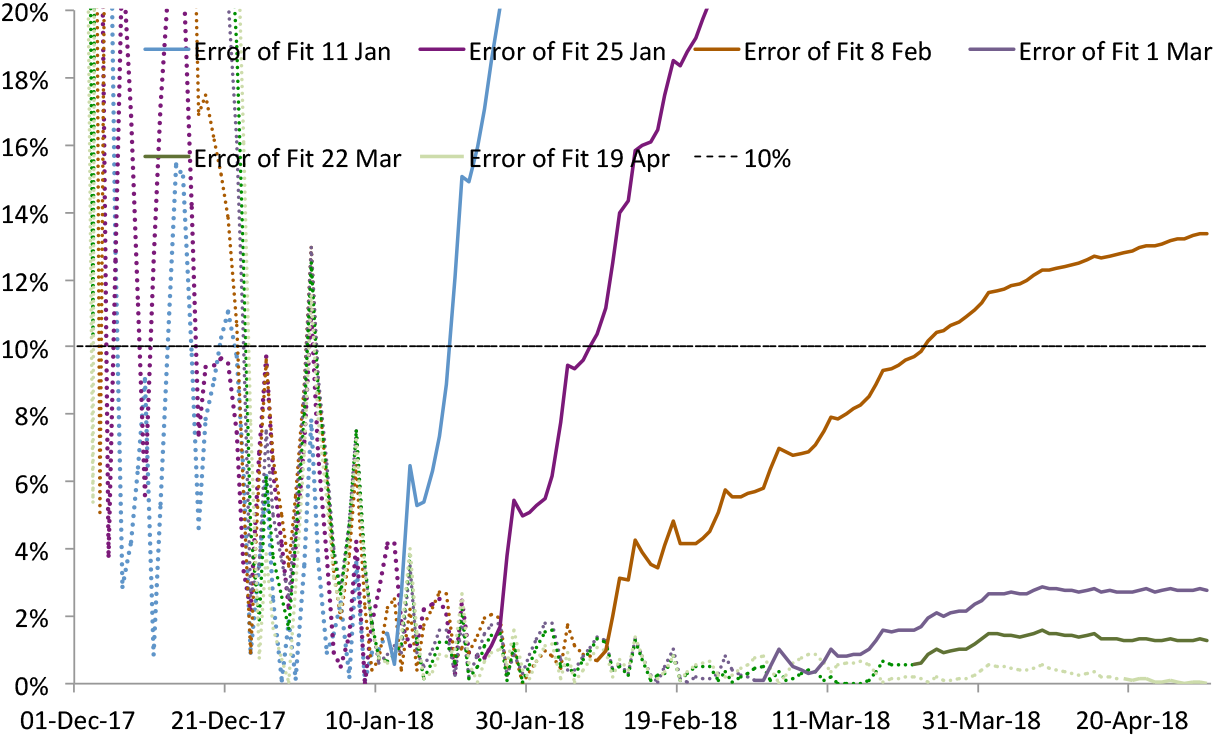
Evolution of the in and out of sample errors in the Gompertz Function fits to Portuguese Influenza Cases

#### 2.3.3 Portuguese Influenza Cases

The Portuguese Influenza Case data begins Gompertz Function growth on 1 December 2017 and continues until 30 April 2018. The out of sample error of the 11 January fit remained below 20% for eleven days but was below 10% for only 5 days.

For the fit made two weeks later, the out of sample error remained below 10% for more than a week. From 1 February’s fit this level was maintained for almost 3 weeks out of sample.

These examples illustrate the predictive power that precedes the final excellent Gompertz Function fits to the observed data. The differences in the time taken to reduce out of sample errors to our 10% target depend on the parameters *a* and *b* in a way that we will return to after outlining the dynamics of the Gompertz Function.

First we examine the performance of the model in the 2020 Covid-19 epidemic across a variety of locations.

### 2.4 Covid-19 Epidemic 2020

Covid-19 has produced an unprecedented volume of data from countries all over the world.

In addition to case and death data, many locations published data on the medical consequences of the outbreak: hospital and intensive care admissions for Covid-19, and/or daily updates of total hospital and intensive care patients, numbers on ventilators etc.

Any process driven by the epidemic–such as hospitalisations–that also follows Gompertz Function growth will be predictable.

We should expect that cumulative hospitalisations will evolve as a percentage of cases. Likewise a certain percentage of Covid-19 patients will be admitted to Intensive Care Units or other specialised treatment. But these percentages cannot be known in advance and may vary over time.

The Gompertz Function Model doesn’t require that we know, estimate or approximate these percentages in advance. As data accumulate (typically two weeks of daily observations is enough to start) Gompertz Function fits can be obtained and tested for their predictive power. Based on previous epidemics, we can expect predictive power to increase quickly as time goes on.

We have used hospital and death data as our primary illustrations rather than cases. Hospital admissions generally involve a diagnosis of illness rather than a simple test for the presence of the virus so they should be expected to provide a good proxy for the progress of serious cases of infection. By contrast, case data are subject to considerable uncertainty (regardless of testing regime) and are frequently revised retrospectively.^11^

These data are available in many locations with a short lag. This means that historic data for hospitalisations and intensive care admissions will be a very good approximation of the data and model performance we would have seen in real time, as long as the lags are taken into account.

For the Northern Hemisphere, the locations we use to illustrate the Gompertz Function fits are, in order of decreasing latitude, Sweden, London, Isle de France, the Province of Ontario and Portugal. In the tropical Southern Hemisphere we use the State of Rio de Janeiro. These are all of roughly the same order of magnitude in population (approximately 10 to 15 million people).

For an equatorial example we use the Brazilian State of Amazonas (approximately 4 million people).

We have been able to obtain data for Hospital admissions in all of these locations and for ICU admissions in all but London and Ontario. We have data for the ICU admissions for England, Wales and Northern Ireland combined, and have used that as a proxy comparison for London.

We also have daily records of the number of Covid-19 patients in Intensive Care and in Ventilator beds for London.These data are well approximated by the derivative 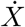 of a Gompertz Function and hence predictable. We illustrate this in Figure 26 where we have made a fit to the cumulative.^12^

Across all of these examples, the Gompertz Function fits are excellent. As a result, the predictive power was also good, exactly as illustrated in the previous epidemics. This is particularly important for making accurate projections of health care loads.

Figures 20 to 41 show the final Gompertz Function fits.

**Figure 20:**
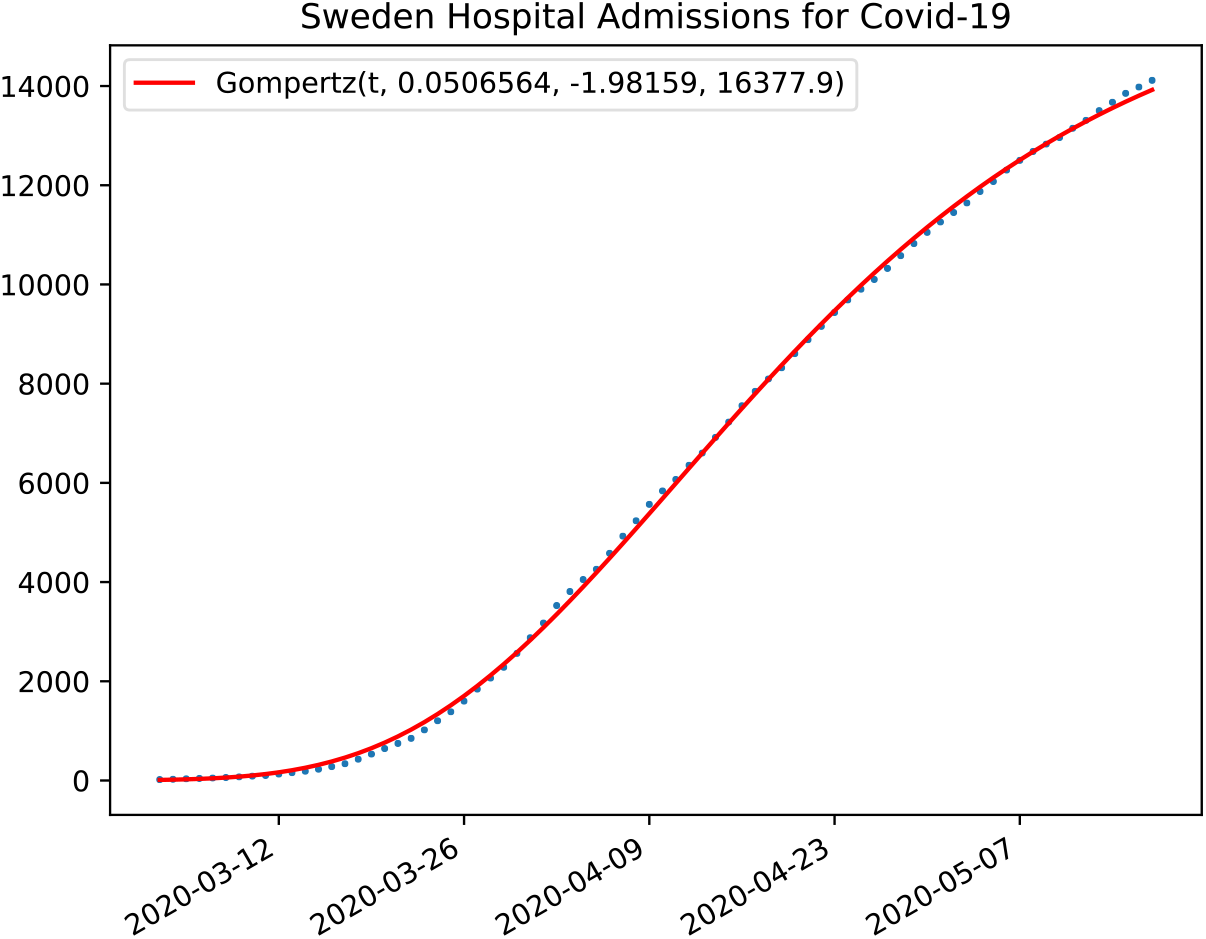
Covid-19 Hospitalisations in Sweden from 3 Mar to 17 May 2020

**Figure 21:**
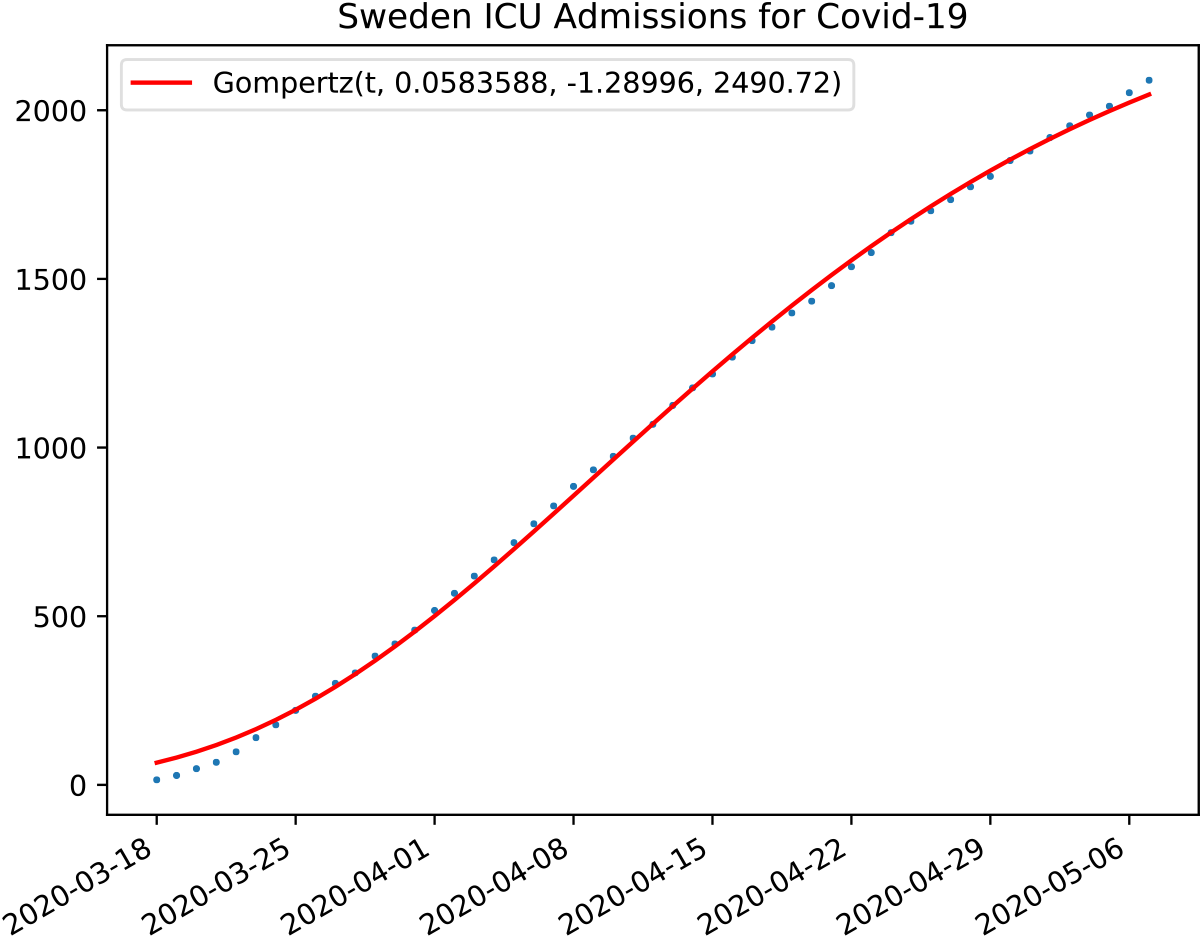
ICU admissions for Covid-19 in Sweden from 18 Mar to 7 May 2020

**Figure 22:**
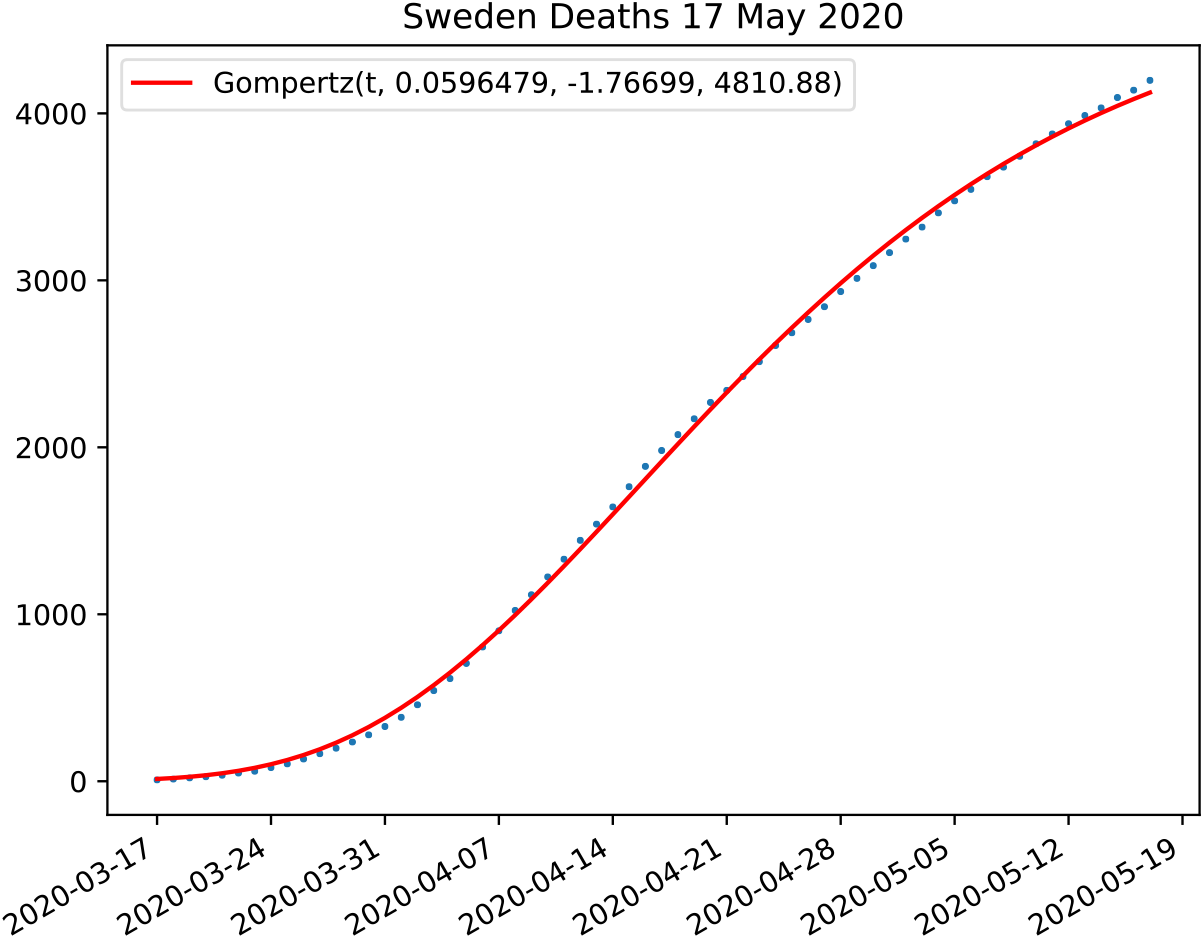
Covid-19 Deaths in Sweden from 17 Mar to 17 May 2020

**Figure 23:**
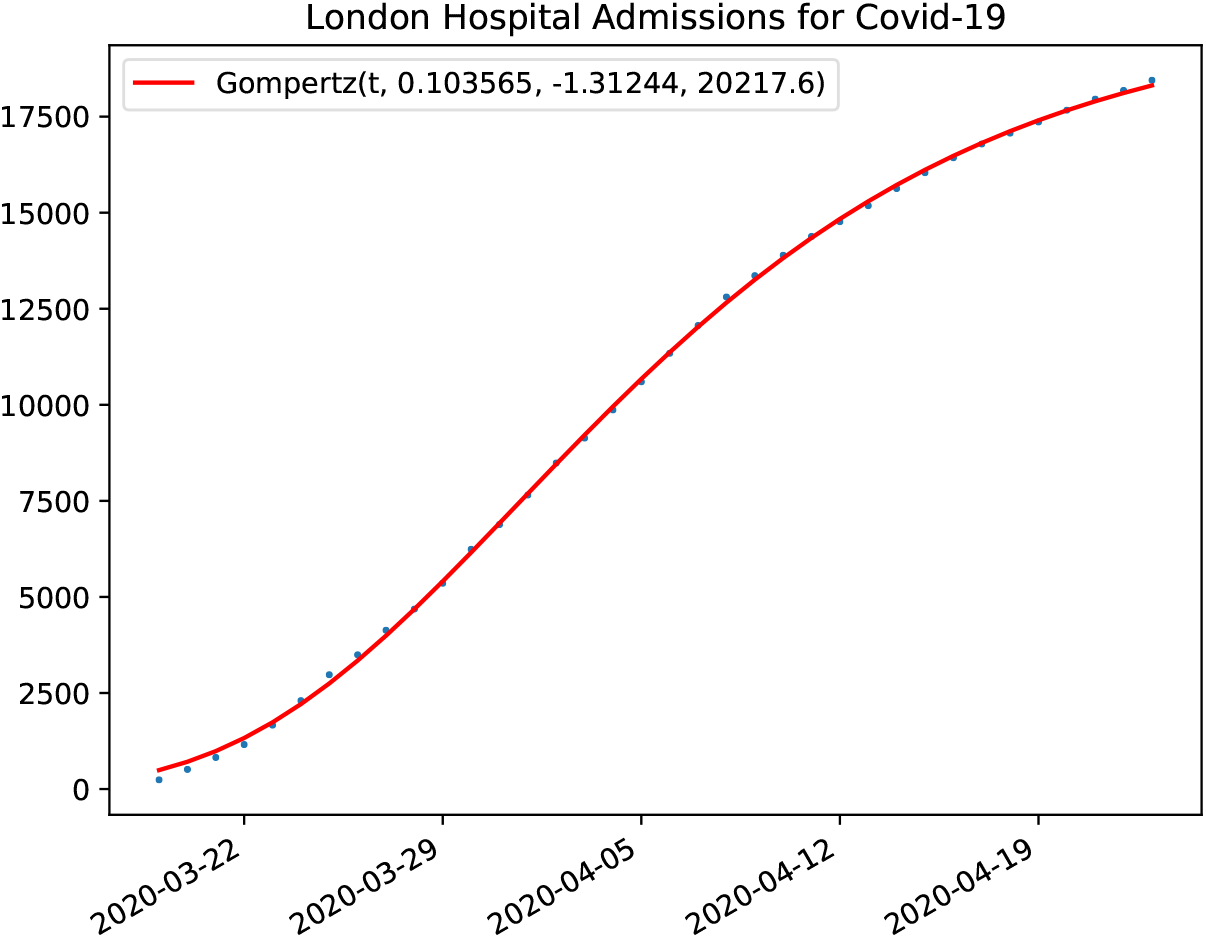
London Covid-19 Hospital Admissions, 19 Mar to 23 April 2020

**Figure 24:**
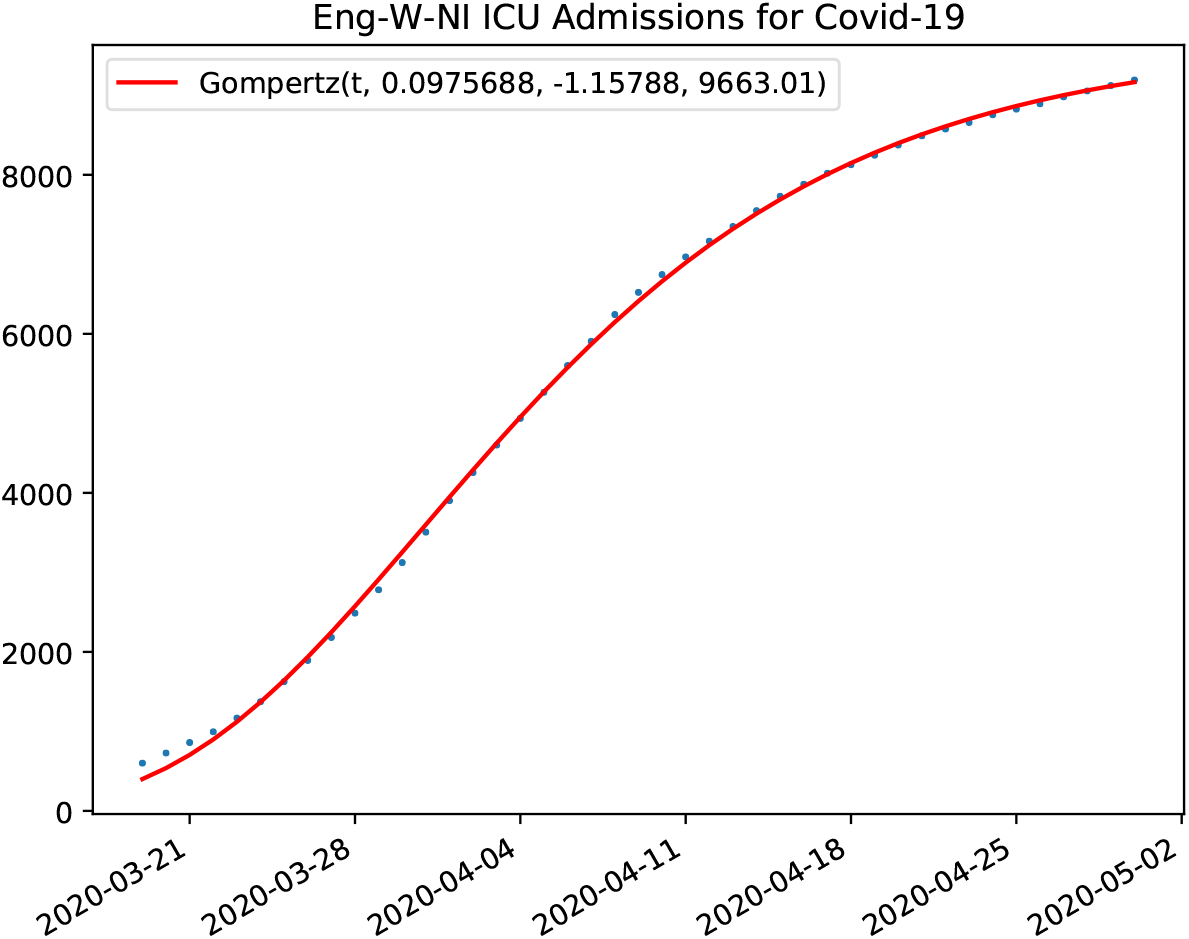
ICU admissions England Wales N. Ireland 19 Mar to 30 Apr 2020

**Figure 25:**
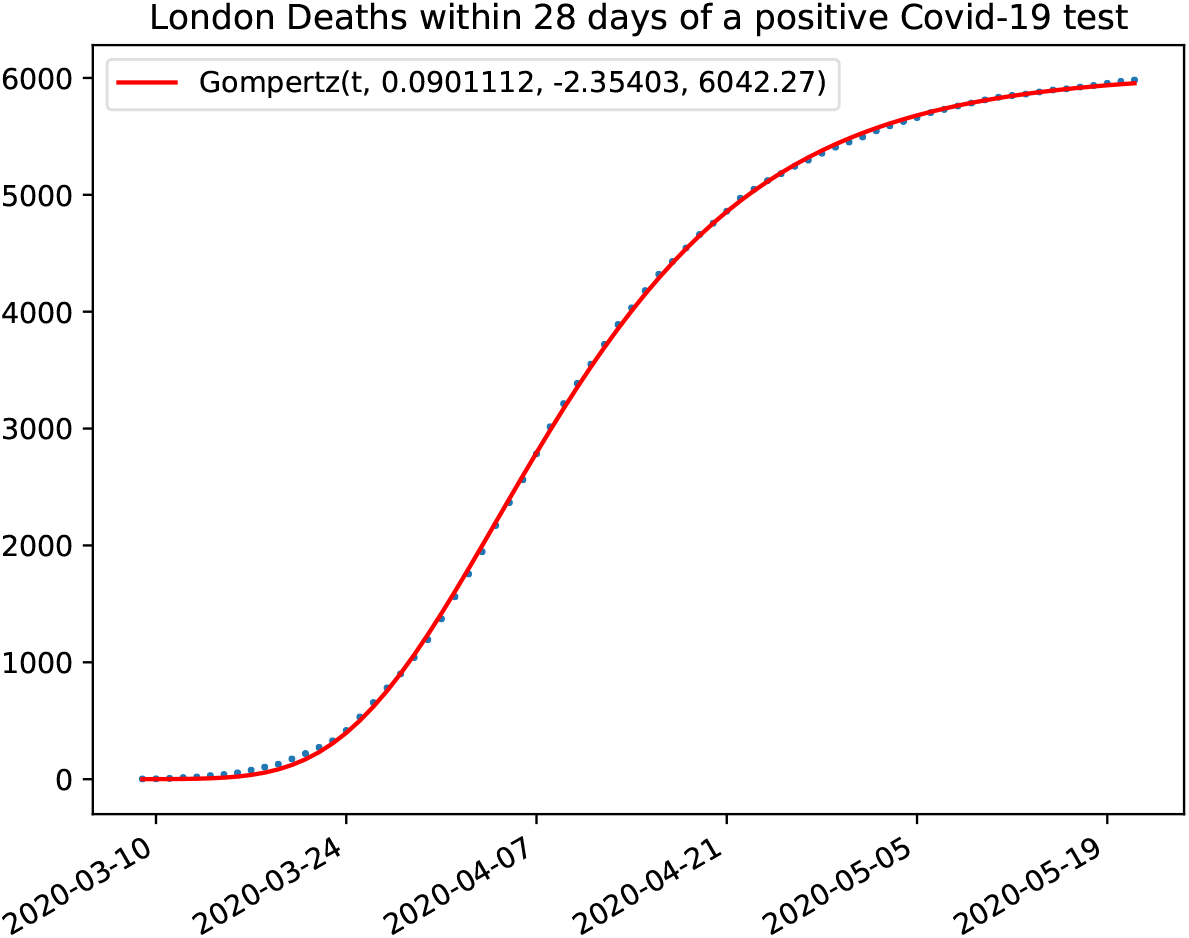
London Deaths attributed to Covid-19 from 9 Mar to 21 May 2020

**Figure 26:**
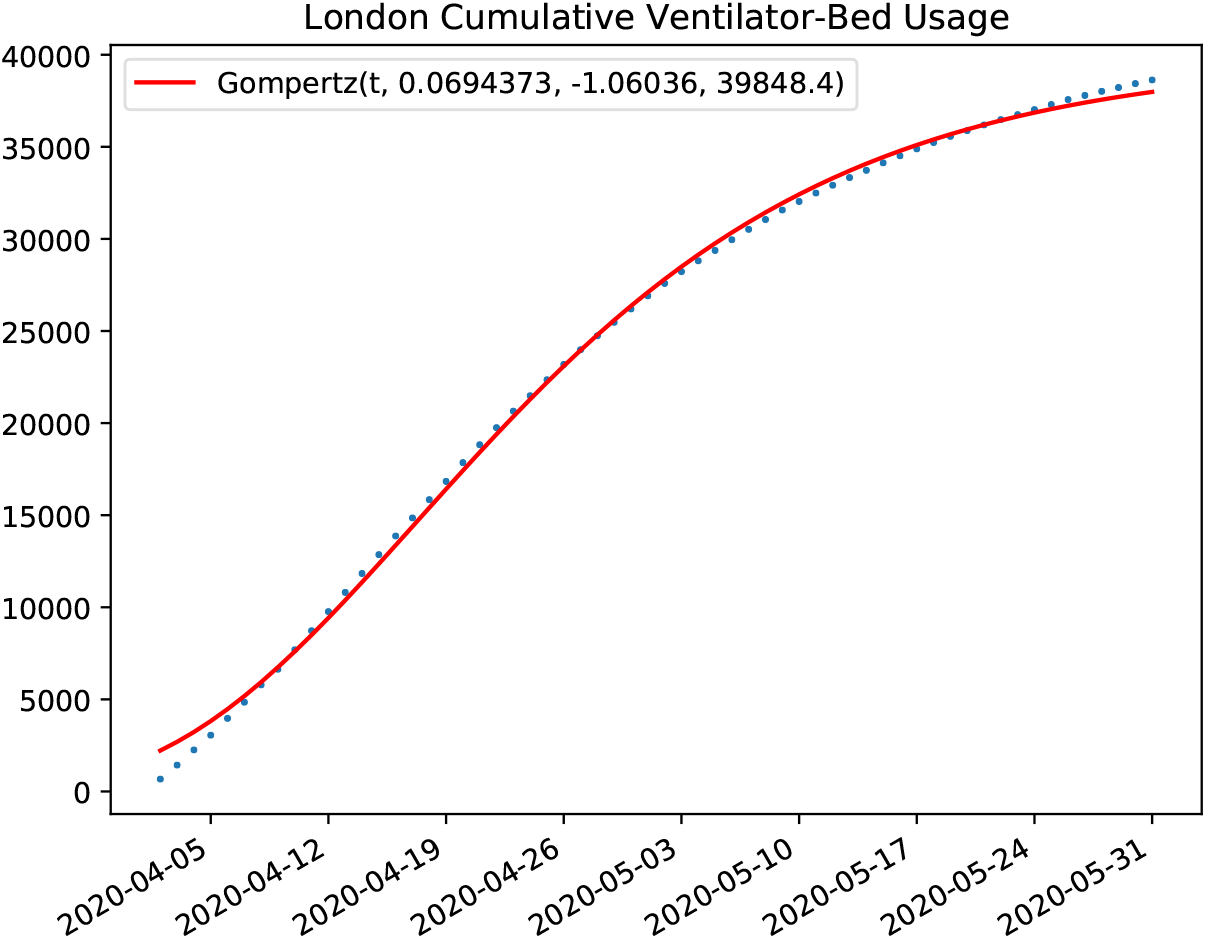
Patient ventilator bed days in London from 2 Apr to 31 May 2020

**Figure 27:**
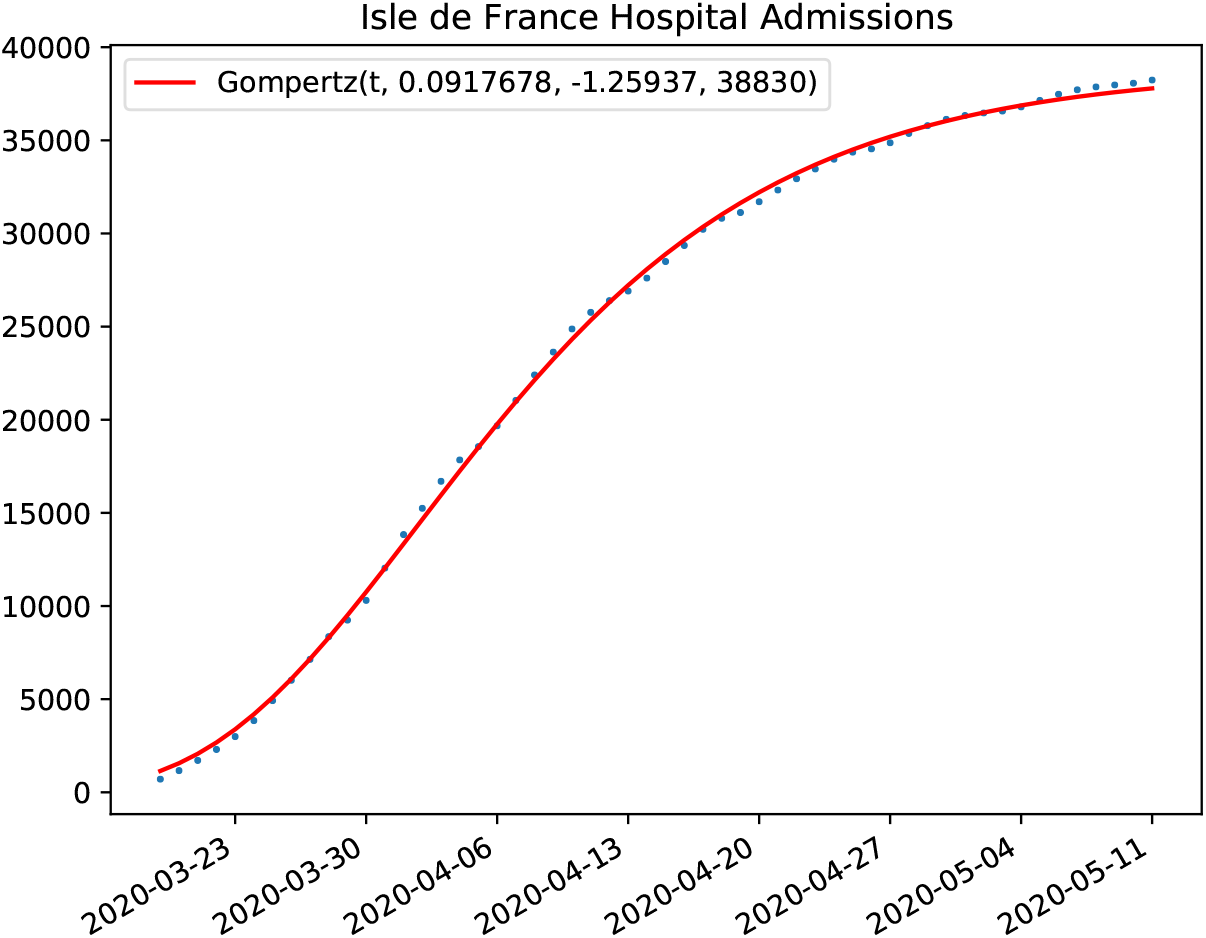
Isle de France Covid-19 Hospital Admissions, 18 Mar to 11 May 2020

**Figure 28:**
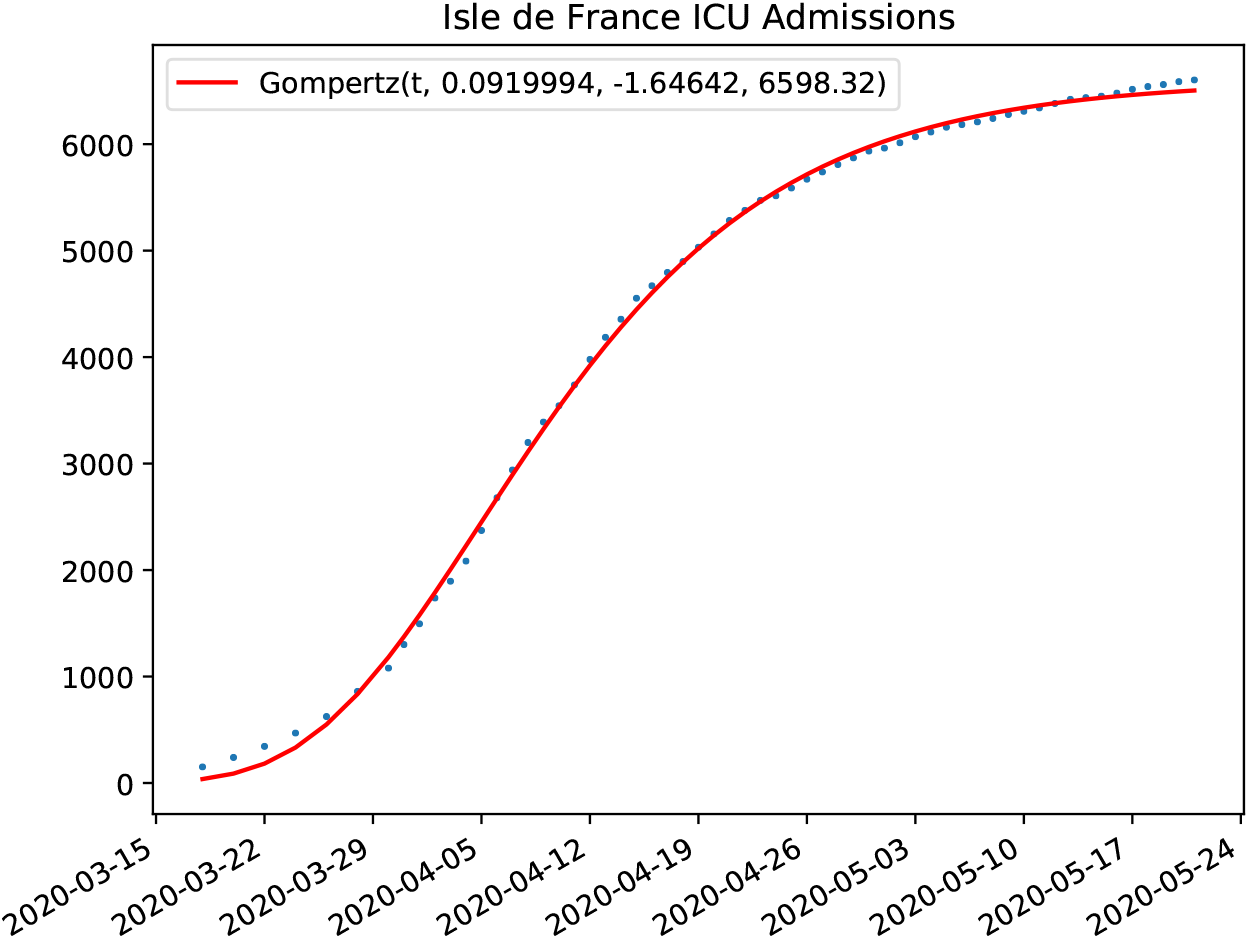
Isle de France Covid-19 ICU Admissions, 18 Mar to 21 May 2020

**Figure 29:**
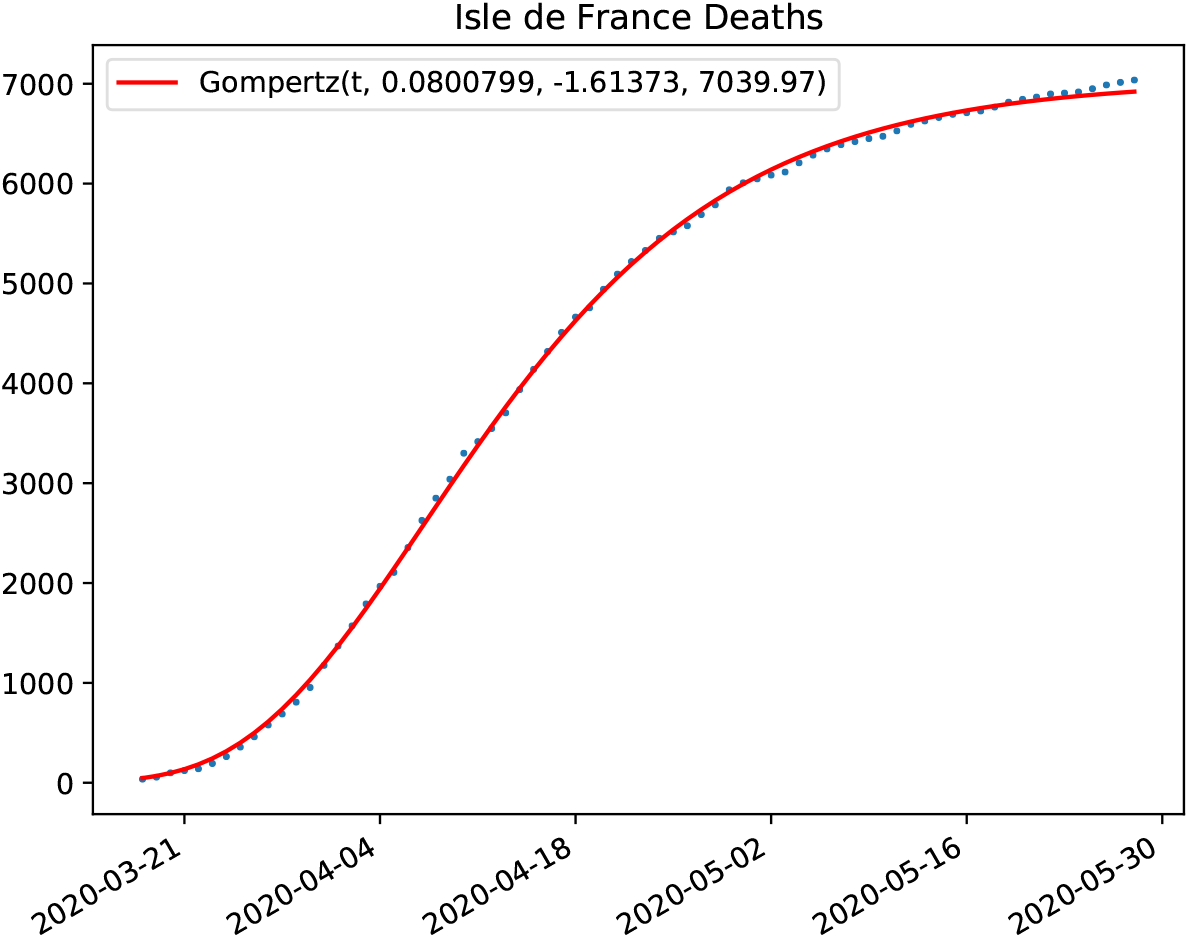
Isle de France Covid-19 Deaths 18 Mar to 31 May 2020

**Figure 30:**
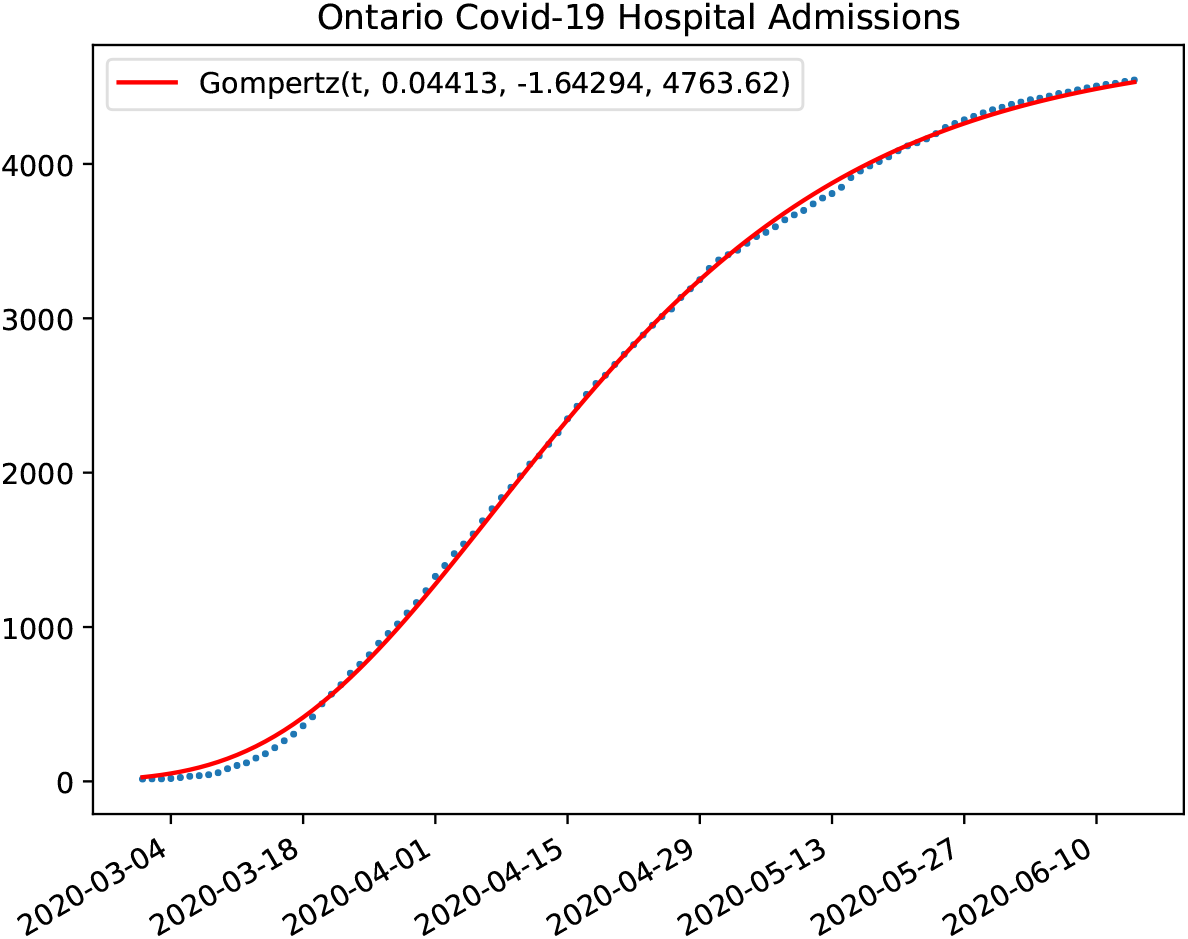
Hospital admissions for Covid-19 in Ontario 1 Mar to 14 Jun 2020

**Figure 31:**
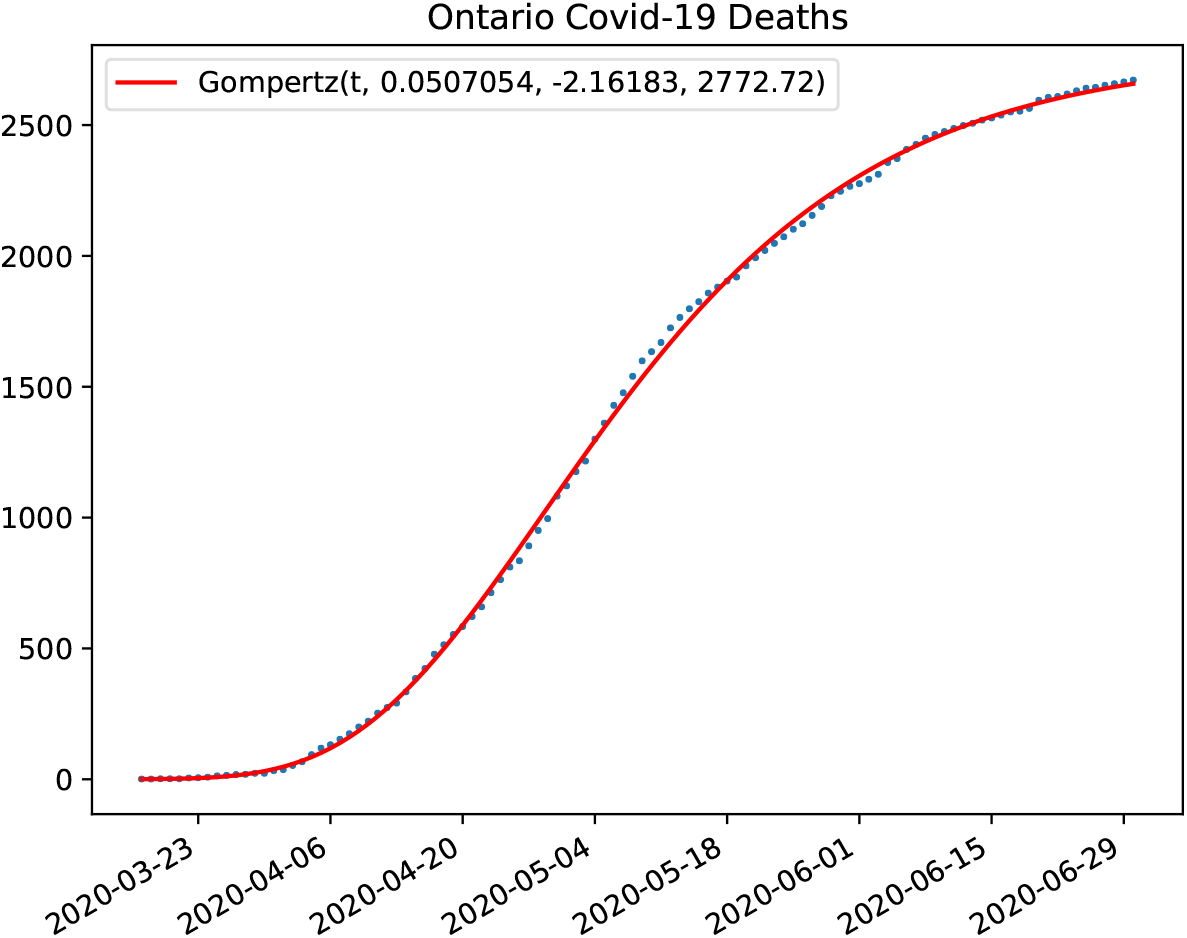
Covid-19 Deaths in Ontario from 17 Mar to 30 Jun 2020

**Figure 32:**
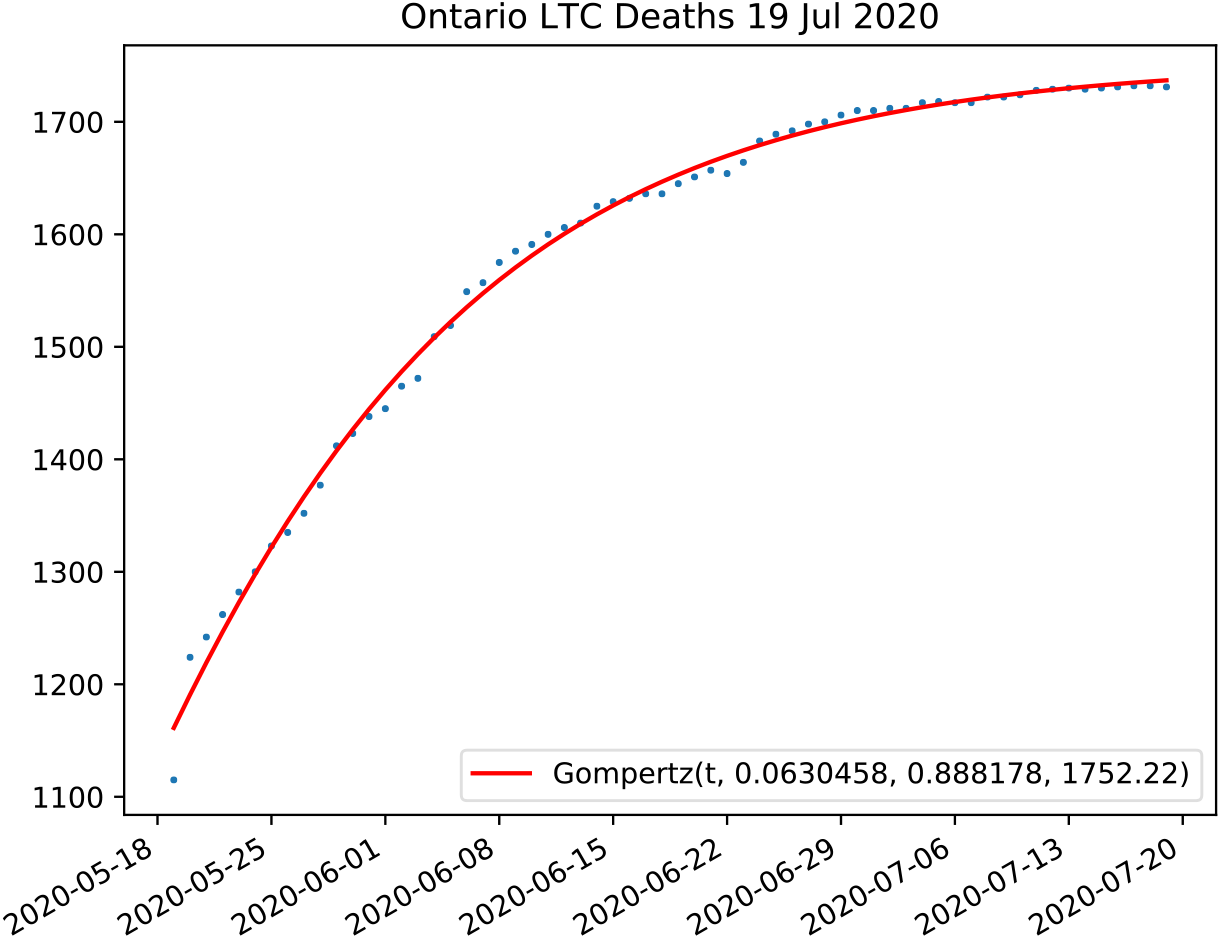
Long Term Care Deaths Ontario from 19 May to 19 Jul 2020

**Figure 33:**
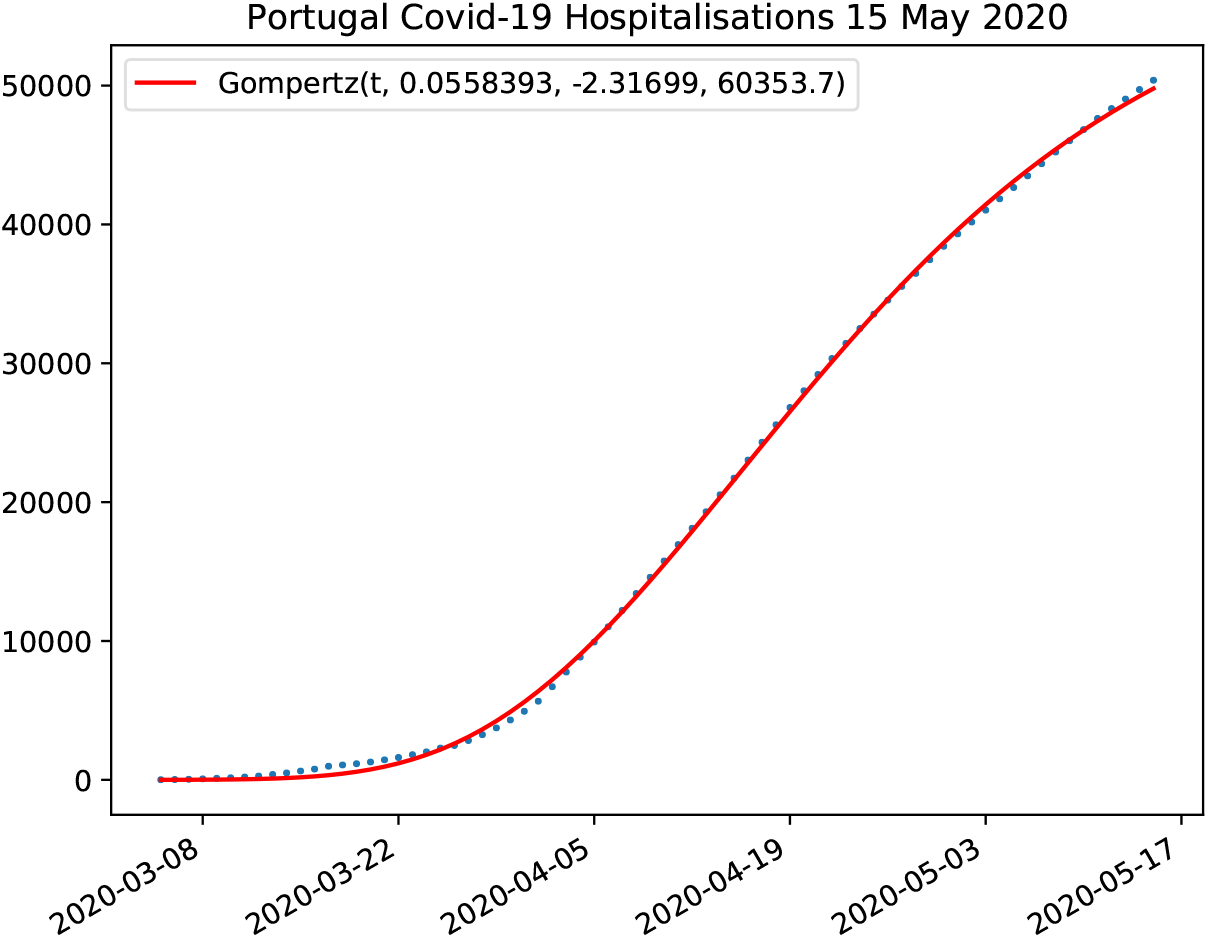
Covid-19 Hospitalisations in Portugal from 5 Mar to 15 May 2020.

**Figure 34:**
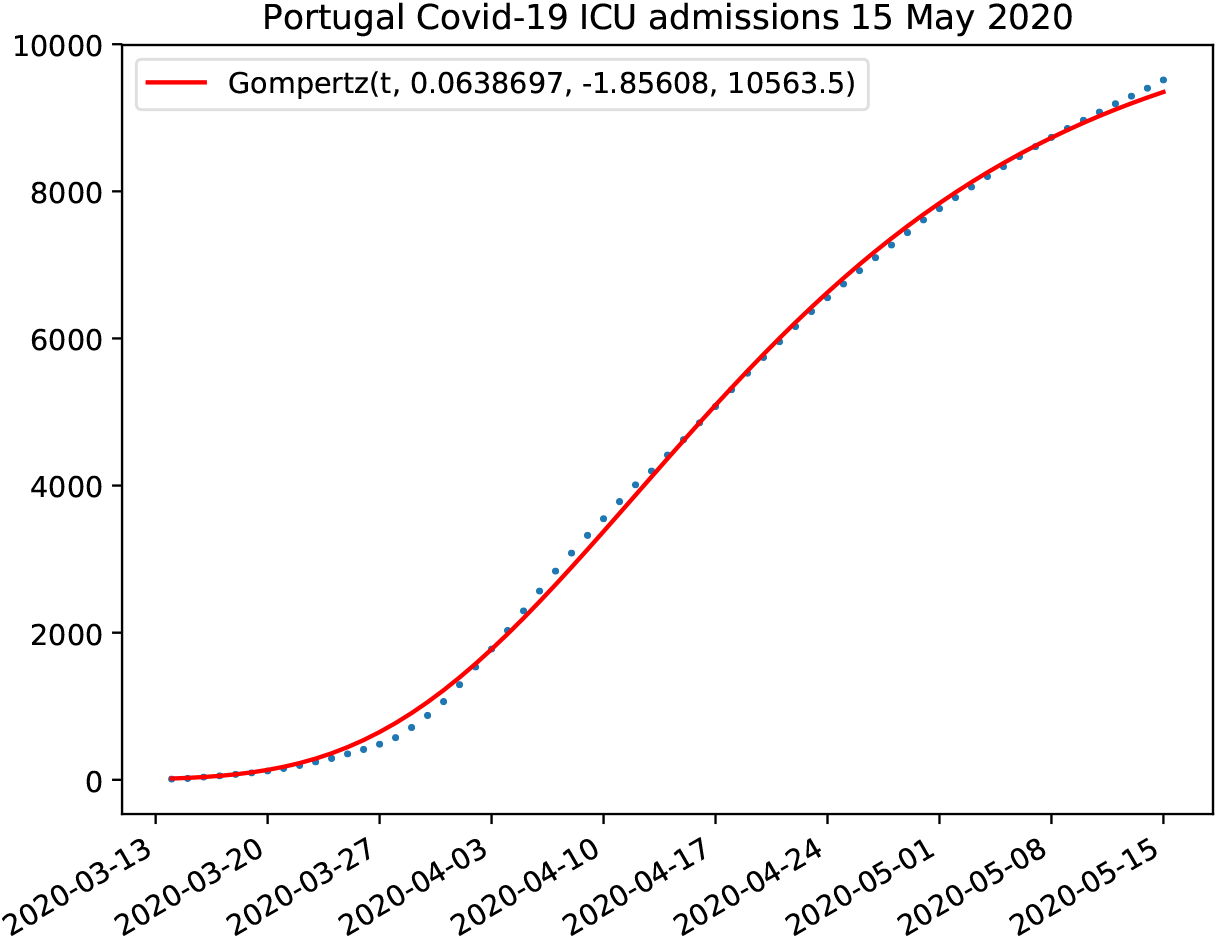
Covid-19 ICU admissions in Portugal from 14 Mar to 15 May 2020.

**Figure 35:**
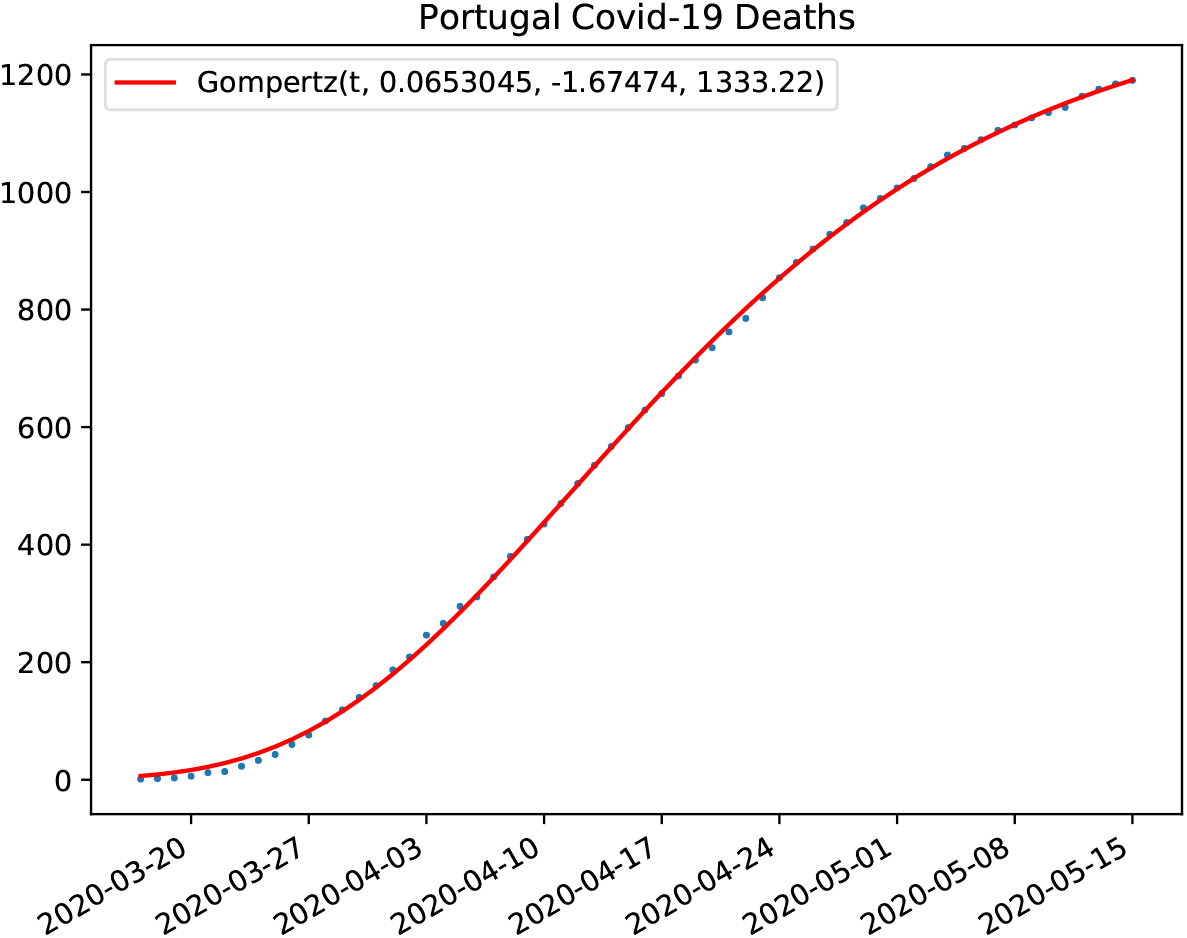
Covid-19 Deaths in Portugal from 17 Mar to 15 May 2020.

**Figure 36:**
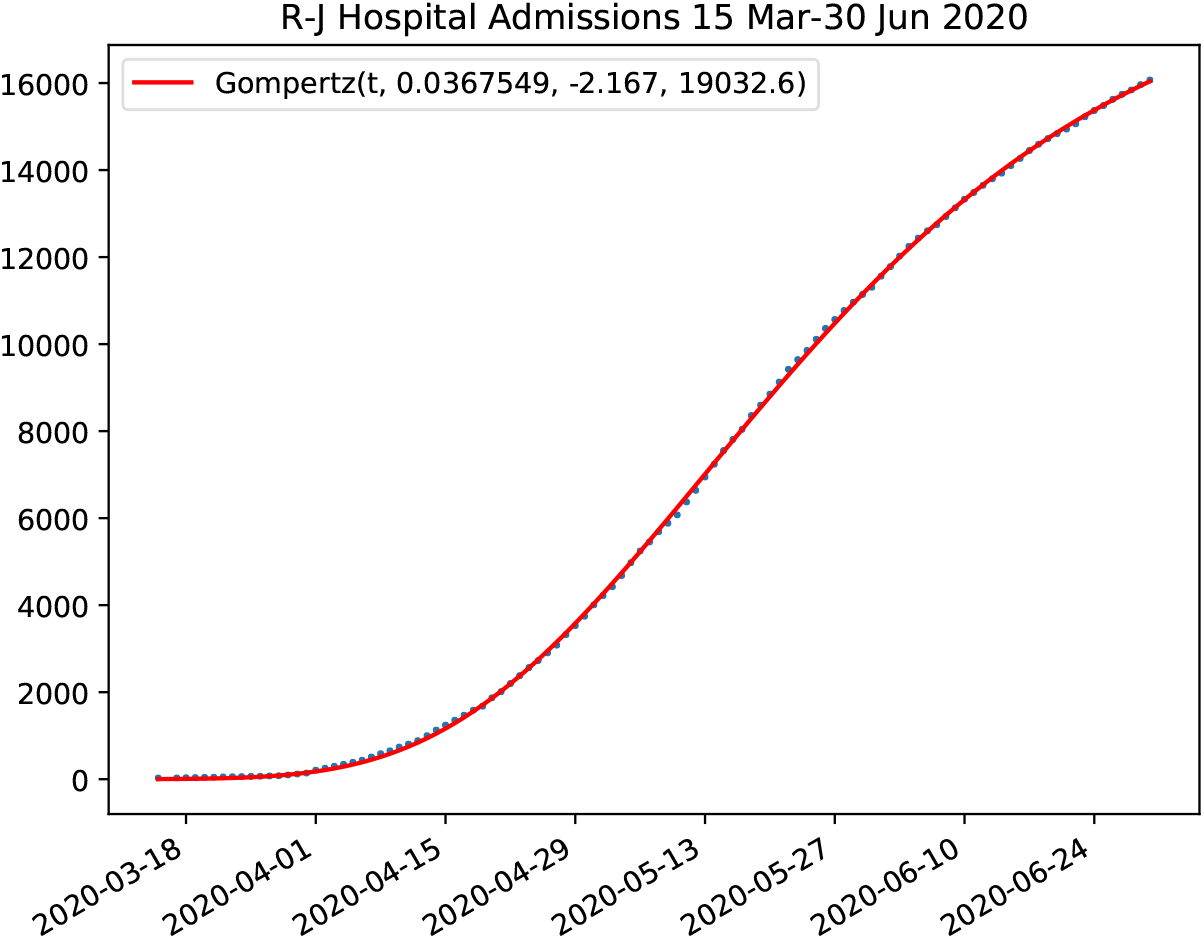
Covid-19 Hospitalisations in RJ from 15 Mar to 30 Jun 2020.

**Figure 37:**
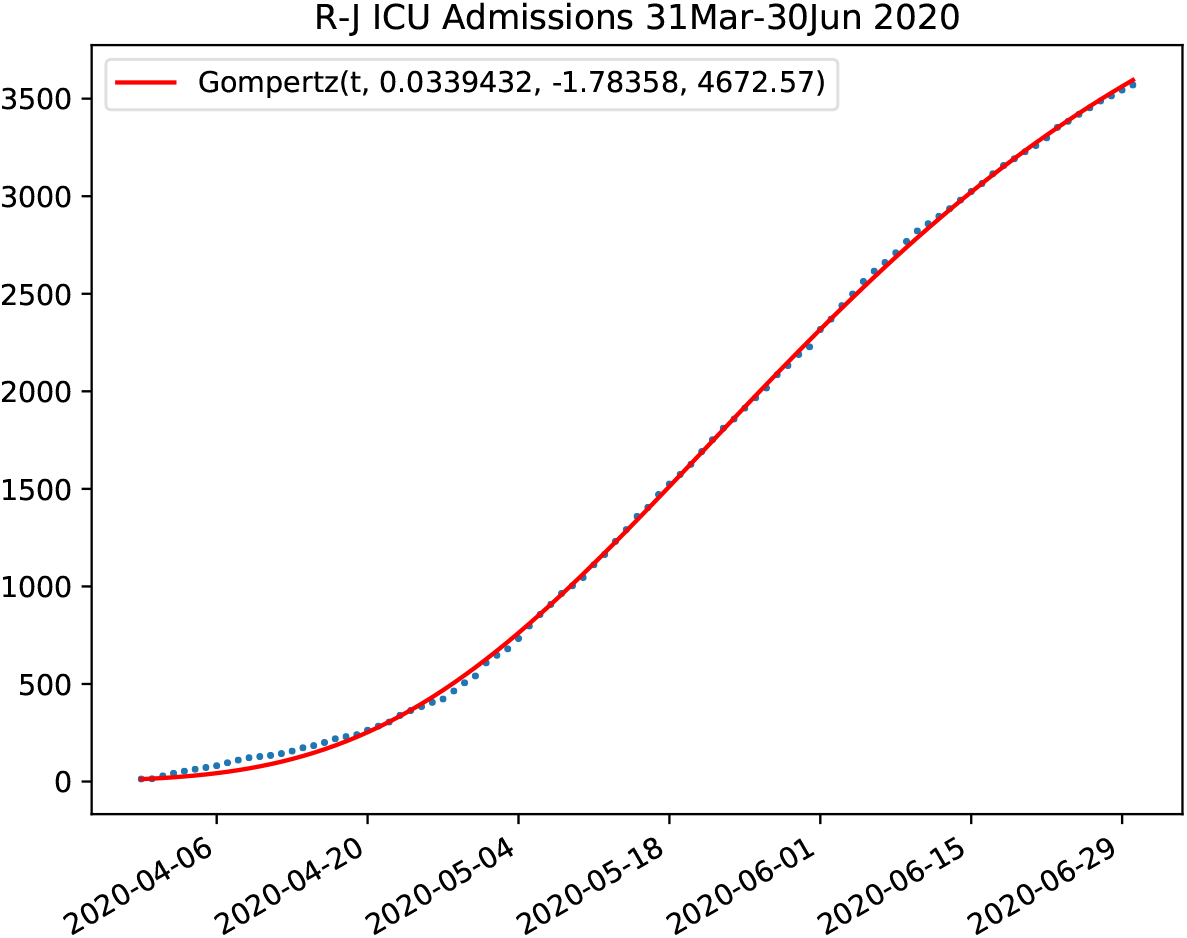
Covid-19 ICU admissions in RJ from 31 Mar to 30 Jun 2020.

**Figure 38:**
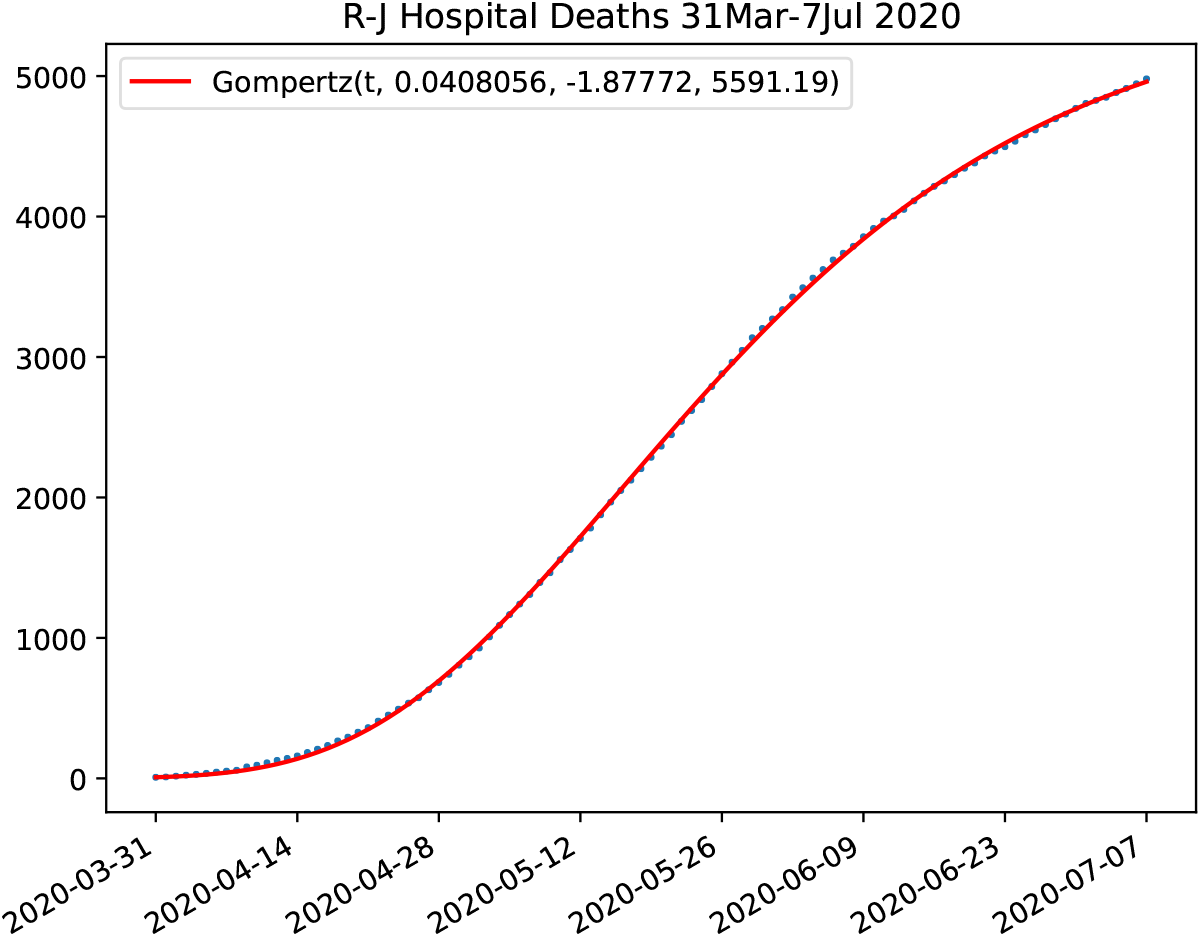
Covid-19 Hospital Deaths in RJ from 31 Mar to 7 Jul 2020.

**Figure 39:**
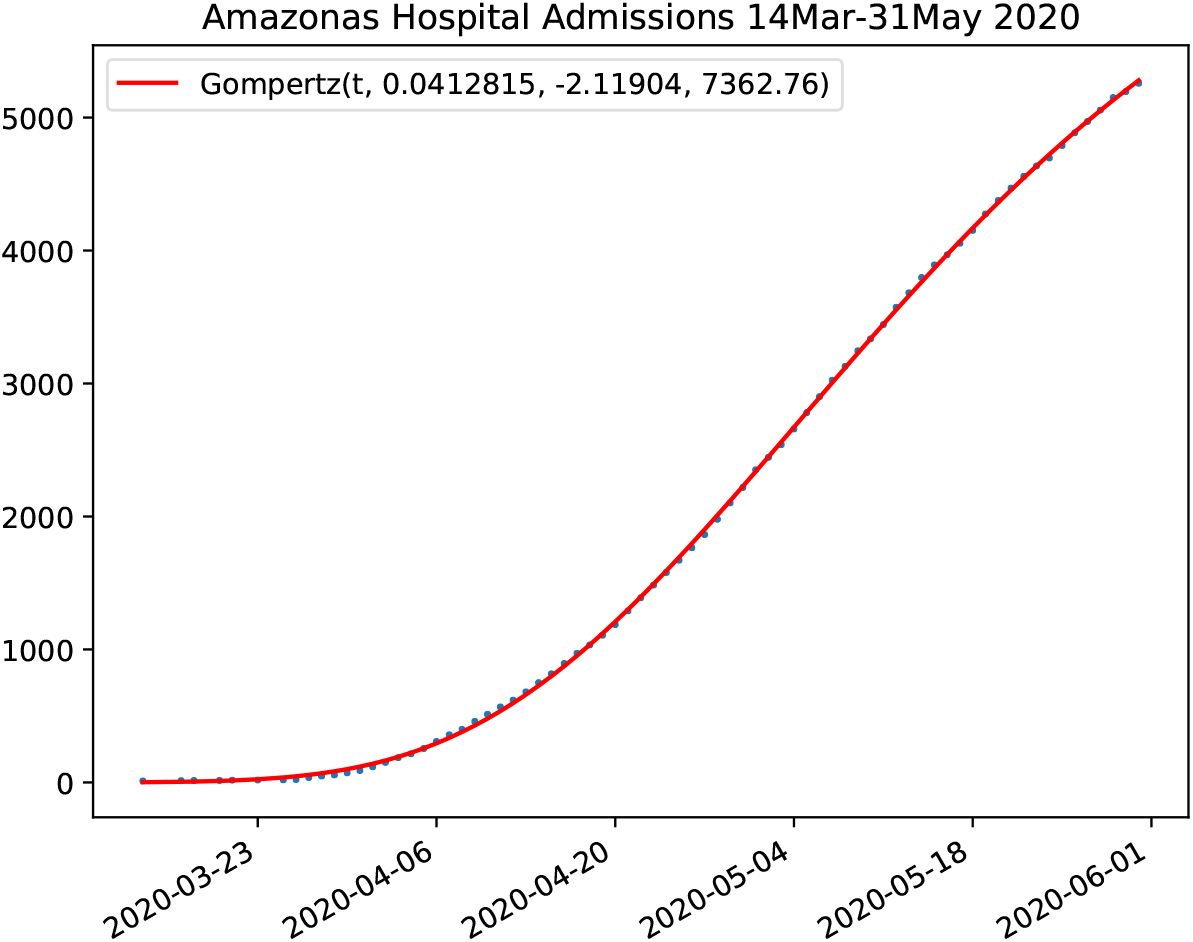
Covid-19 Hospitalisations in Amazonas from 14 Mar to 31 May 2020.

**Figure 40:**
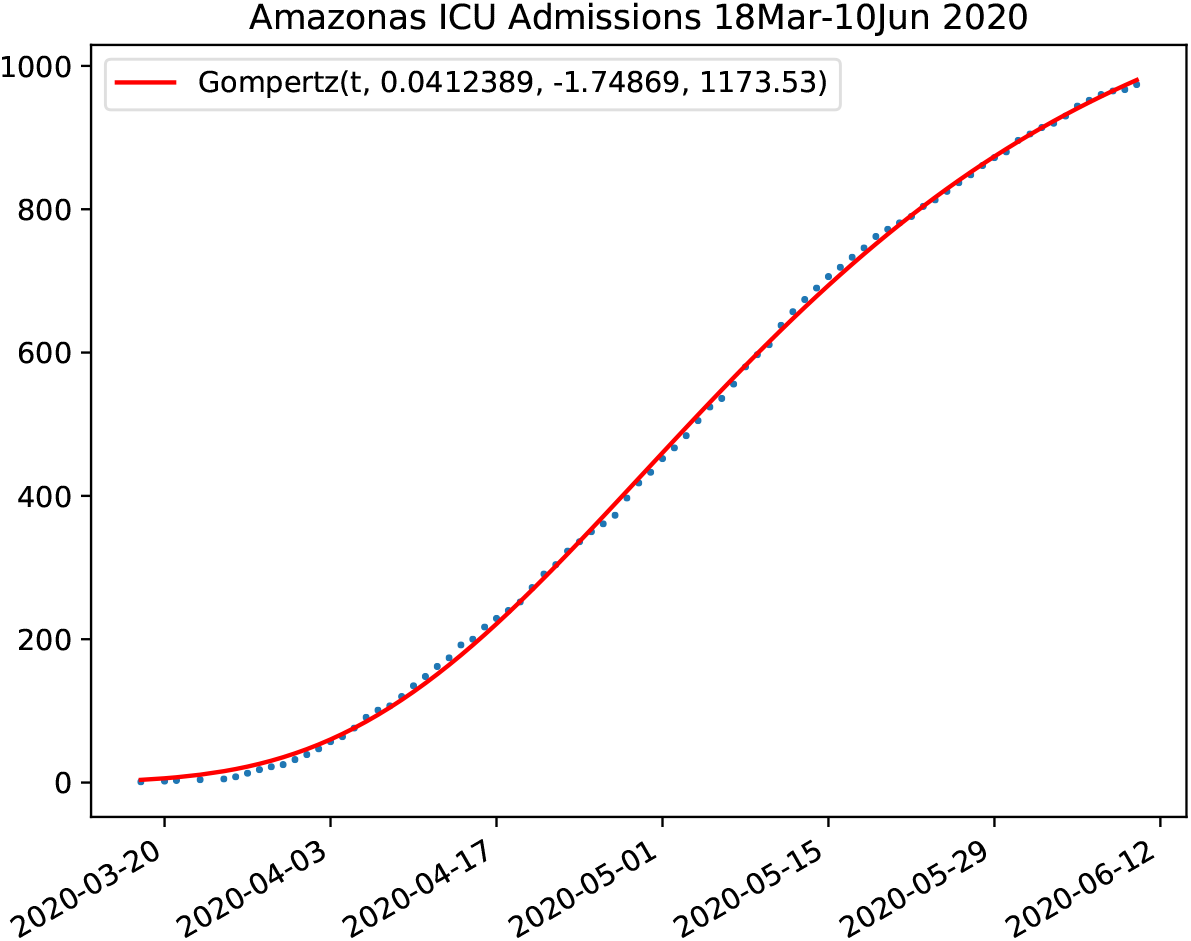
Covid-19 ICU admissions in Amazonas from 18 Mar to 10 Jun 2020.

**Figure 41:**
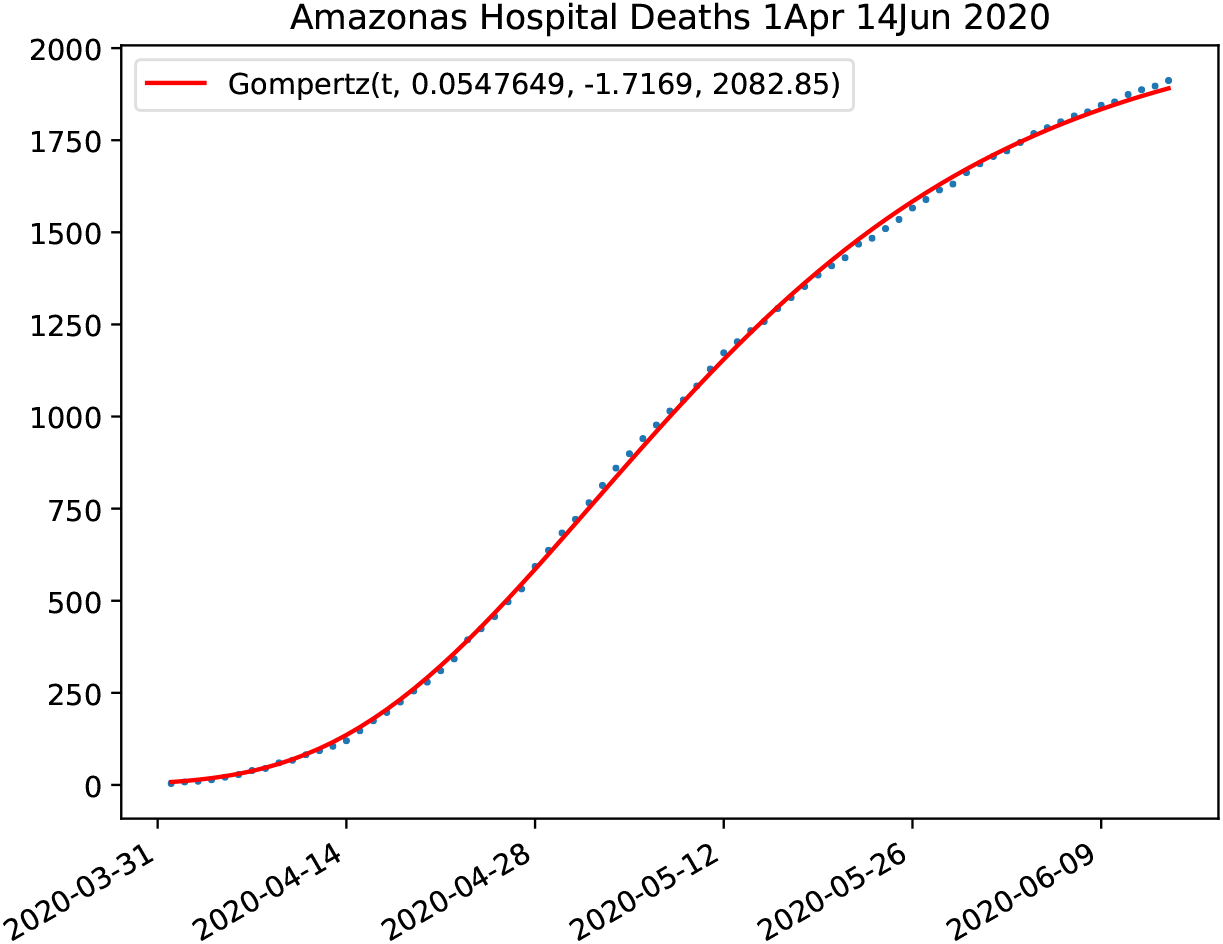
Covid-19 Hospital Deaths in Amazonas from 1 Apr to 14 Jun 2020.

We note that, in the tests of predictive power we have presented, no attempt has been made to speed up the convergence to the ultimate model parameters. Figures 42 to 48 show examples of out of sample errors in for Gompertz Function fits to hospital data.

**Figure 42:**
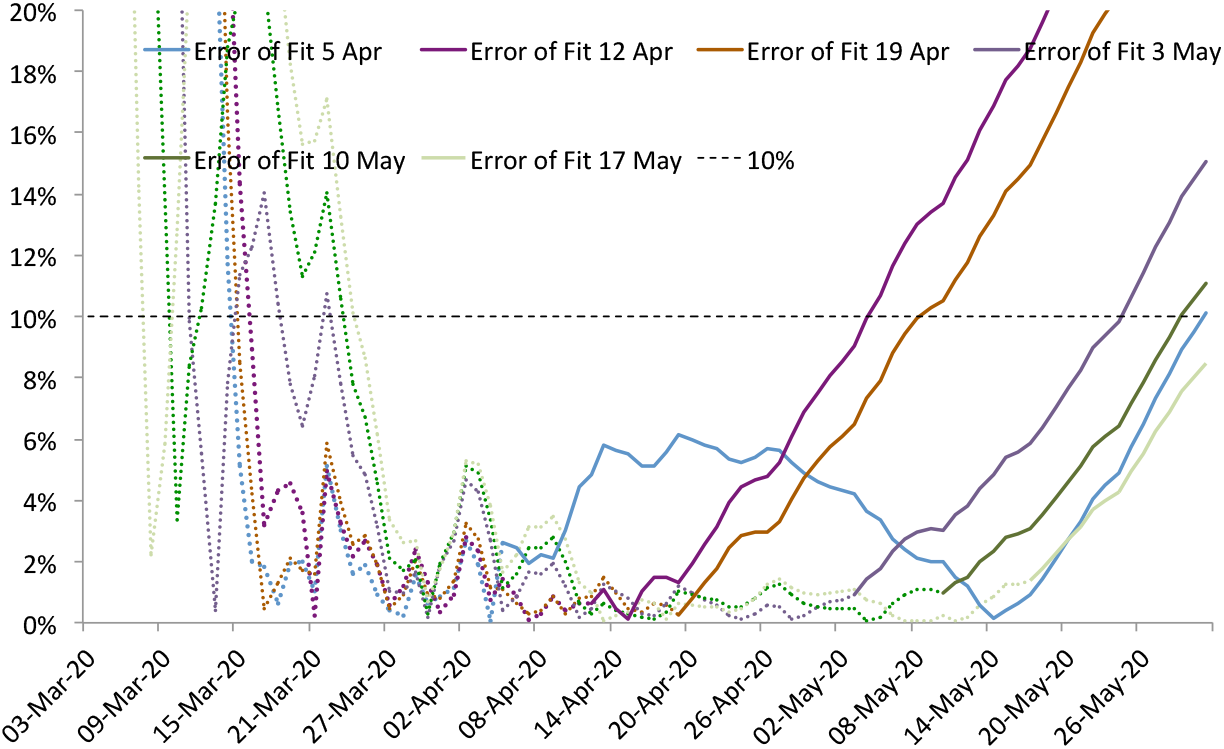
Evolution of errors in Gompertz Function fits to Swedish Hospital Admissions.

**Figure 43:**
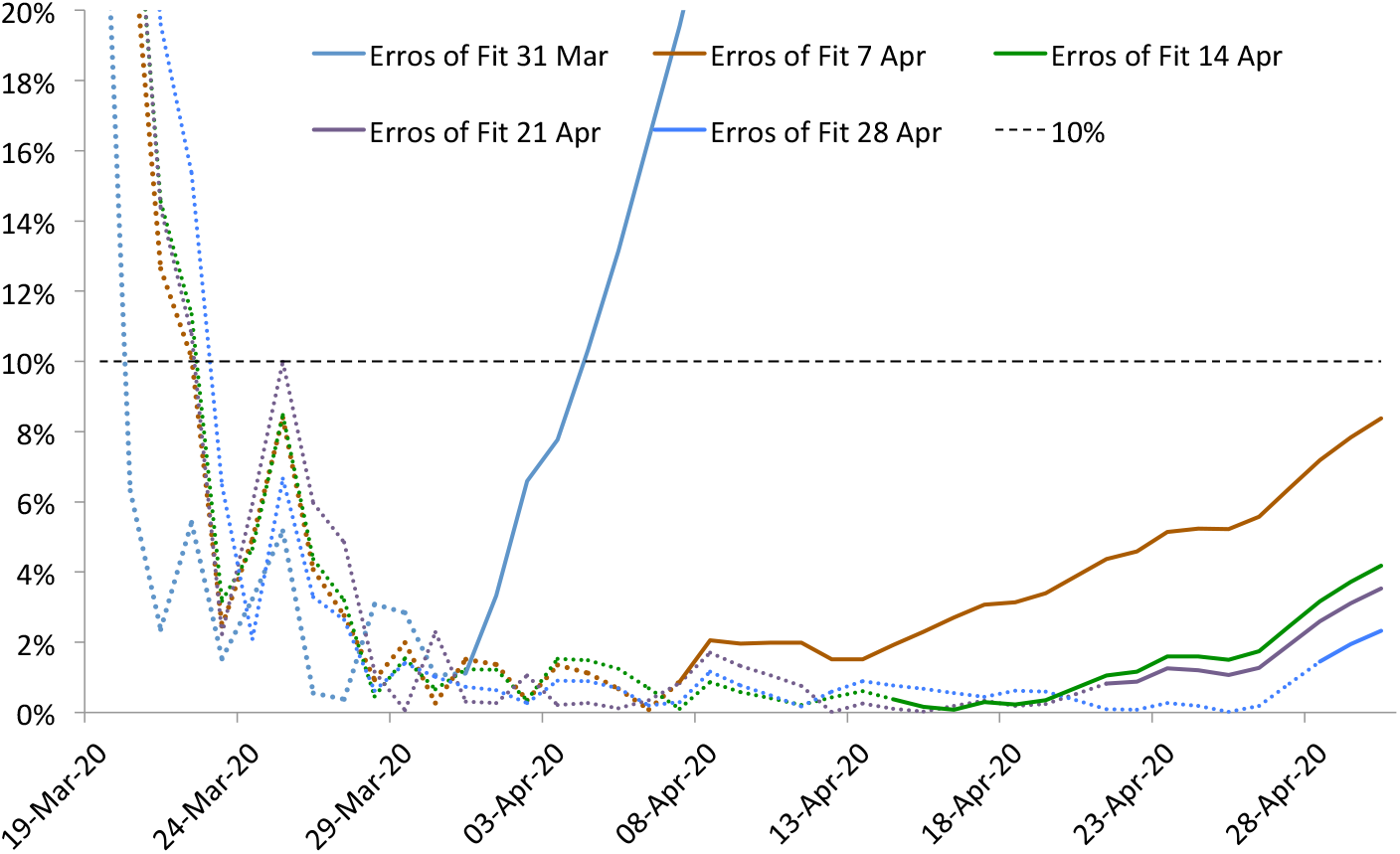
Evolution of errors in Gompertz Function fits to London Hospital Admissions.

**Figure 44:**
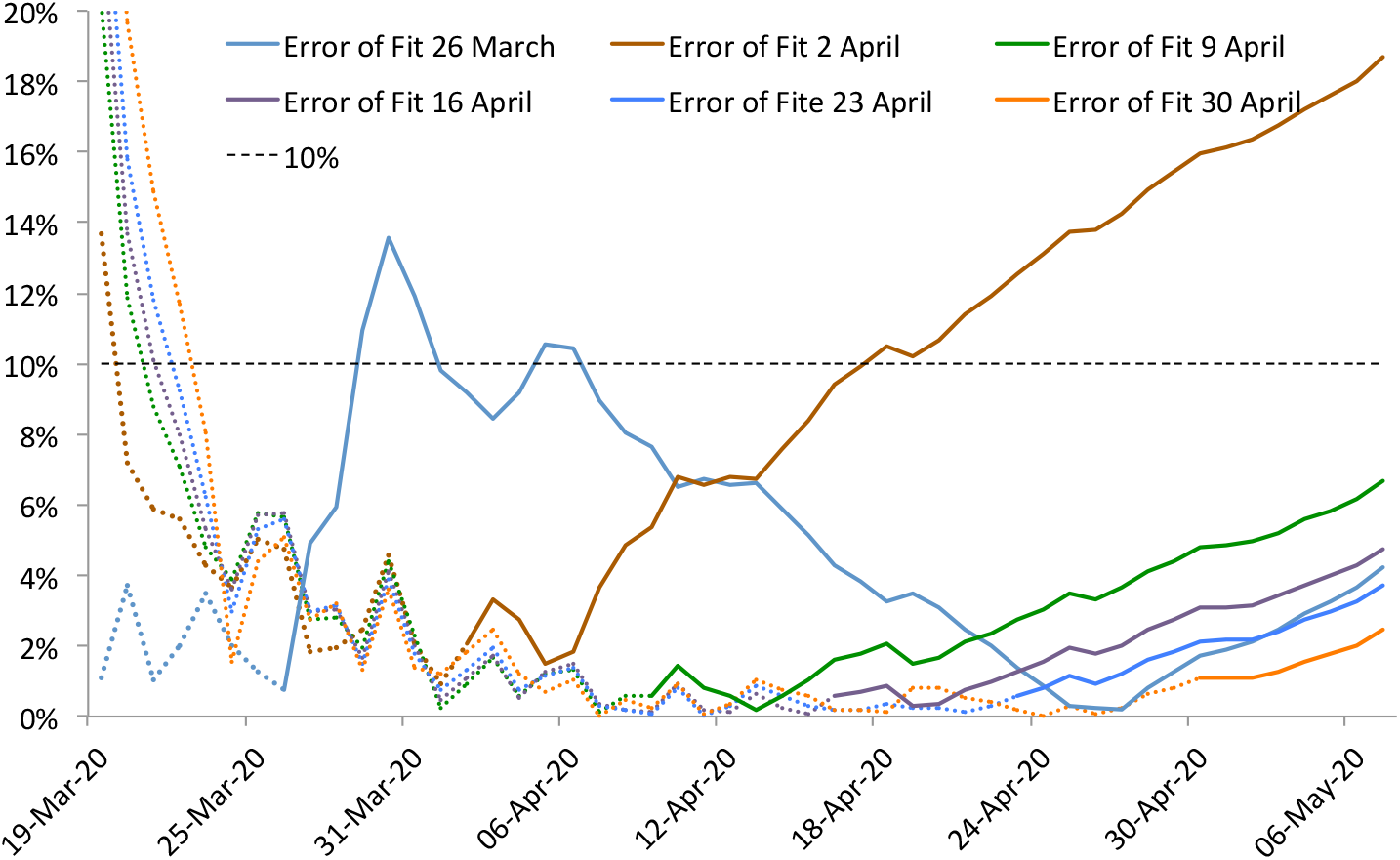
Evolution of errors in Gompertz Function fits to Isle de France ICU Admissions.

**Figure 45:**
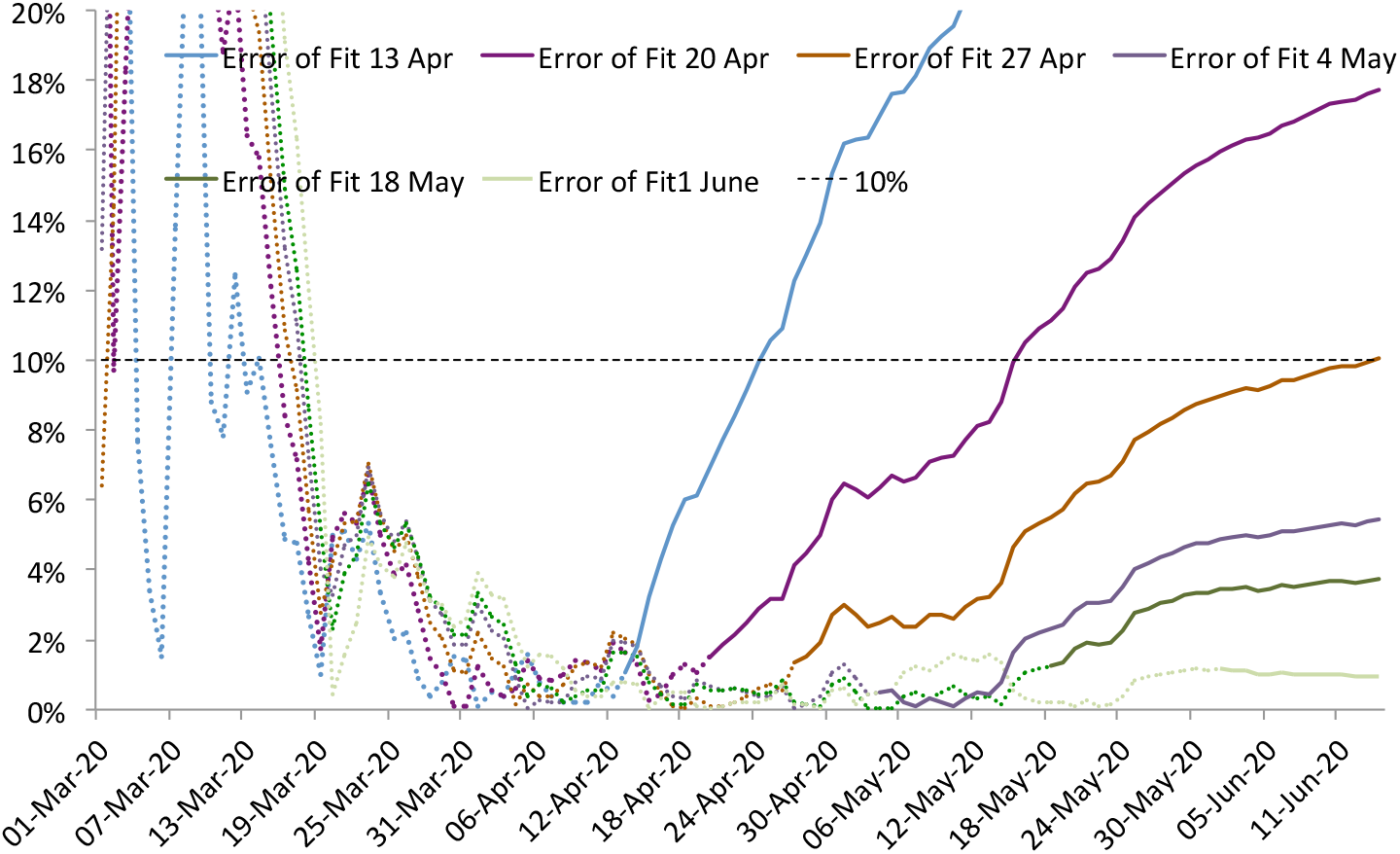
Evolution of errors in Gompertz Function fits to Ontario Hospital Admissions.

**Figure 46:**
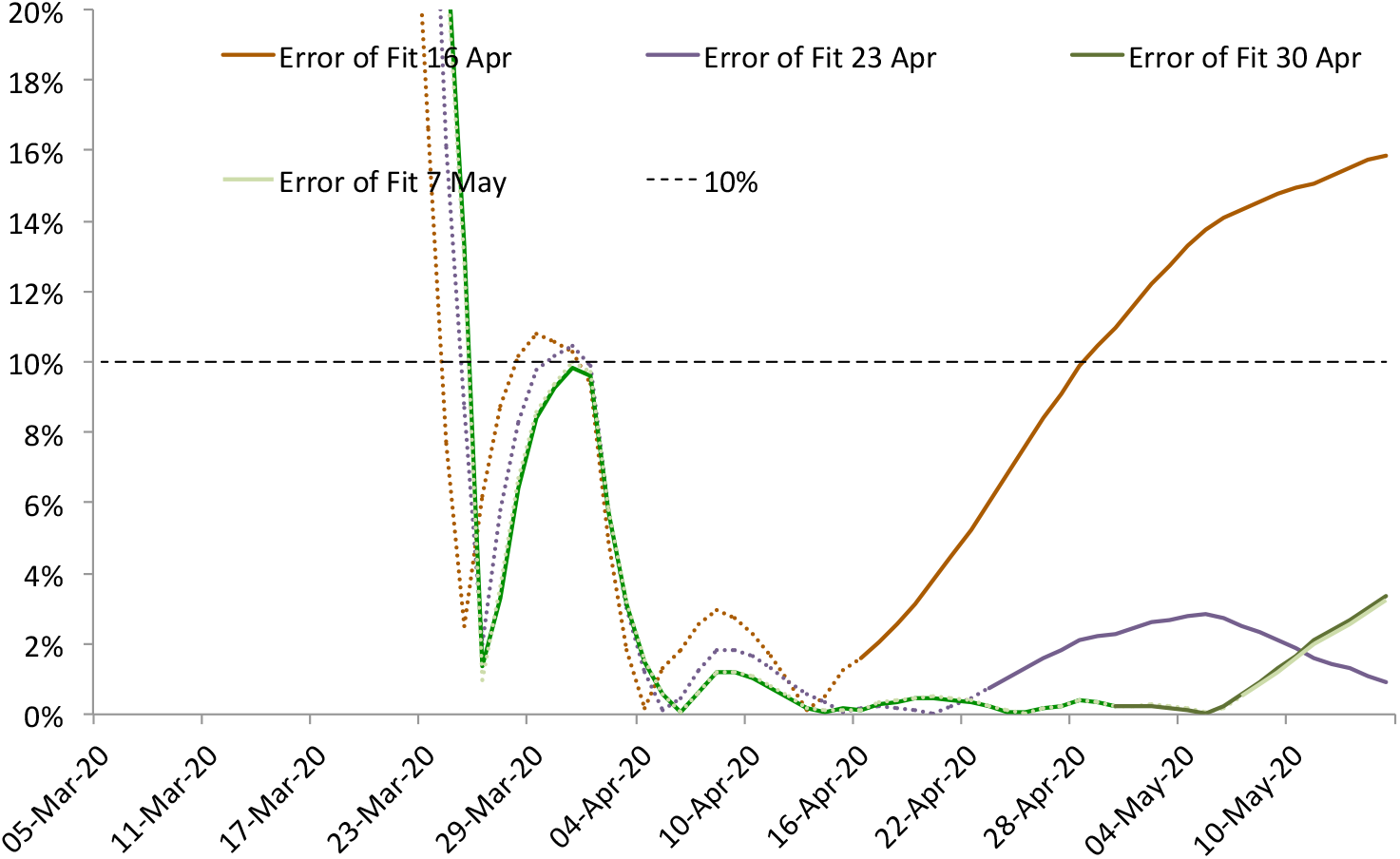
Evolution of errors in Gompertz Function fits to Portuguese Hospital Admissions.

**Figure 47:**
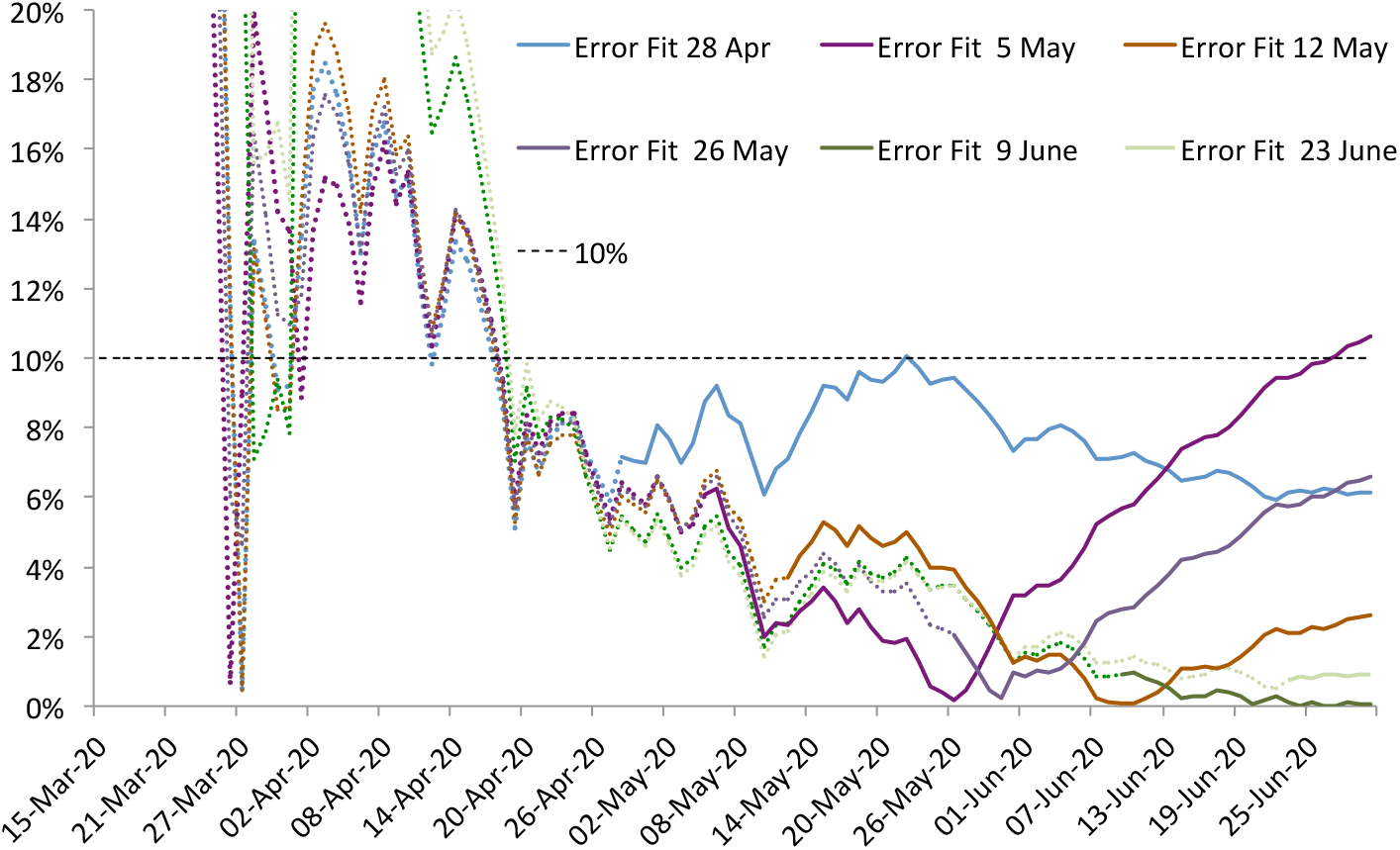
Evolution of errors in Gompertz Function fits to RJ Hospital Admissions.

**Figure 48:**
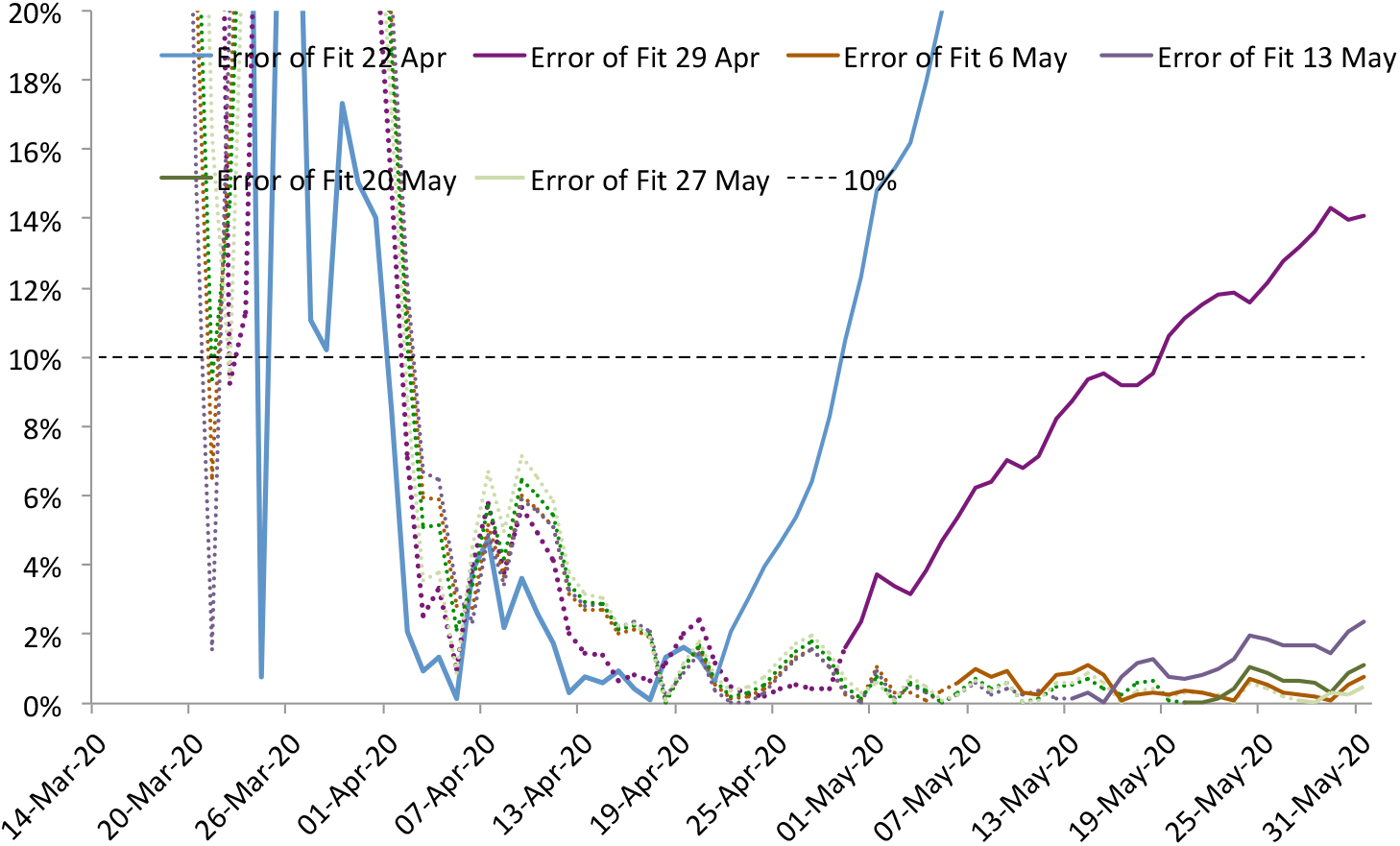
Evolution of errors in Gompertz Function fits to Amazonas Hospital Admissions.

These examples show that, as expected, the good fits observed in Figures 20 to 41 mean that as the epidemic progressed, the errors in the out of sample predictions of the Gompertz Function model improved to remain below the 10% level we specified.

This convergence is close to monotone for periods long enough to be practical for use in planning for health infrastructure demands. Knowledge that realistic short to medium term estimates of these demands can be made is extremely important given the universal fear of overwhelming healthcare systems that the Covid-19 epidemic has generated.

While the errors always converge to a level below 10%, the time this takes varies significantly across our examples. The errors at the beginning of the in-sample data also remain large even as they converge to very low levels out of sample as the epidemic progresses. In order to discuss these and other regularities and to examine the the extension of the Gompertz Function Model to subsequent Covid-19 outbreaks, we will make use of some observations about the Gompertz Function dynamics to which we devote the next section.

## 3 Gompertz Function Dynamics

### 3.1 The Gompertz Function and the Gumbel Distribution

If *X*(*t*) is the cumulative number of cases at day *t* and *N* is the final (as yet unknown) level, then, writing 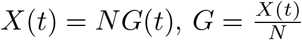 is the fractional number of cases at time *t*.

Therefore *G* is a non-decreasing function whose values lie in [0, 1]–a probability distribution.^13^ The distribution *G*(*t, a, b*), has (observable) scale and location parameters *a* and *b* which vary over time but converge as the epidemic tends to its conclusion.

For the Gompertz Function, *G* is the *Gumbel distribution*

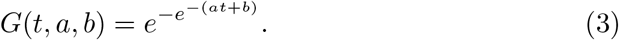

The constant *N* determines the final *scale* of the epidemic, but the dynamical properties of the Gompertz function are completely determined by the Gumbel distribution.^14^

In addition to the daily rate of change of *X*, denoted by 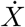, a critical quantity for the impact on a population is the *relative* rate of change 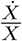– the new infections or deaths each day as a fraction of the total population affected so far.

Since *N* is constant, 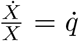 where *q* = *log*(*G*(*t, a, b*)). It’s immediate from the definition of the Gumbel distribution that

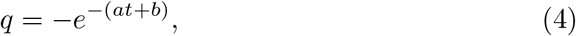

and

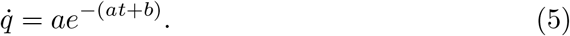

It follows from the equation for 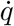 that, unlike the case of exponential growth where the relative rate of change is constant, in Gompertz Function growth the relative rate of change of *X* is *decreasing exponentially* as time increases.

Using the first difference approximation to 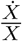, the derivative of *log*(*X*), we have 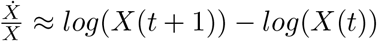.

Since 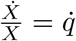 this means that *log*(*log*(*X*(*t* + 1)) −*log*(*X*(*t*))) ≈ *log*(*a*) − *b*− *at*, a straight line with slope −*a* and intercept *log*(*a*) −*b*.

We can observe *a*_*i*_ and *b*_*i*_ from the slope and intercept of the line at time *t*_*i*_. Of course in practice the plot of *log*(*log*(*X*(*t*_*i*+1_)) − *log*(*X*(*t*_*i*_))) is only approximately a straight line and the observations of the parameters will be volatile, especially early in the epidemic. Nevertheless, it’s easy to see in epidemic data that, as time increases, this series trends downward steadily and tends to a straight line with negative slope.(See Figure 49)

**Figure 49:**
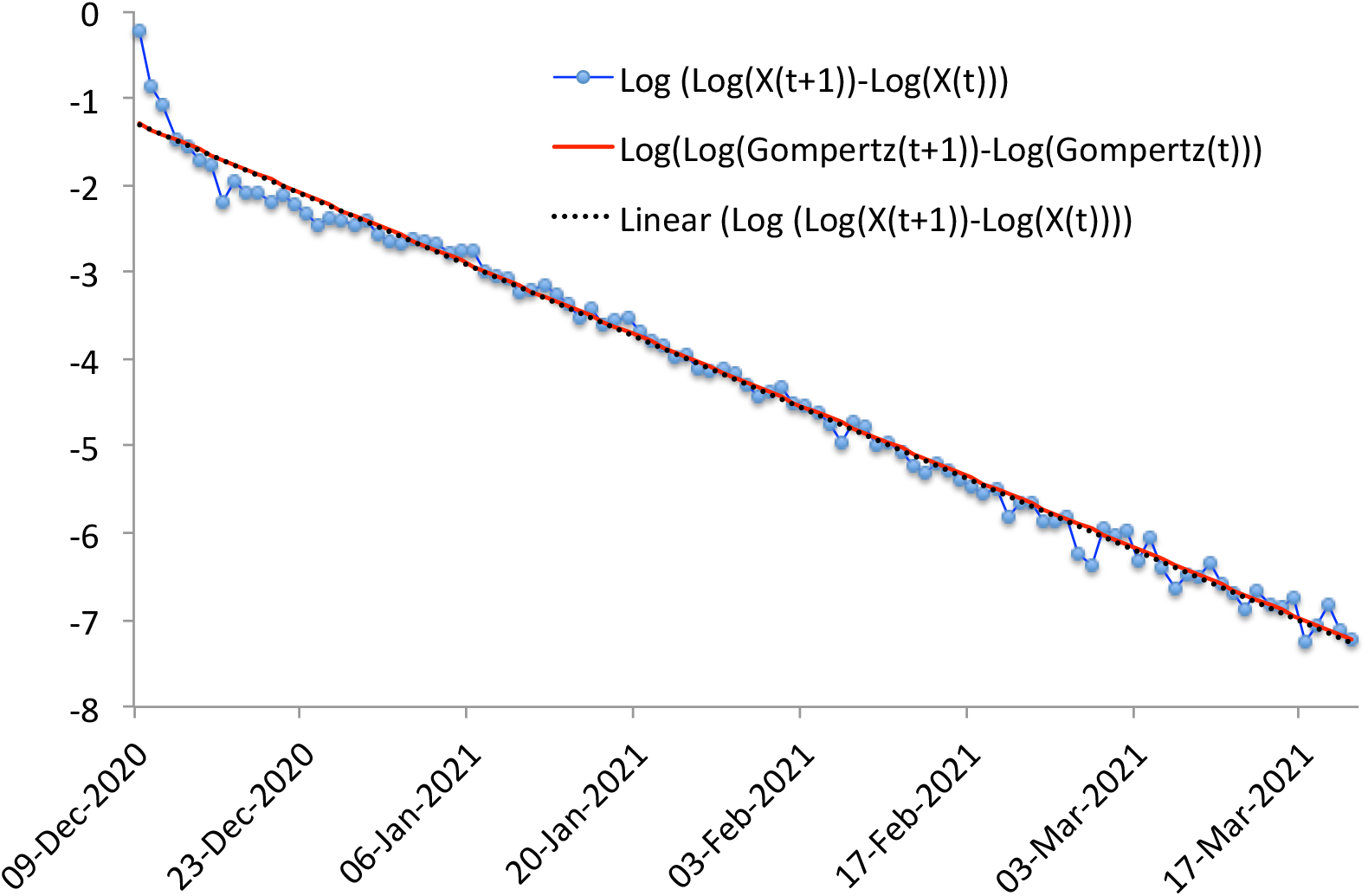
Covid-19 Hospital Admissions in London and the Gompertz Function Fit 8 Dec 2020 to 21 Mar 2021.

While the Gompertz function never increases exponentially, it does grow rapidly at first with both its velocity 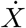 and its acceleration 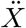 increasing.^15^The parameters *aN* and *a*^2^*N* control these rates of growth. Note that *a* has units of *t*^−1^ so that *aN* and *a*^2^*N* have the correct units for the velocity and acceleration of *X*.

### 3.2 The Scaled, Time-shifted Version of a Gompertz Function

Suppose that the daily increments for one process *Y* (for example ICU admissions) is a multiple *λ* of another process *X* (for example Hospital Admissions) but with a shift of *l* days. If *X* is a Gompertz function then

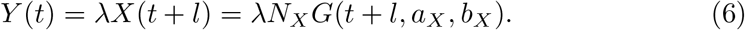

It follows that *Y* is a Gompertz function whose parameters are

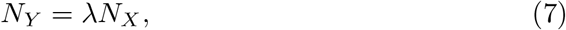

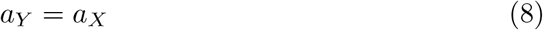

and

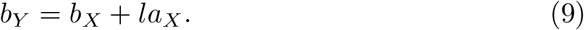

These relations hold approximately among some of the Gompertz function groups for hospital admissions, ICU admissions and deaths presented in the previous section. For example, although the hospital and ICU admissions data for Isle de France both begin on the same day, Equation 9 says that ICU process lags hospitalisations by about 4.2 days. This is also the lag between the maxima of their Gompertz function derivatives. For this case the values of *N* in ICU and Hospital admissions predict that 17% of those hospitalised will be admitted to ICU. At the end of the data sample a comparison of ICU admissions with Hospital admissions 4 days earlier gives a ratio of 17.8%.

### 3.3 Critical Times and a Natural Time Scale

The Gumbel distribution’s features produce a number of characteristic times for Gompertz Function growth.

We denote by *T*_1_(*a, b*), the time at which the acceleration 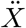 reaches its peak. It’s easy to check by finding the zeros of the third *t*-derivative of *X*, that *T*_1_ is given by

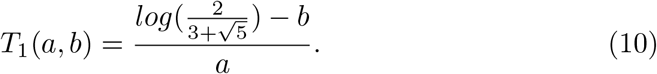

At time *T*_1_, 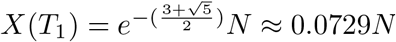.

The velocity 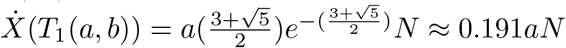.

The peak acceleration 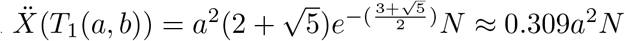.

Thus, by the time about 7.3% of the susceptible population *N* has been infected, the acceleration has reached its peak of approximately 0.309*a*^2^*N*, and begins to decline quickly.

The velocity which is approximately 0.191*aN* at *T*_1_(*a, b*) continues to increase until time *T*_2_(*a, b*)

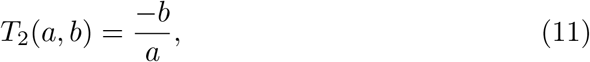

when 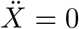.

The parameter *a* is the value of 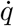 at *T*_2_. The dimensionless parameter *b* determines the fraction of the final level *N* observed on day 0:

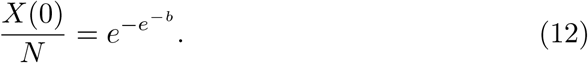

At time 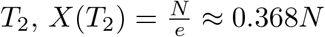 so the velocity 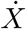 begins to decline once about 37% of the susceptible population is infected. It is easy to verify that the peak velocity 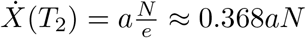 –so between peak acceleration and peak velocity the velocity has almost doubled: 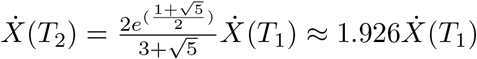.

The acceleration 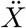 remains negative for all time *t* > *T*_2_ and reaches its minimum at time *T*_3_(*a, b*) given by

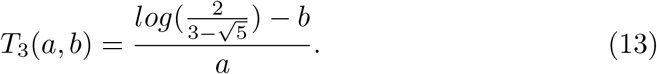

At time 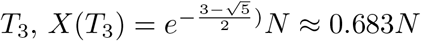 and about 68% of the susceptible population is infected.

The velocity 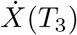 has fallen to 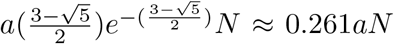 or about 71% of its peak value.

A natural time scale is 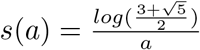.

*T*_2_ is midway between *T*_1_ and *T*_3_ and *s*(*a*) is the common distance between them. The larger the value of *a* the shorter the interval between the critical times.

As an indication of how quickly 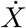 is growing in the early phase of the epidemic, between time *T*_1_ − *s*(*a*), where 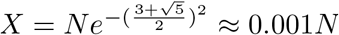, and time *T*_1_, 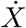 increases by a factor of more than 26.

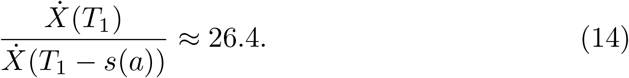

Likewise, 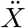 increases by a factor of approximately 7.3 in the same period. But even over this period of rapid increase, the growth is *never* exponential. In fact the third derivative of *X* decreases steadily to 0 over the interval from 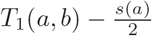 to *T*_1_(*a, b*), then remains negative until *T*_3_(*a, b*).

The decline in 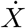 after it reaches its peak is slower than its initial growth producing the characteristic asymmetry seen in Figure 50.

**Figure 50:**
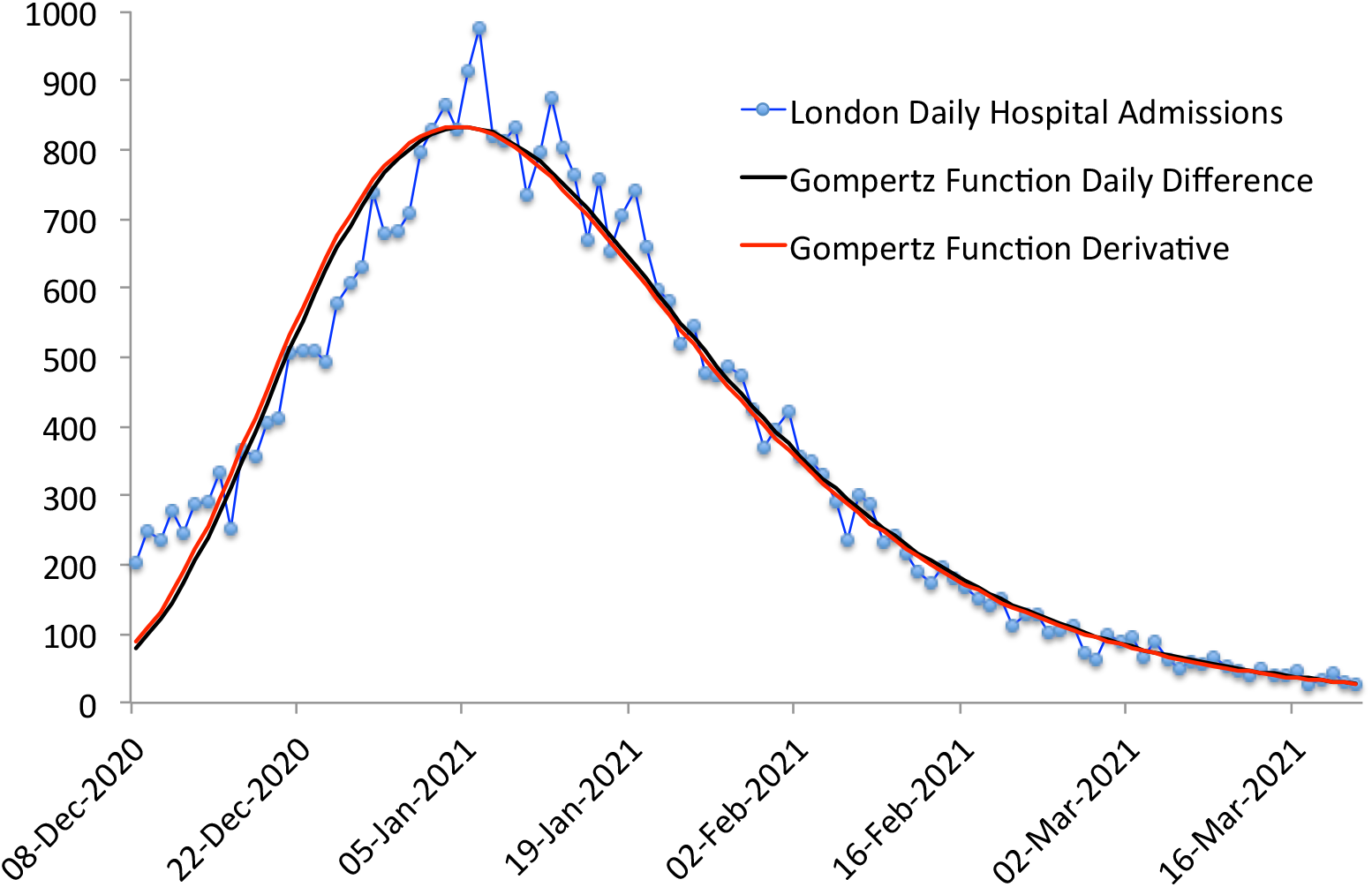
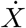 and Daily Differences for London Hospital Admissions 8 Dec 2020 to 21 Mar 2021 show the characteristic rapid rise and slow decline in 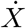.

It is not until time 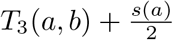 that 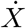 drops back to about the value it had at *T*_1_. 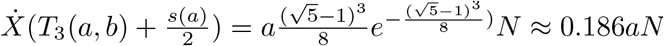.

### 3.4 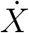, Daily Increments and Asymmetry

The Gompertz Function Model fits the cumulative observations, so by taking the differences *X*(*t*_*i*+1_) −*X*(*t*_*i*_) we have a model for the daily observations. The Gompertz Function time derivative 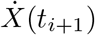 is a very good approximation to this first difference as can be seen in Figure 50.

The maximum and minimum of the acceleration are located symmetrically around the peak of the Gompertz function velocity, but as we have seen before, 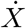 itself is asymmetric–as Figure 50 shows. For a symmetric ‘S-curve’ peak velocity comes at the point where 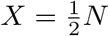 while for the Gompertz Function it is 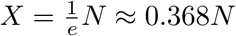.

As a result, any growth function *Y* for which 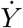 is symmetric will underestimate the asymptotic level *N* for any epidemic following a Gompertz Function. For example, when the curvature changes sign for a Logistic function at time 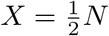 so we get *N* = 2*X*(*T*) instead of *eX*(*T*)–an underestimate of *N*.^16^

### 3.5 Epidemic Duration

The natural timescale *s*(*a*) can be used to estimate the time remaining in the epidemic if it continues to follow the Gompertz Function.^17^

As soon as we have stable values for the parameters *a, b* and *N*, we can calculate the remaining time, in days, to reach any desired fraction of *N*. These times are conveniently described in terms of *T*_2_ and *s*(*a*).

For example, we have already observed that *X*(*T*_3_) = *X*(*T*_2_(*a, b*) + *s*(*a*)) ≈ 0.68*N*. We also have *X*(*T*_2_(*a, b*) + 2*s*(*a*)) ≈ 0.84*N, X*(*T*_2_(*a, b*) + 3*s*(*a*)) ≈ 0.95*N* and *X*(*T*_2_(*a, b*) + 4*s*(*a*)) ≈ 0.99*N*.

### 3.6 The Reproduction Number and the Herd Immunity Threshold

The effective Reproduction Number *R*_*t*_ for an epidemic at time *t* is defined as the average number of secondary infections that are produced by each primary infection.[5]There is generally no way to observe infections and this number must be inferred from other quantities–such as cases–so the value of *R*_*t*_ is typically subject to considerable uncertainty.

Nevertheless, we can draw some conclusions about *R*_*t*_ for an epidemic which is following a Gompertz Function, at least if the growth of cases is a reasonable proxy for the growth of infections.

For any time *t* > *T*_2_, the acceleration 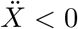, so 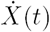 is strictly decreasing. Because the derivative at time *t* + 1 is almost identical to the difference *X*(*t* + 1) −*X*(*t*), the number of new cases must also be smaller at time *t* + 1 than it was at time *t*. Independent of the number of days *τ* that it takes for an infected individual to become infectious, the number of new infections on day *t* + *τ* is less than on day *t* for all *t* > *T*_2_.

As a result, when the cases in an epidemic follow a Gompertz Function, for all times *t* > *T*_2_, *R*_*t*_ is less than 1–so *X*(*T*_2_) is the proportion of the population infected in order for the epidemic to begin to decline. This is the Herd Immunity Threshold.^18^

The generically good fits of the Gompertz Function Model to epidemic data makes it clear that the empirically observed Herd Immunity Threshold across all of the outbreaks illustrated above is always close to 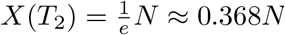.

## 4 Extending the Gompertz Function Model from Epidemic to Endemic Disease

For an epidemic which passes the Herd Immunity Threshold and continues to follow a Gompertz Function Model, *R*_*t*_ will tend to 0 and the epidemic will simply die out (as in the case of the Danish Cholera epidemic of 1853).

But rather than dying out, the disease may become endemic with periodic outbreaks.These may be sporadic as is the case with Ebola. Or the outbreaks may follow an annual cycle–as they do in influenza.

In endemic disease, *R*_*t*_ must be very close to 1. If it were exactly 1 then the growth in cumulative cases would be linear. Conversely, if the cumulative is exactly linear, then the number of cases between *t* and *t* + *t*_1_ is constant for all values of *t*_1_ and this means that *R*_*t*_ = 1.

If *R*_*t*_ is very close to 1, there will be fluctuations in the number of daily cases but the graph of cumulative cases from time *t* to *t* + *t*_1_ will be nearly a straight line whose slope is the average number of cases in that period.

So if an epidemic ends with a transition from Gompertz Function growth to linear growth, this is an indication that the disease has become endemic.

In our analysis of the initial Covid-19 outbreaks we saw *exactly* this process repeated again and again: Gompertz Function growth ended with a transition to *linear* growth. For example, Figure 51 shows the best line fit to London Covid-19 Hospital Admissions after the Gompertz Function growth shown in Figure 23).The linear fit has *r*^2^ = 0.997, its maximum absolute error is less than 0.6% and its average absolute error is only 0.24%. Linear growth is not just a good approximation. It is an extremely good approximation.^19^

**Figure 51:**
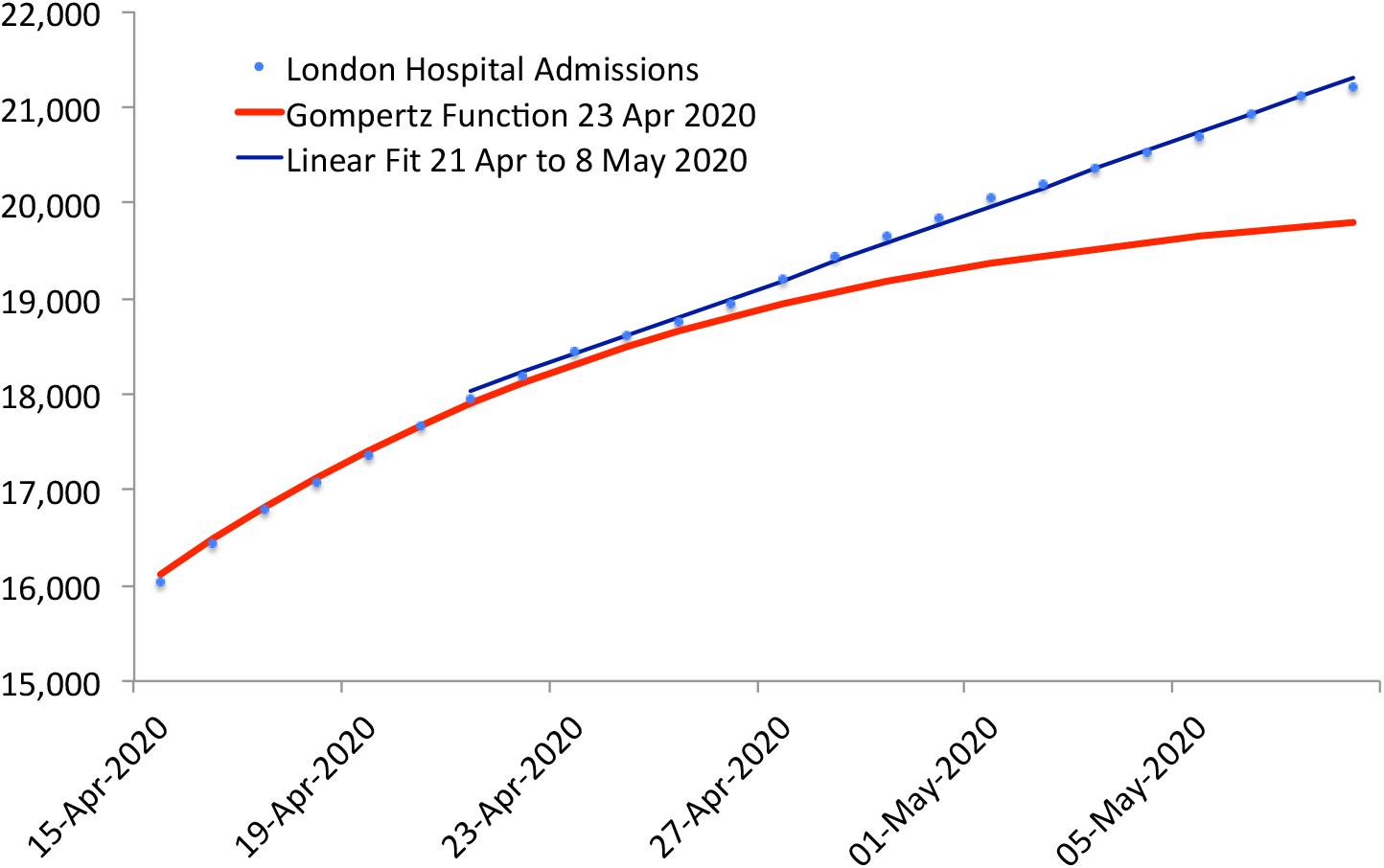
Transition to linear growth in London Hospital Admissions April 2020.

Influenza is an endemic disease which has regular outbreaks of epidemic infection. Thus it provides a natural ‘observatory’ for the endemic-epidemic cycle. In the next section we show that transition from Gompertz Function growth to linear growth (twice) is precisely what happens in influenza cases in Portugal in a regular annual pattern.

### 4.1 An Influenza Observatory

The Portuguese government has published daily influenza case data beginning 1 November 2016.^20^.The main ‘Flu Season’ runs from approximately November to April.^21^ Figure 52 shows the annual cycle for Influenza Cases recorded by the Portuguese National Health Service from 1 November 2016 to 31 May 2020. Each year there is an annual winter peak followed by a dramatic reduction in cases to a much lower level through the late spring and summer. But cases never die out completely and in early autumn they begin to rise again.

**Figure 52:**
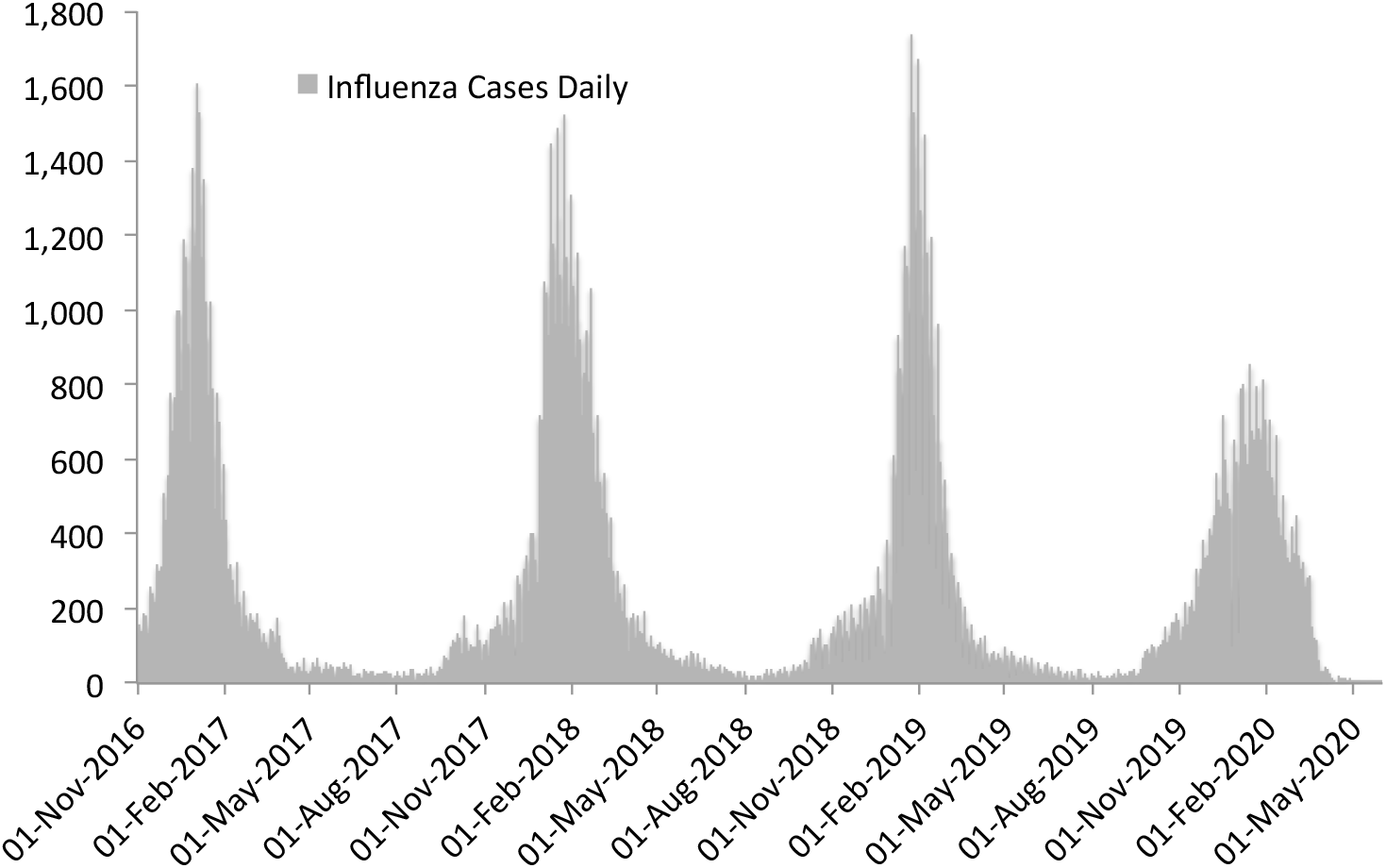
Portuguese Influenza Cases 1 Nov 2016 to 31 May 2020.

The graph of cumulative cases (Figure 53) reveals further regularities with consistent annual patterns. The cumulative cases through each of the annual peaks follow Gompertz Function growth (shown in red).^22^

**Figure 53:**
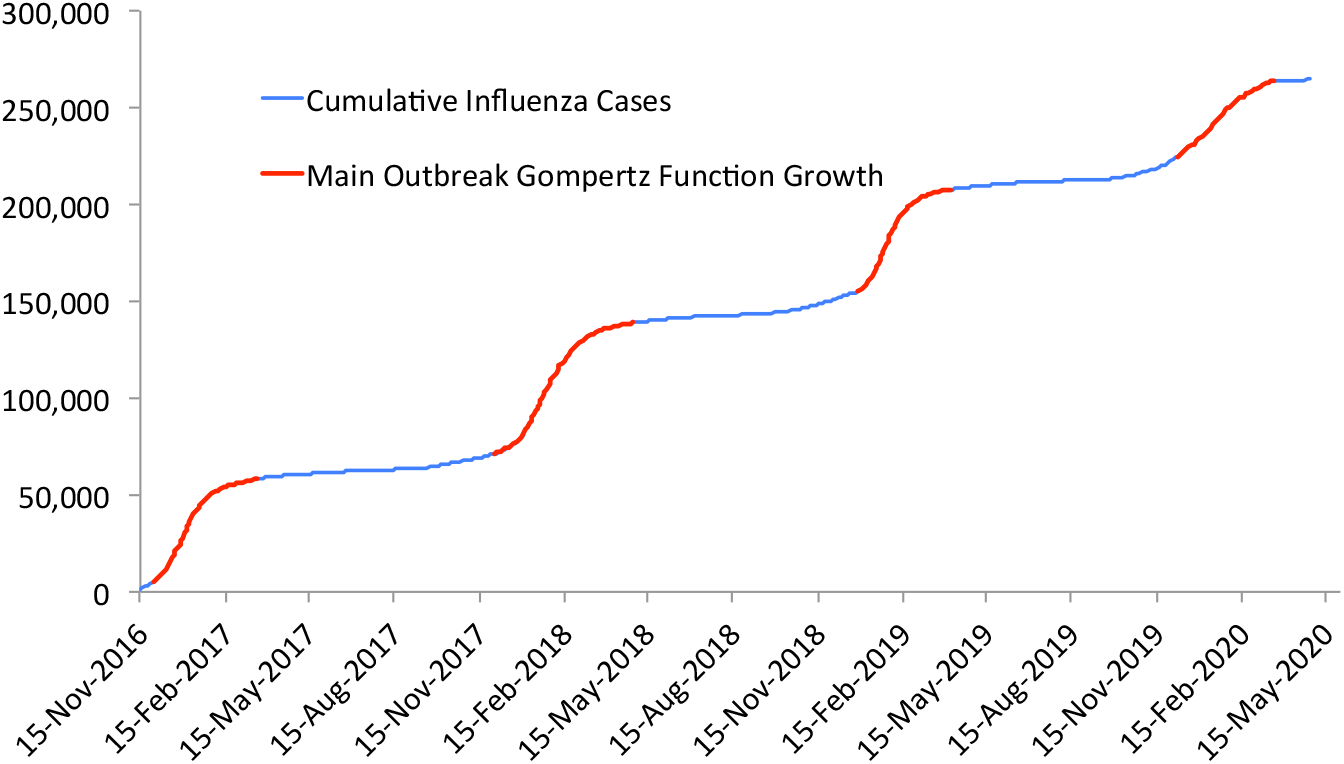
Cumulative Influenza Cases 1 Nov 2016 to 31 May 2020.

We have already shown (Figure 12) that the 2017-18 outbreak from 1 December 2017 until 30 April 2018 followed the Gompertz Function Model. Figures 54 to 56 show the same thing for the remaining years in Figure 53. Figure 57 shows that even though Influenza cases have become much less prevalent, the same Gompertz Function growth marks the main seasonal outbreak in 2020-2021.

**Figure 54:**
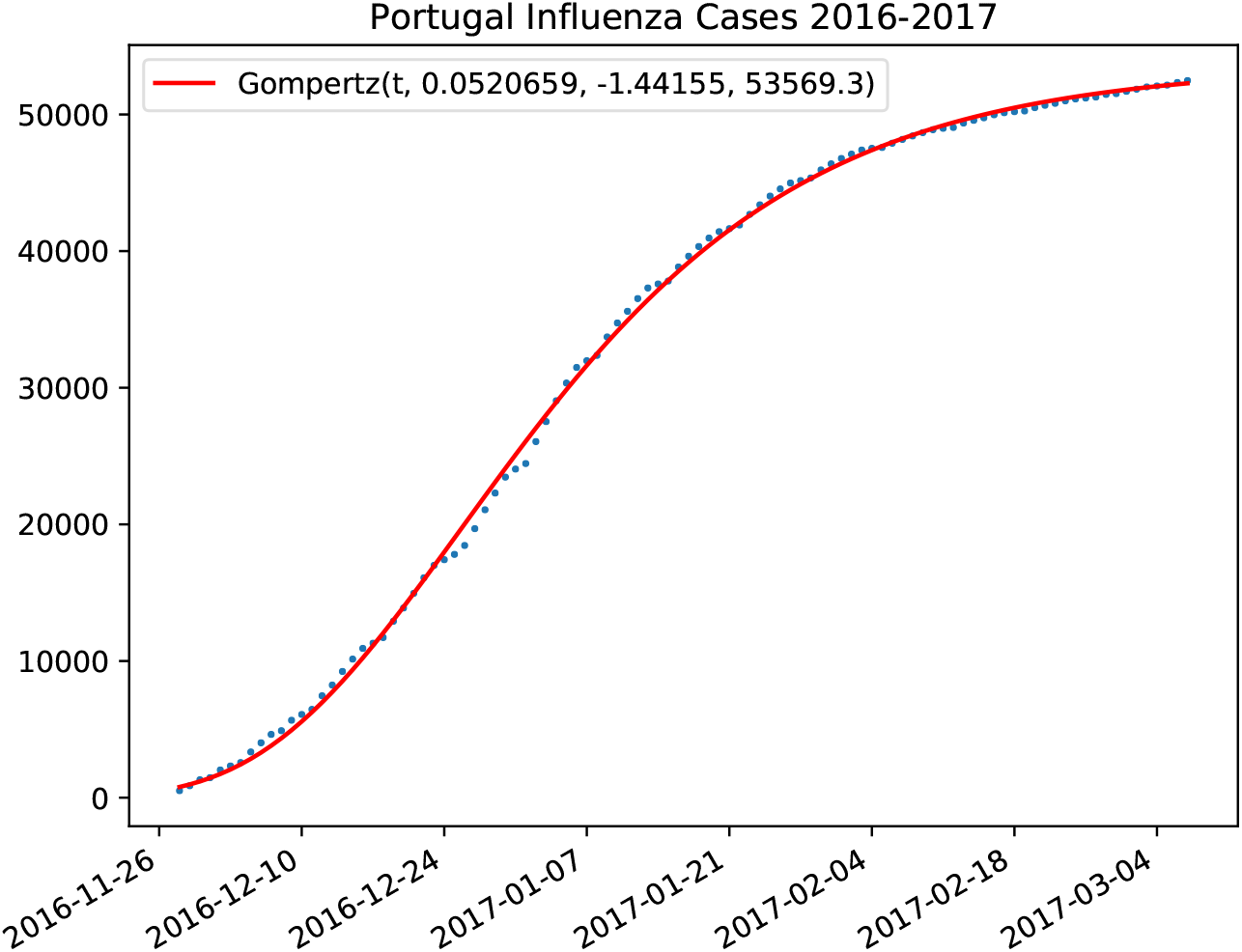
Portugese Influenza Cases 28 Nov 2016 to 7 Mar 2017.

**Figure 55:**
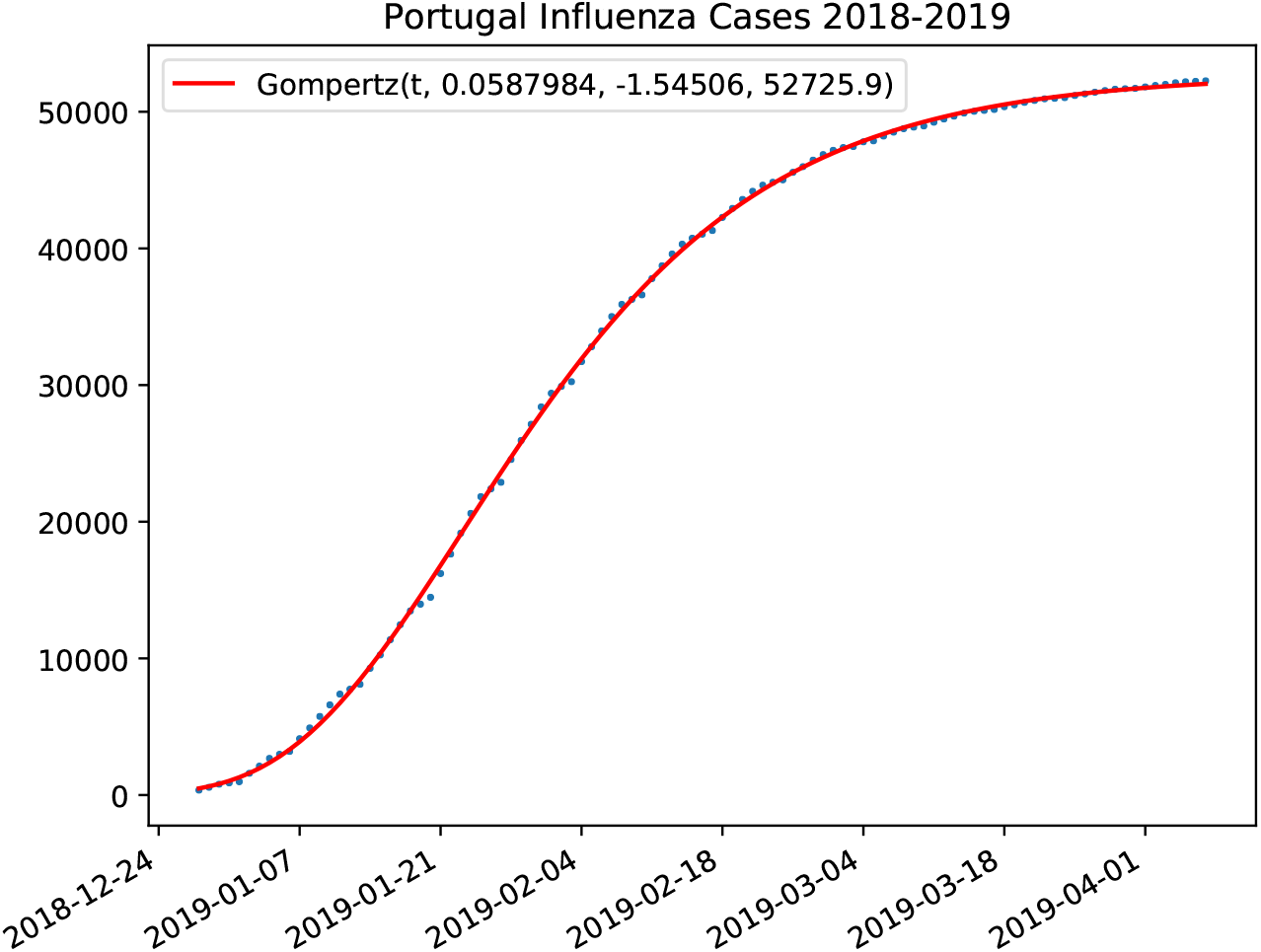
Portuguese Influenza Cases 28 Dec 2018 to 7 Apr 2019.

**Figure 56:**
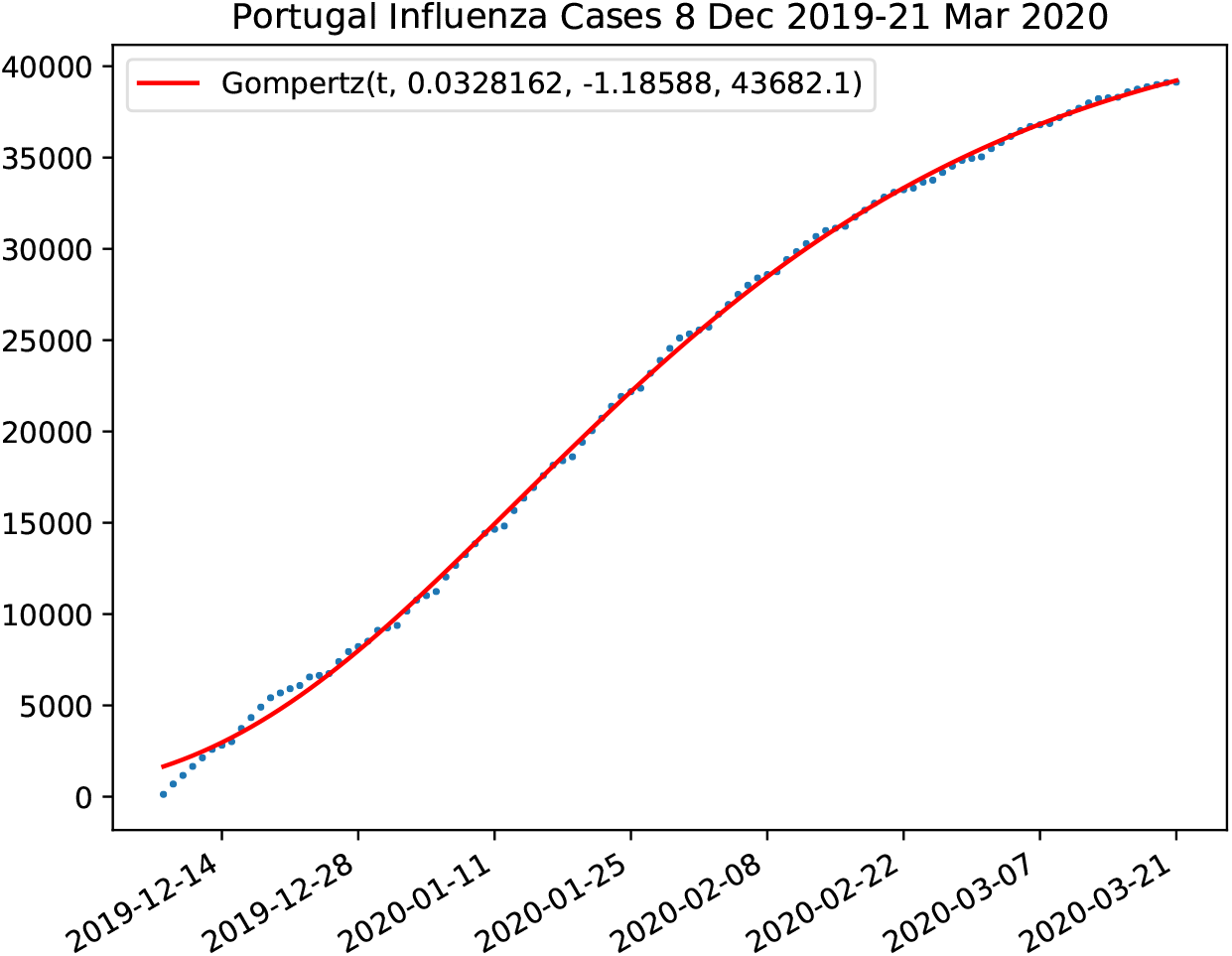
Portuguese Influenza Cases 8 Dec 2019 to 21 Mar 2020.

**Figure 57:**
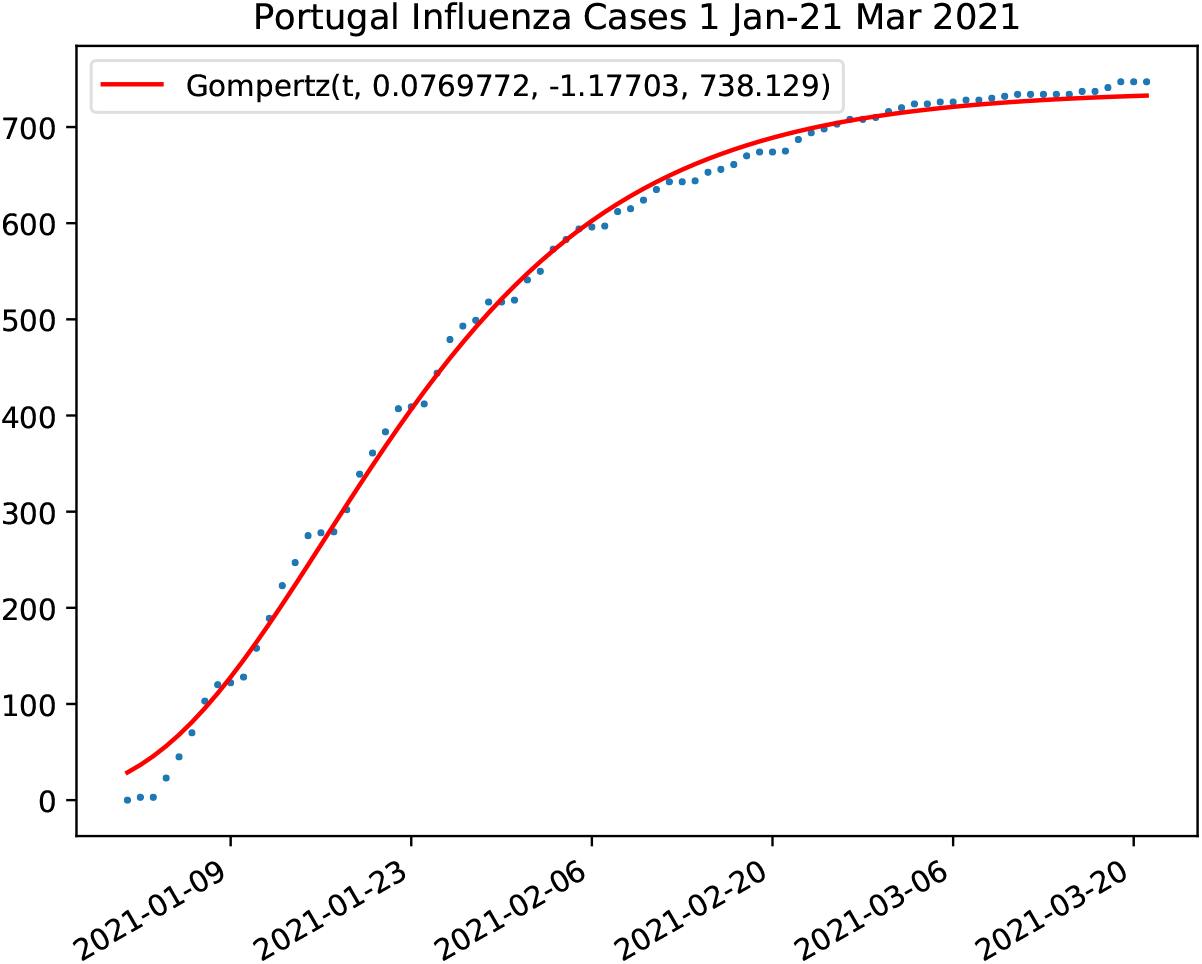
Portuguese Influenza Cases 1 Jan to 21 Mar 2021.

Another important regularity which is visible in the cumulative cases is that the periods of Gompertz Function growth end in a transition to linear growth (just as we have already observed was the case with the initial Covid-19 outbreaks).But each of the main influenza outbreaks is also *preceded* by a period of linear growth.

Figures 58 and 59 show the linear entry to and exit from the 2016-17 Gompertz Function growth phase.The linear fits to the cumulative case graphs shown in these entry and exit figures have *r*^2^ values of over 0.99 and very small absolute errors between the data and the linear fit– so this is not simply ‘close’ to linear, it is almost exactly linear.

**Figure 58:**
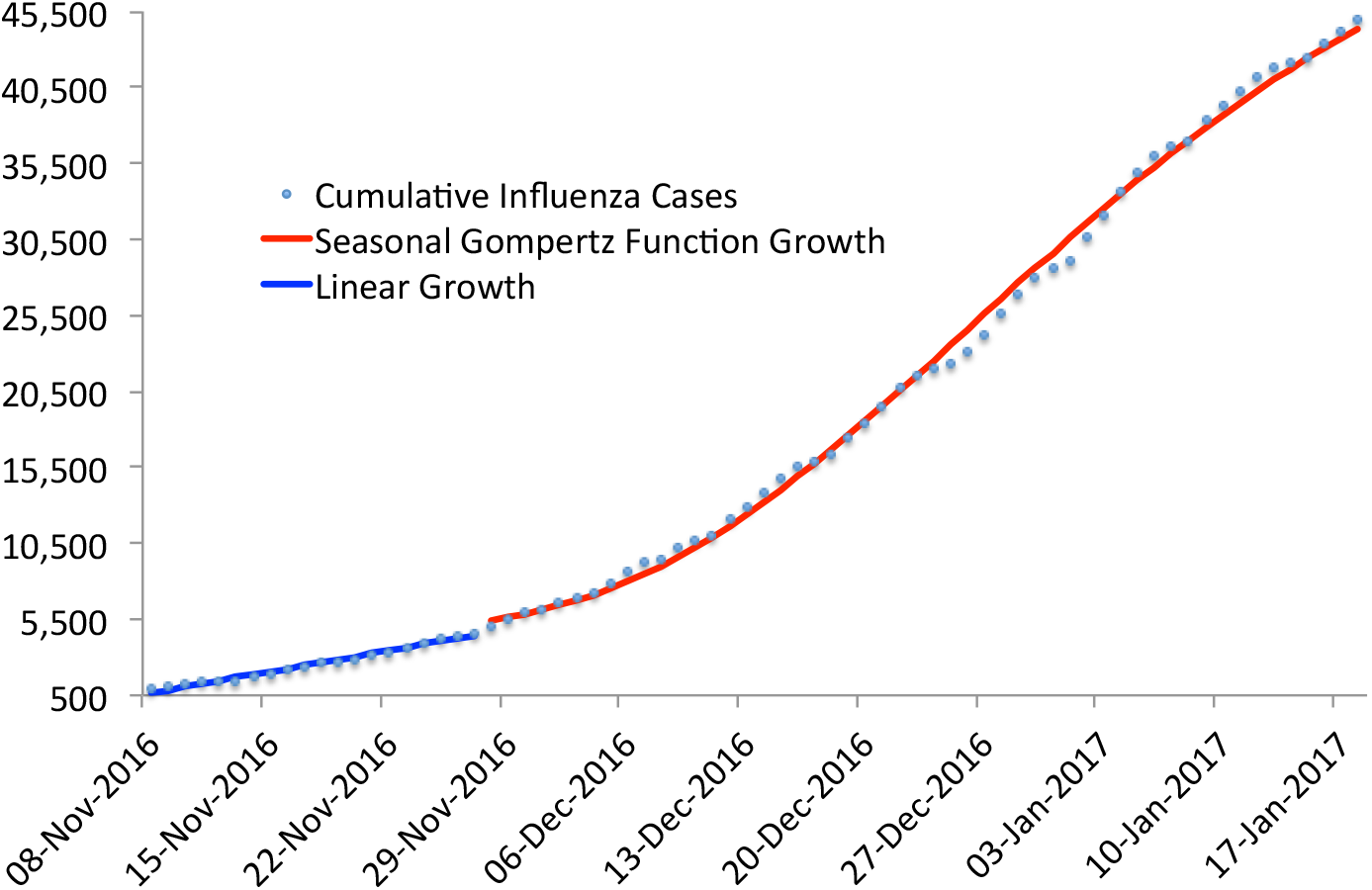
The 2016-2017 Influenza Season begins with the transition from Linear to Gompertz Function Growth.

**Figure 59:**
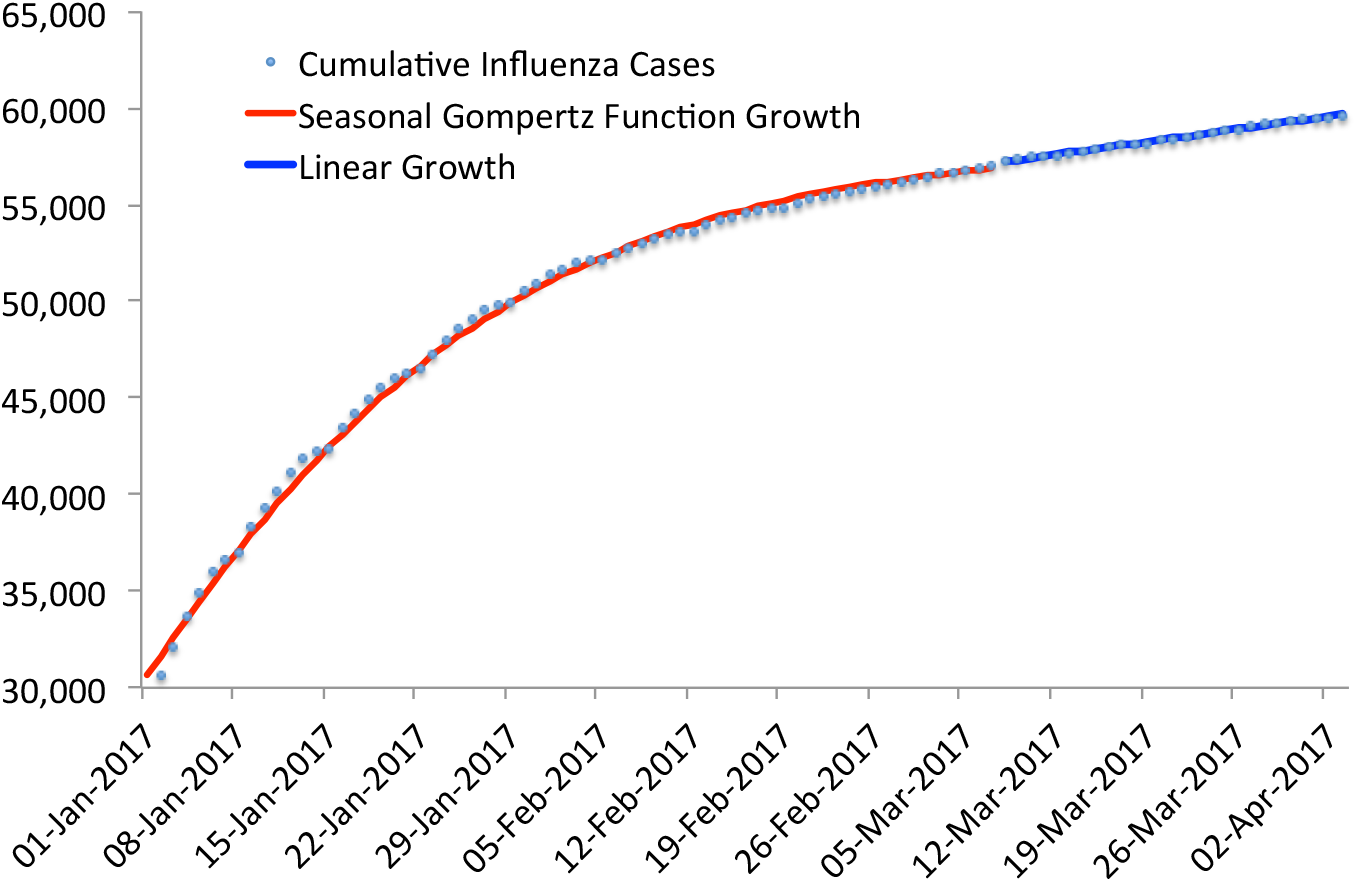
The 2016-2017 Influenza Season ends with the transition from Gompertz Function Growth to Linear Growth.

There is a further regularity in the ‘off season’ portions of the cumulative case data.The growth in cumulative cases continues to be piecewise linear with drops in the slope though the summer minimum. For example, in 2017 the exit from Gompertz Function growth in April had a slope of 102 cases per day which dropped in two stages to only 21 cases per day by August.

But the entry to the Gompertz Function growth phase in November 2017 (Figure 60) had a slope of 125 cases per day. How did the case rate increase to this level?

**Figure 60:**
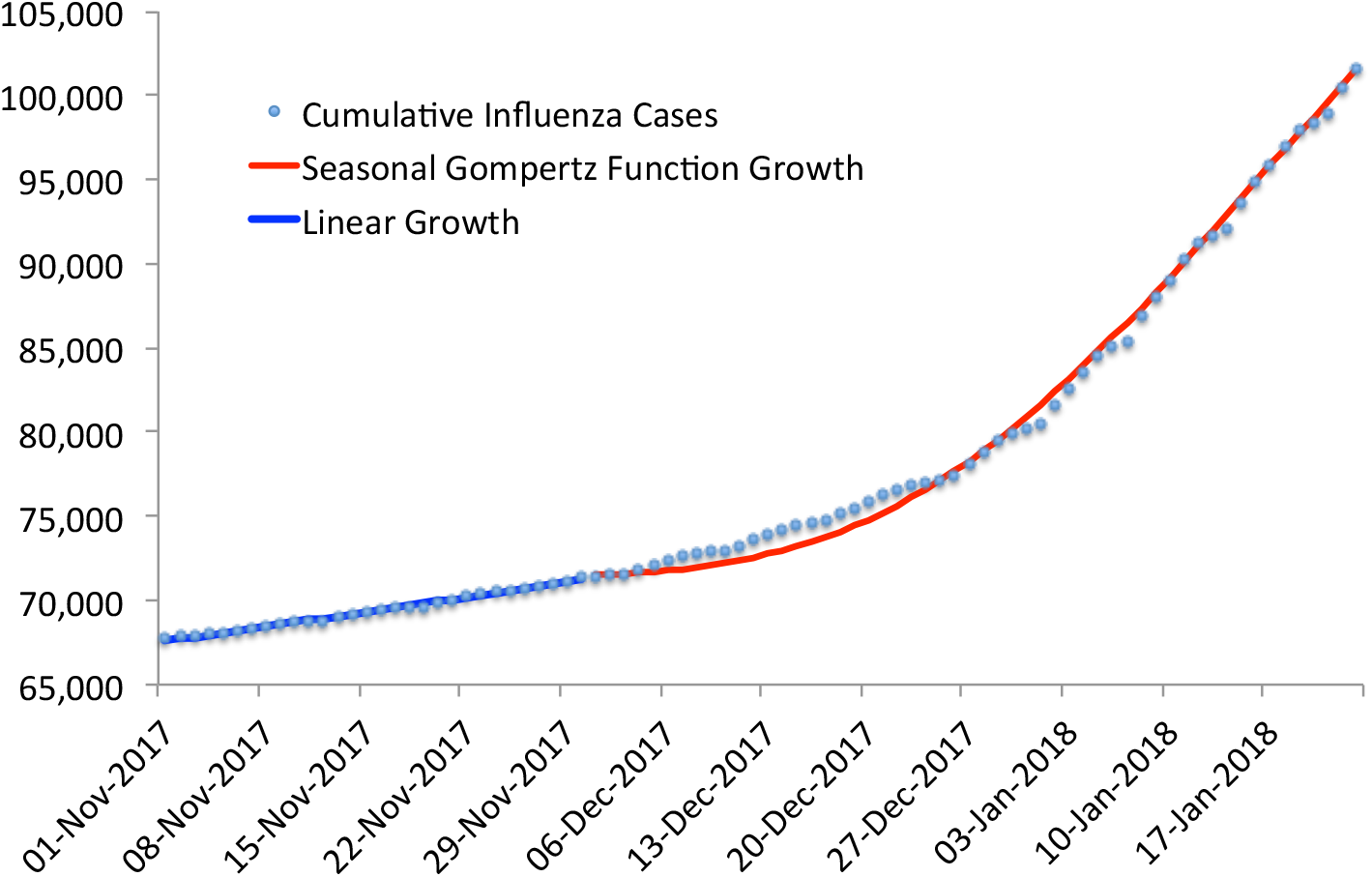
The main outbreak of the 2017-2018 Influenza Season begins with the transition from linear to Gompertz Function growth.

Our conjecture when we observed this was that there was an intermediate phase of rapid Gompertz Function growth whose exit is to linear growth at the higher rate. This is consistent with the apparent universality of Gompertz Function growth in epidemic outbreaks that we have seen all of our other examples.

And this is precisely what we found in the Portuguese Influenza case data–not just in 2017 but each year, including 2020.

Figures 61 and 62 show the cycle from January to December 2017 and the detail of the piecewise linear spring and summer decline followed by a Gompertz Function growth outbreak in September and October.

**Figure 61:**
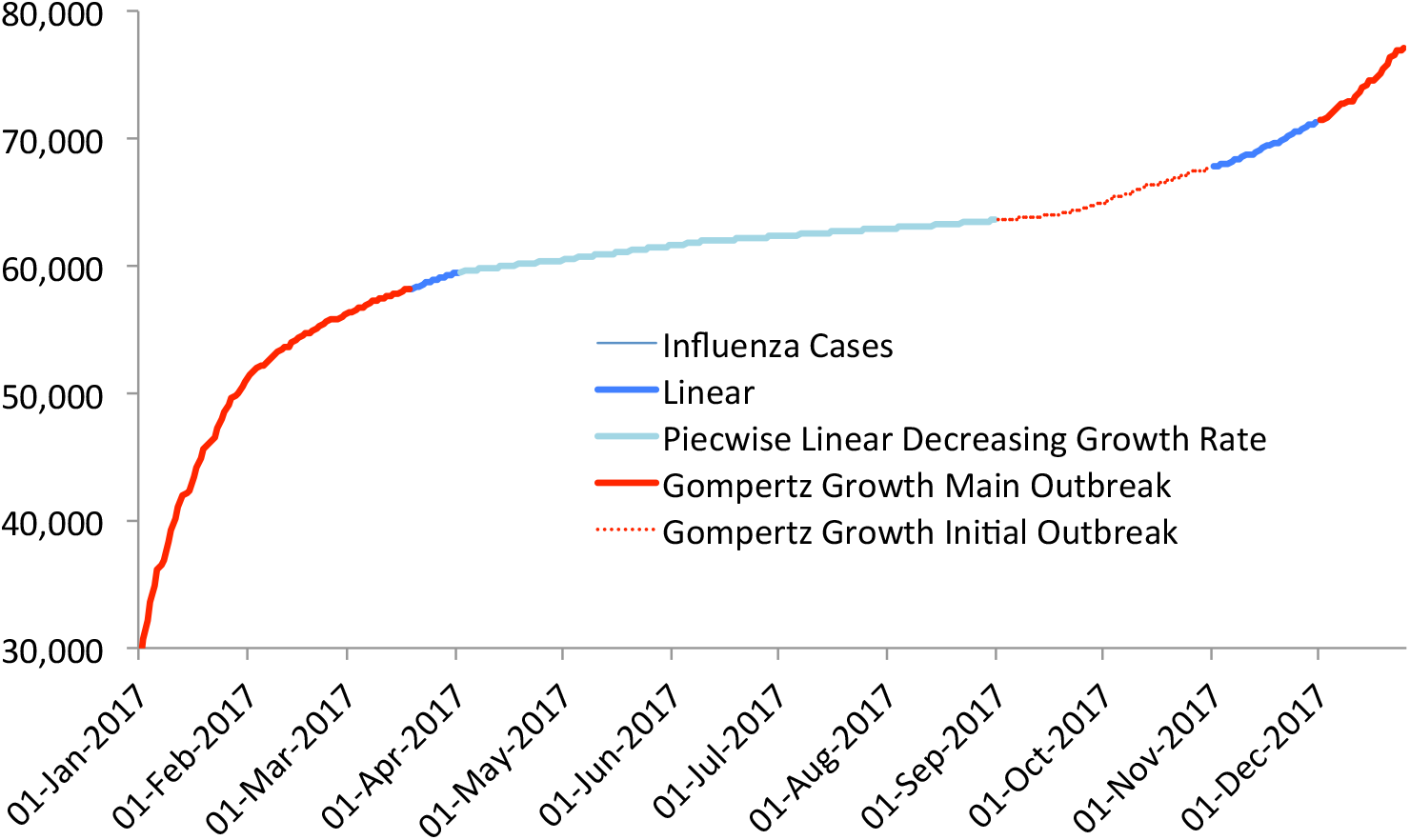
The annual cycle Gompertz Growth to Linear Growth to Gompertz Growth.

**Figure 62:**
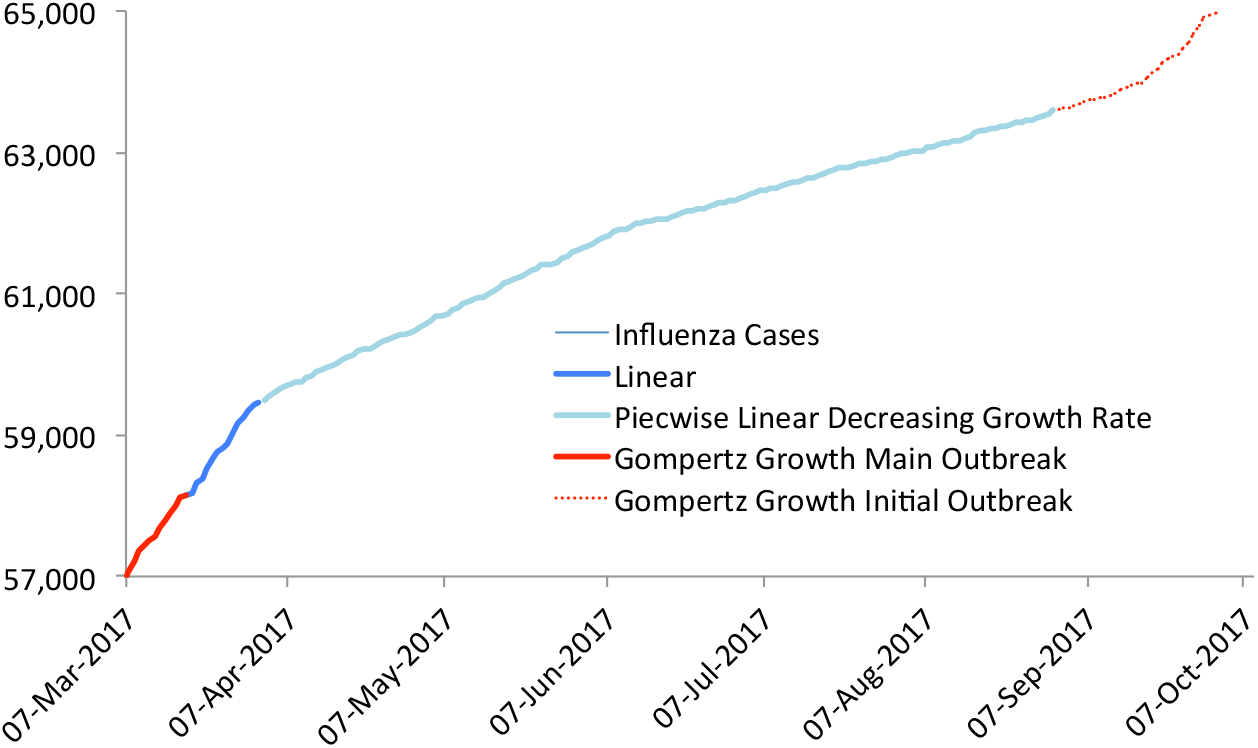
Detail of the 2016-17 cycle shows declining growth in the piecewise linear phase and a very rapid increase in September.

This is followed (as Figure 60 showed in closeup) by the linear transition to the main Gompertz Growth phase in November and December 2017.

The intermediate Gompertz Function growth phase is repeated each year at almost the same time.(See Figures 63 to 66).Table 1 shows the dates of the Gompertz Function growth and linear growth cycles from December 2016 to March 2021.

**Figure 63:**
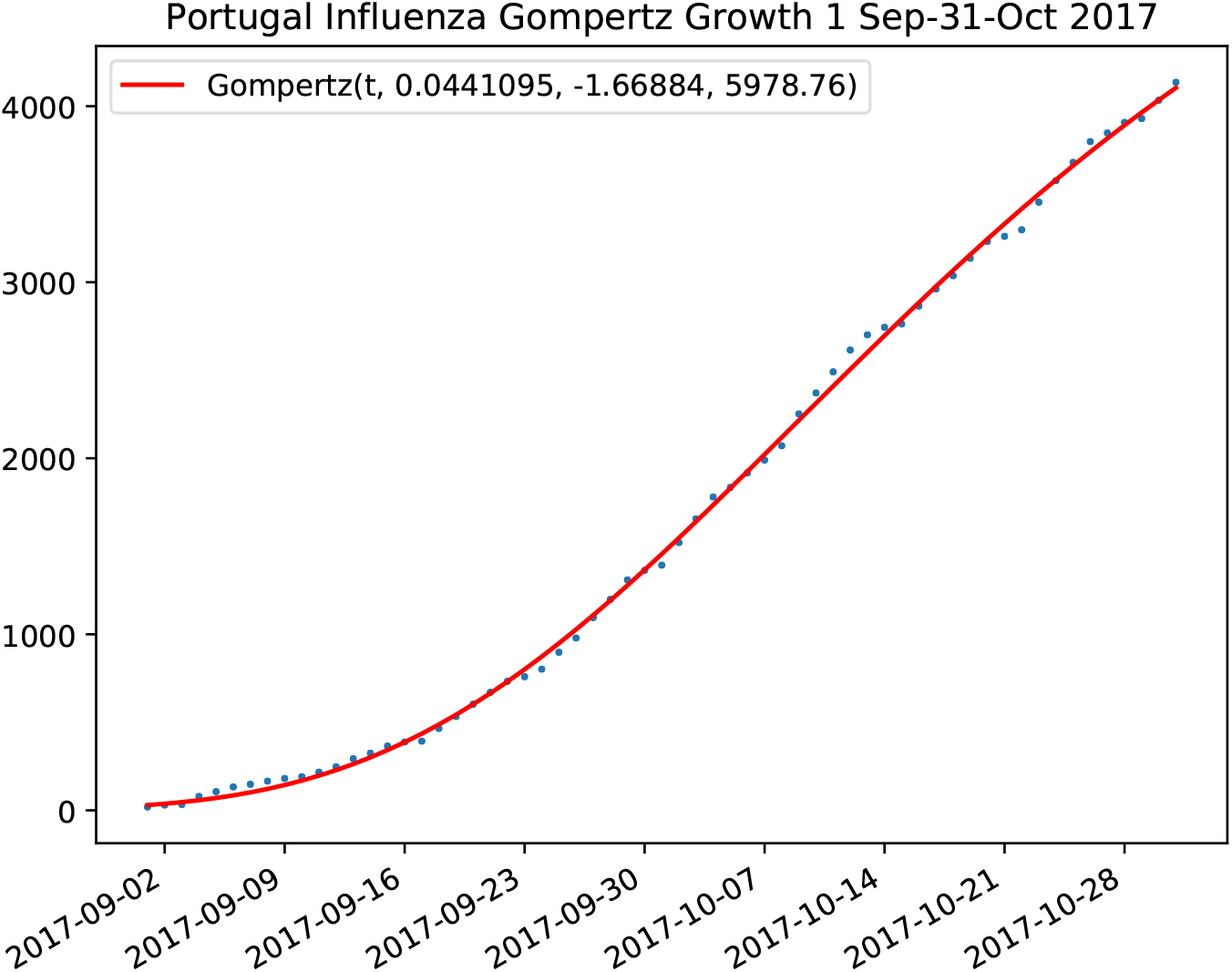
Gompertz Growth of Influenza Cases 1 Sep to 31 Oct 2017.

**Figure 64:**
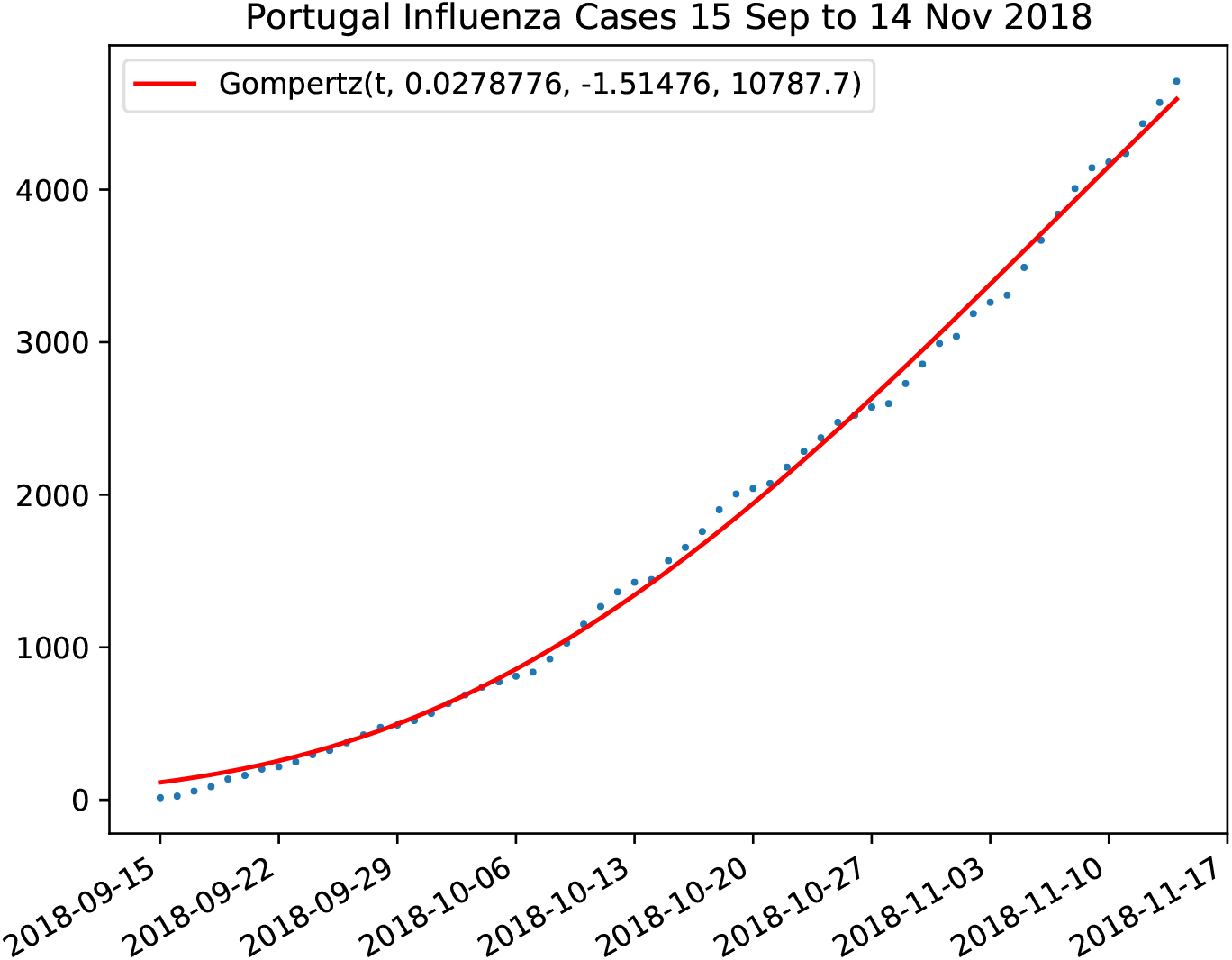
Gompertz Growth of Influenza Cases 15 Sep to 14 Nov 2018.

**Figure 65:**
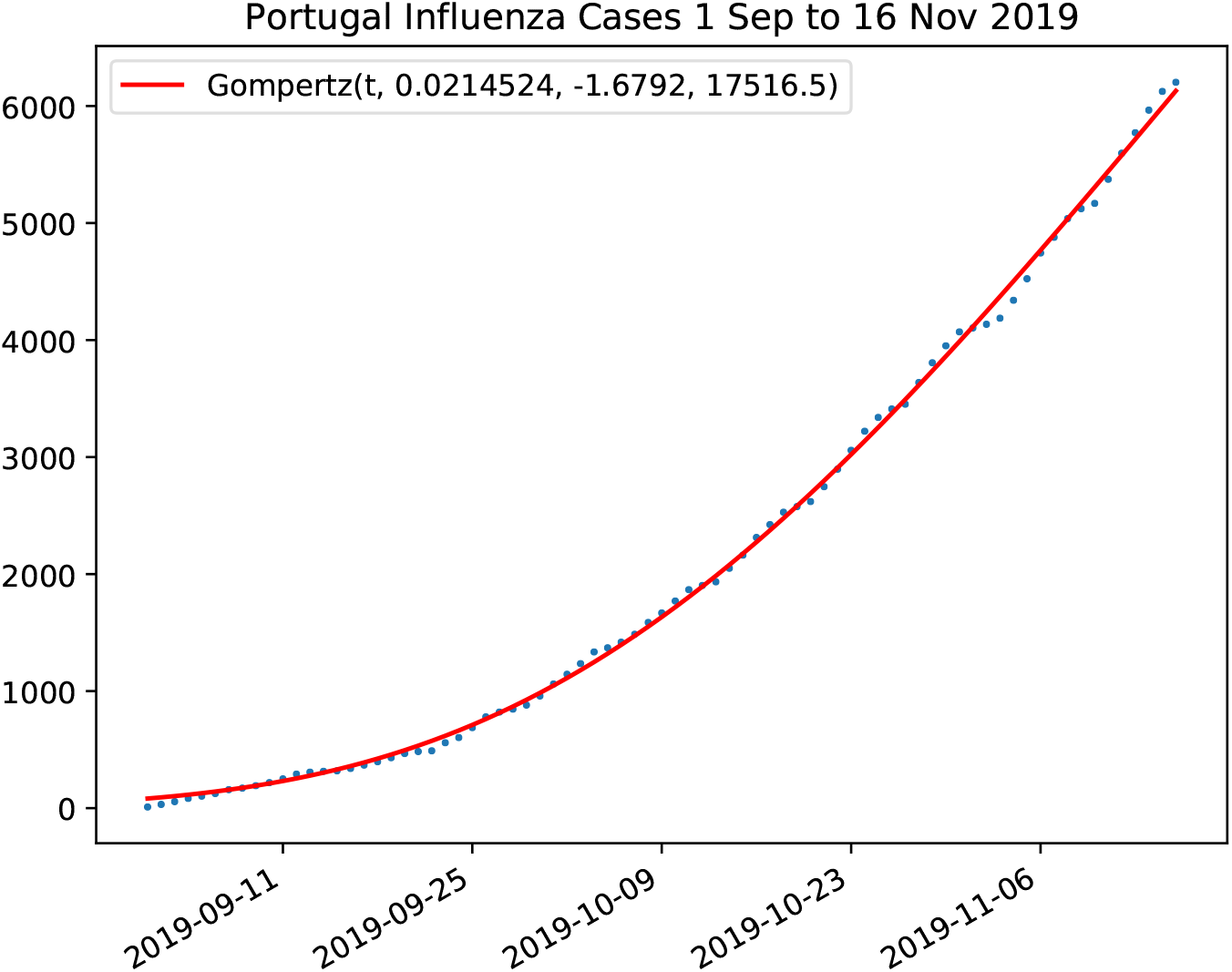
Gompertz Growth of Influenza Cases 1 Sep to 16 Nov 2019.

**Figure 66:**
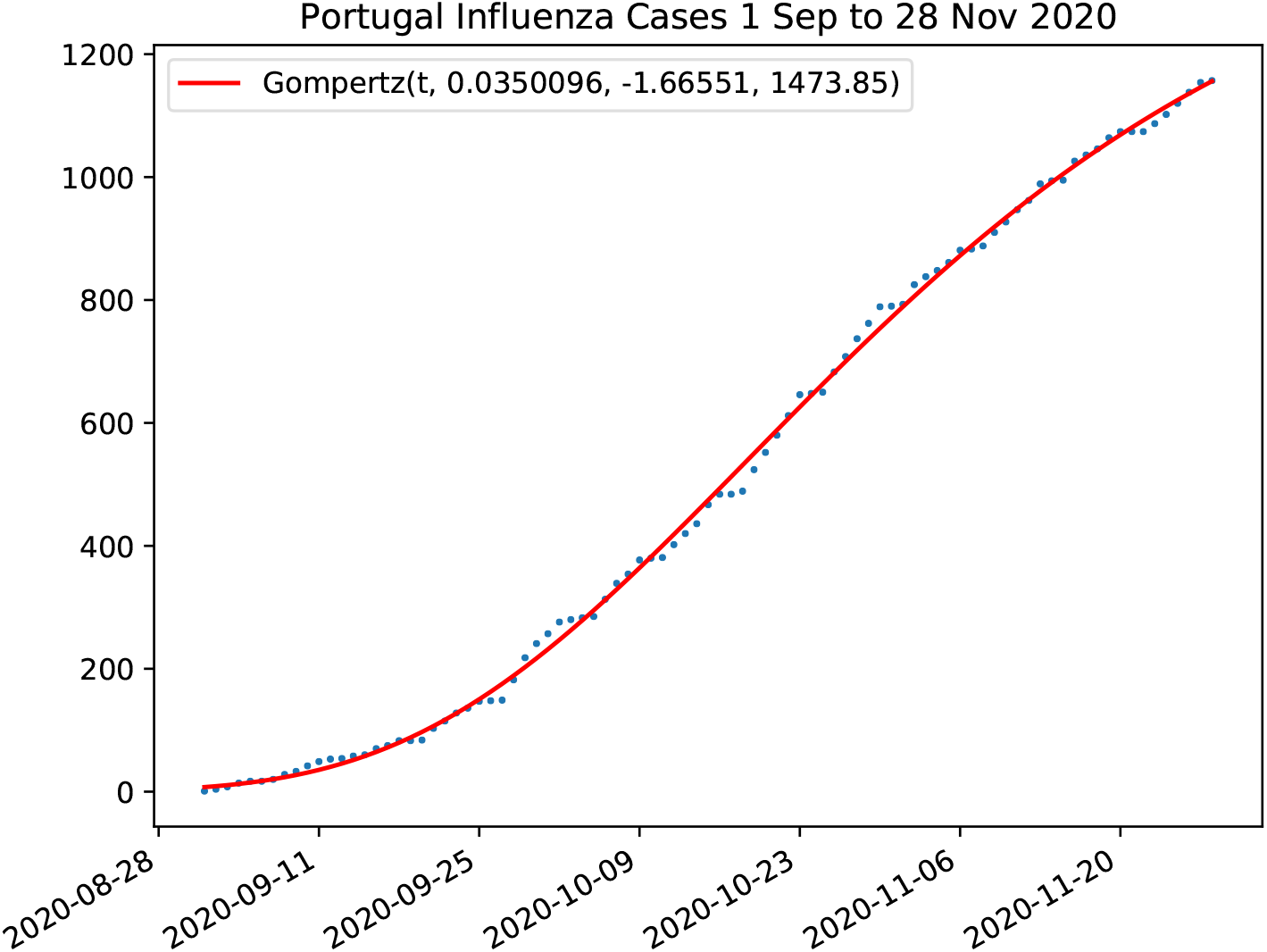
Gompertz Growth of Influenza Cases 1 Sep to 28 Nov 2020.

**Table 1:**
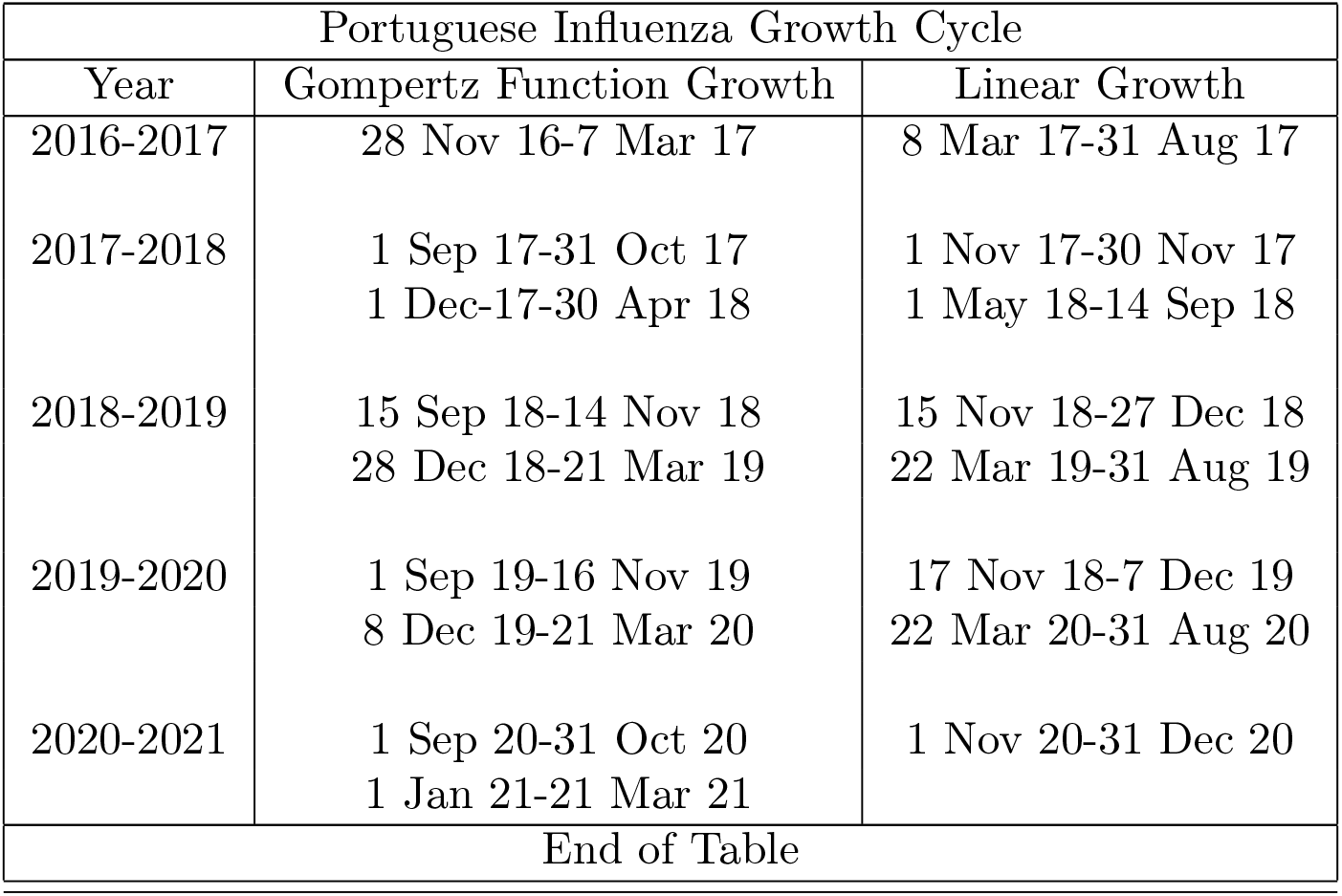
Portuguese Influenza Cycle 2016-2021.

It is clear that, at least in Portugal, the influenza cycle has, to a very good approximation, two modes: linear growth and Gompertz Function growth. Moreover, the Gompertz Function growth phases agree with Edgar Hope-Simpson’s cycle in the Northern Temperate Zone [9] –so we expect to see this pattern repeated in daily influenza cases in other countries as well.

If this conjecture is born out, it would have very important implications for the prediction of the influenza cycle and the resulting demands on healthcare systems which are often severely stressed in annual influenza outbreaks. During these periods the Gompertz Function Model has good predictive power. In the piecewise linear growth periods linear extrapolation will suffice.

The prediction problem is therefore reduced to identifying the transitions, especially from linear to Gompertz Function growth.

We now show the same alternation in growth separated waves in the ‘Spanish Flu’ epidemic.

### 4.2 Spanish Flu Waves

The worst pandemic of the last century, the ‘Spanish Flu’, was felt in waves in various locations around the world.[6]

In England and Wales, it came in three distinct waves, the mechanisms behind which are still uncertain.[20],[7] Weekly records of Spanish Flu deaths from the Registry for England and Wales^23^ record deaths parish by parish across England and Wales. The cumulative deaths for England and Wales combined show two periods of almost exactly linear growth: five weeks from 24 August to 21 September 1918 between the first and second waves and four weeks from 11 January and 1 February 1919, between the second and third. In both cases the linear fit to the data is almost exact with *r*^2^ over 0.99 and very small absolute errors (maximum of 0.11% in the first and 0.05% in the second) between the cumulative deaths data and linear fits. (See Figures 67 to 70). In the complement to the linear growth periods we see excellent fits by Gompertz Functions. (Figures 71, 72 and 73 show the fits to the cumulative cases from the beginning to the end of each period.) The calculation of *T*_2_ in each wave gives peaks which agree with the report of the Registrar for England and Wales’ description of the epidemic wave peaks [11].

**Figure 67:**
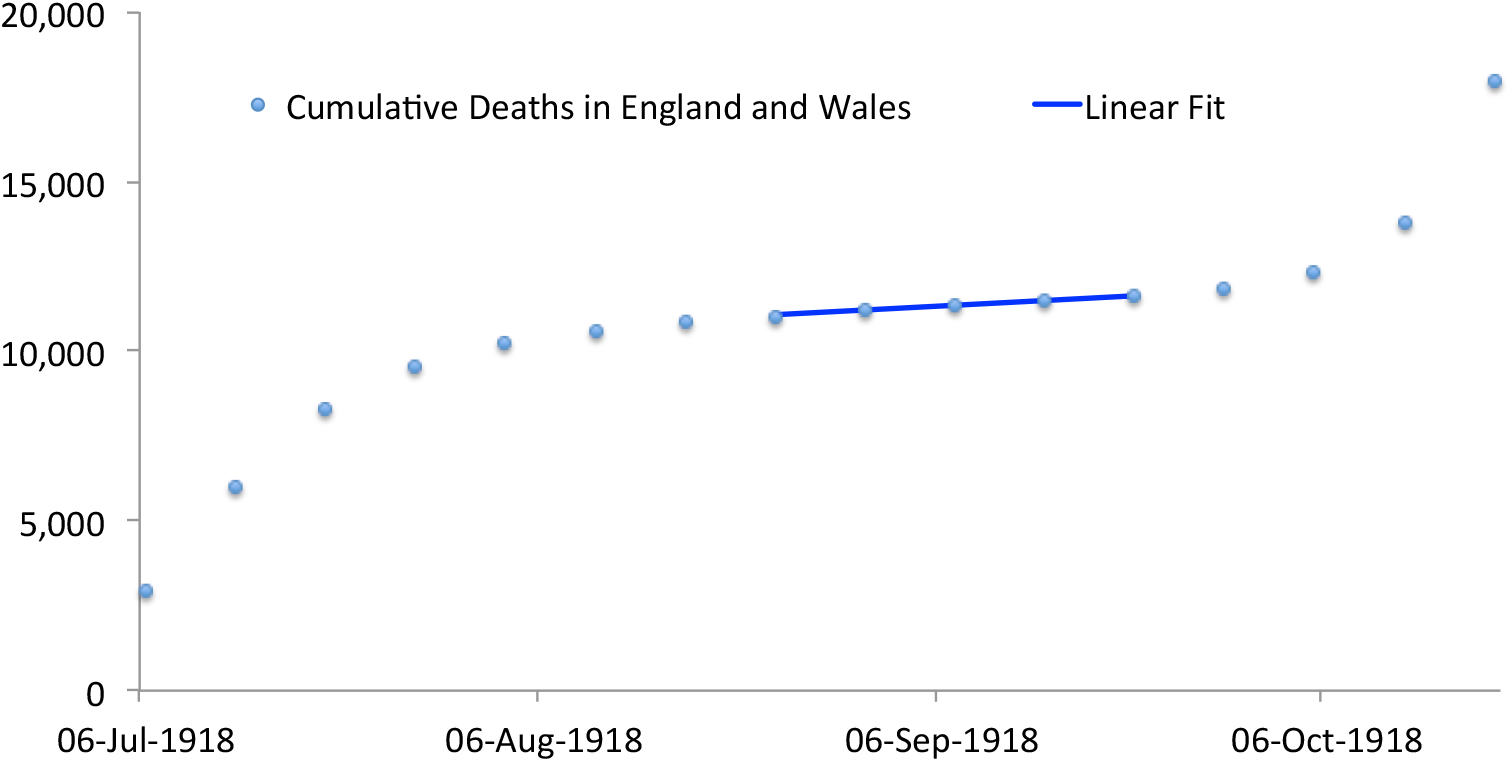
Spanish Flu Deaths Grew Linearly from 24 Aug to 21 Sep 1918.

**Figure 68:**
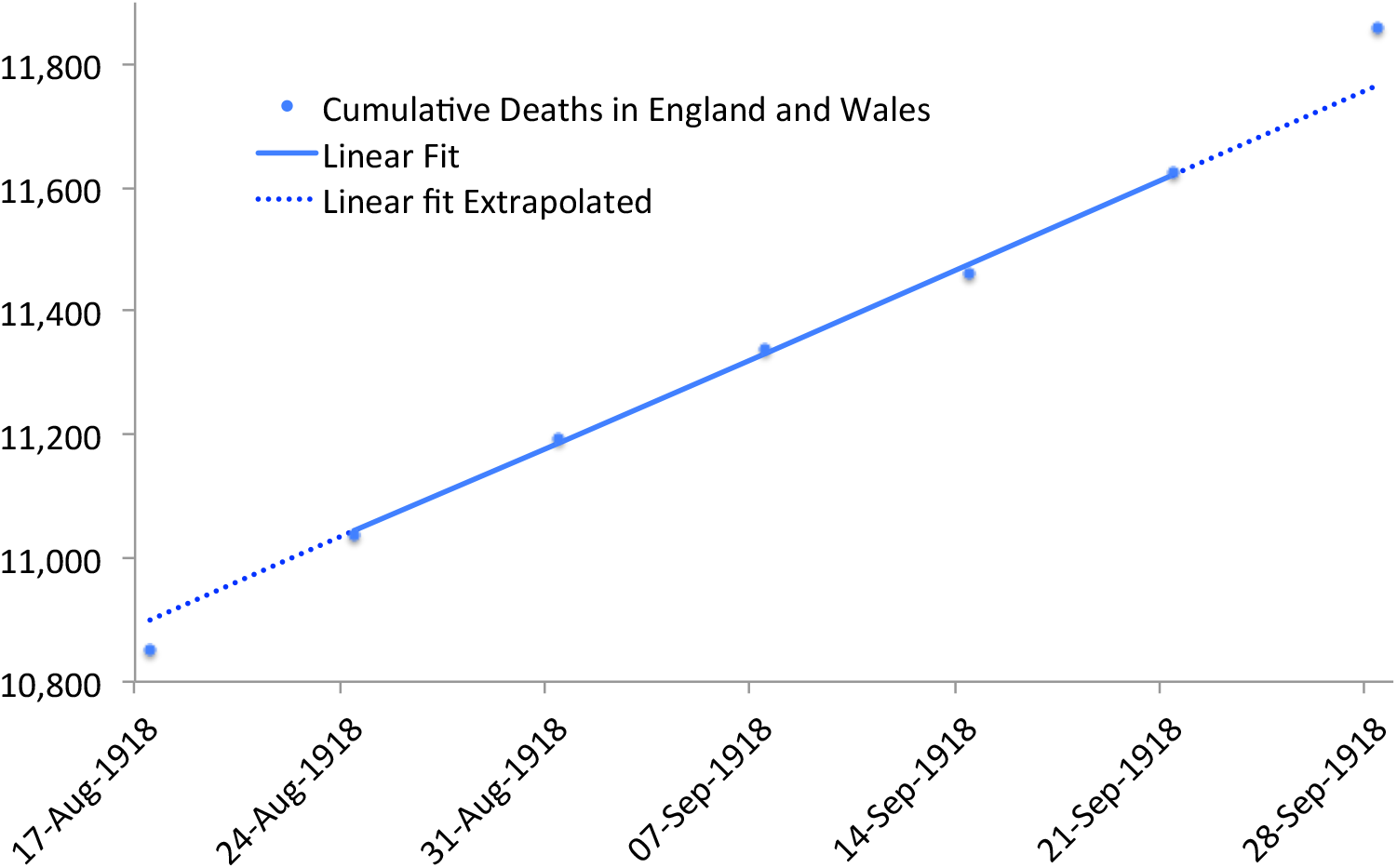
Detail Spanish Flu Deaths 24 Aug to 21 Sep 1918.

**Figure 69:**
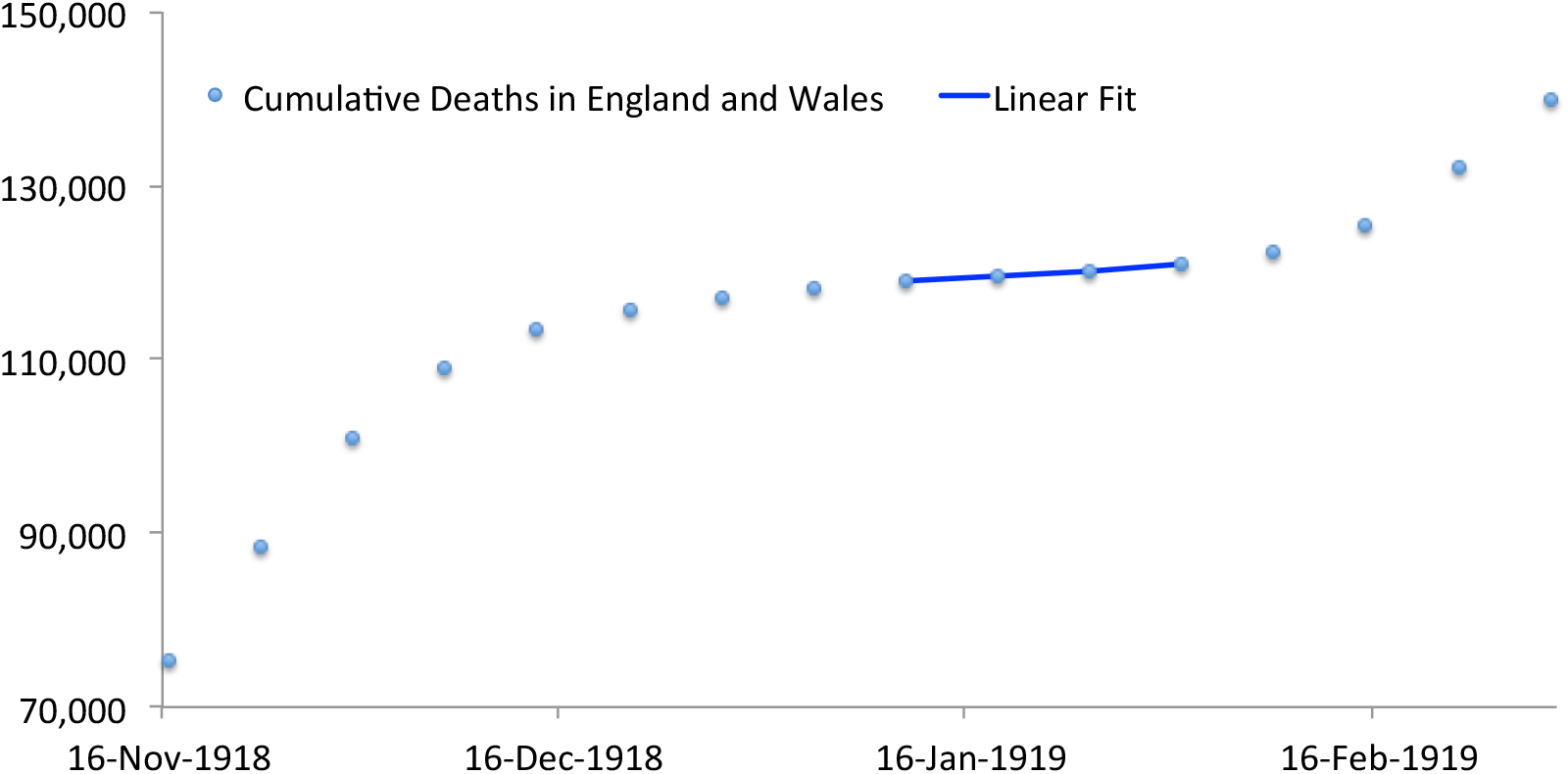
Spanish Flu Deaths Grew Linearly from 11 Jan to 1 Feb 1919.

**Figure 70:**
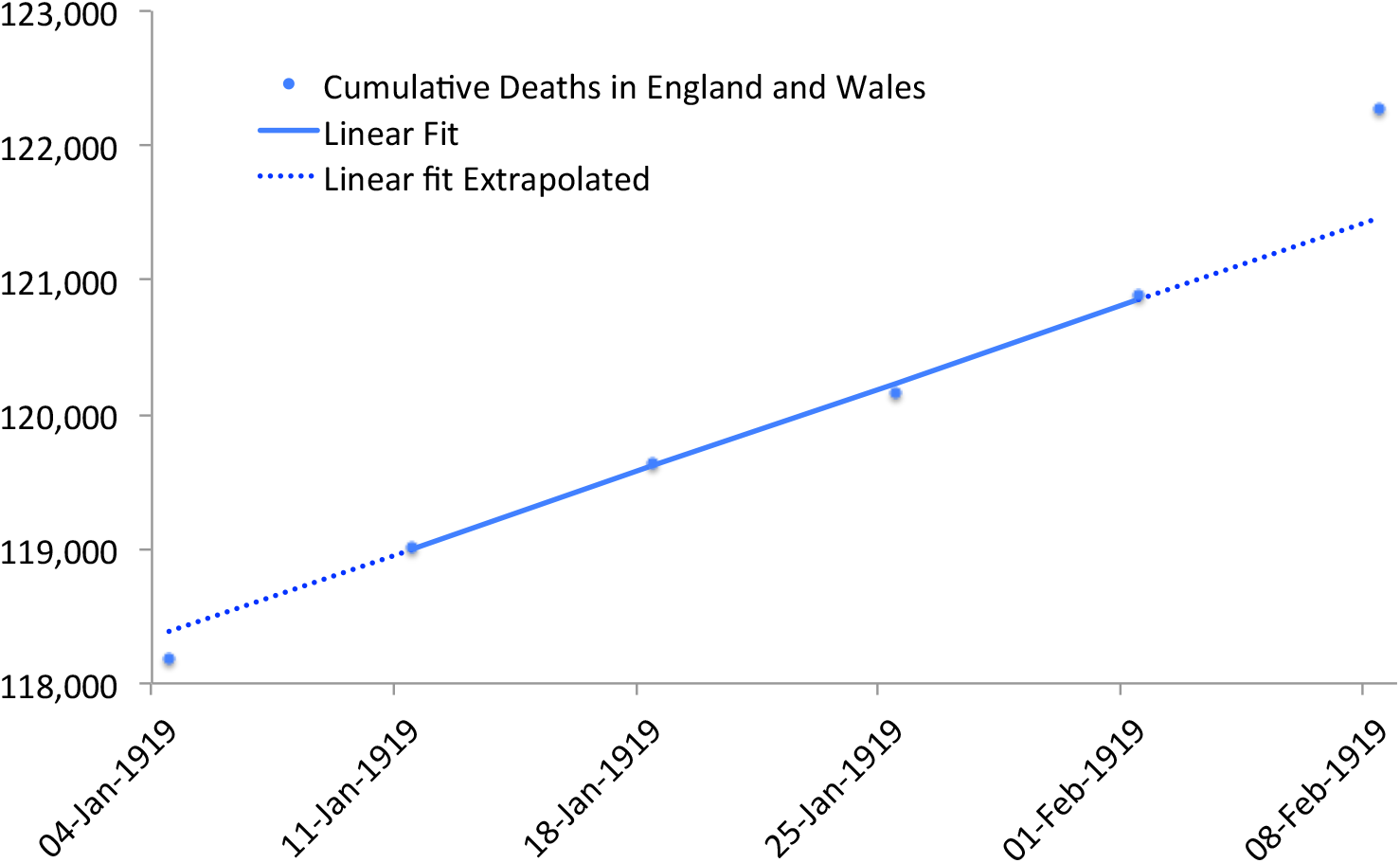
Detail Spanish Flu Deaths 11 Jan to 1 Feb 1919

**Figure 71:**
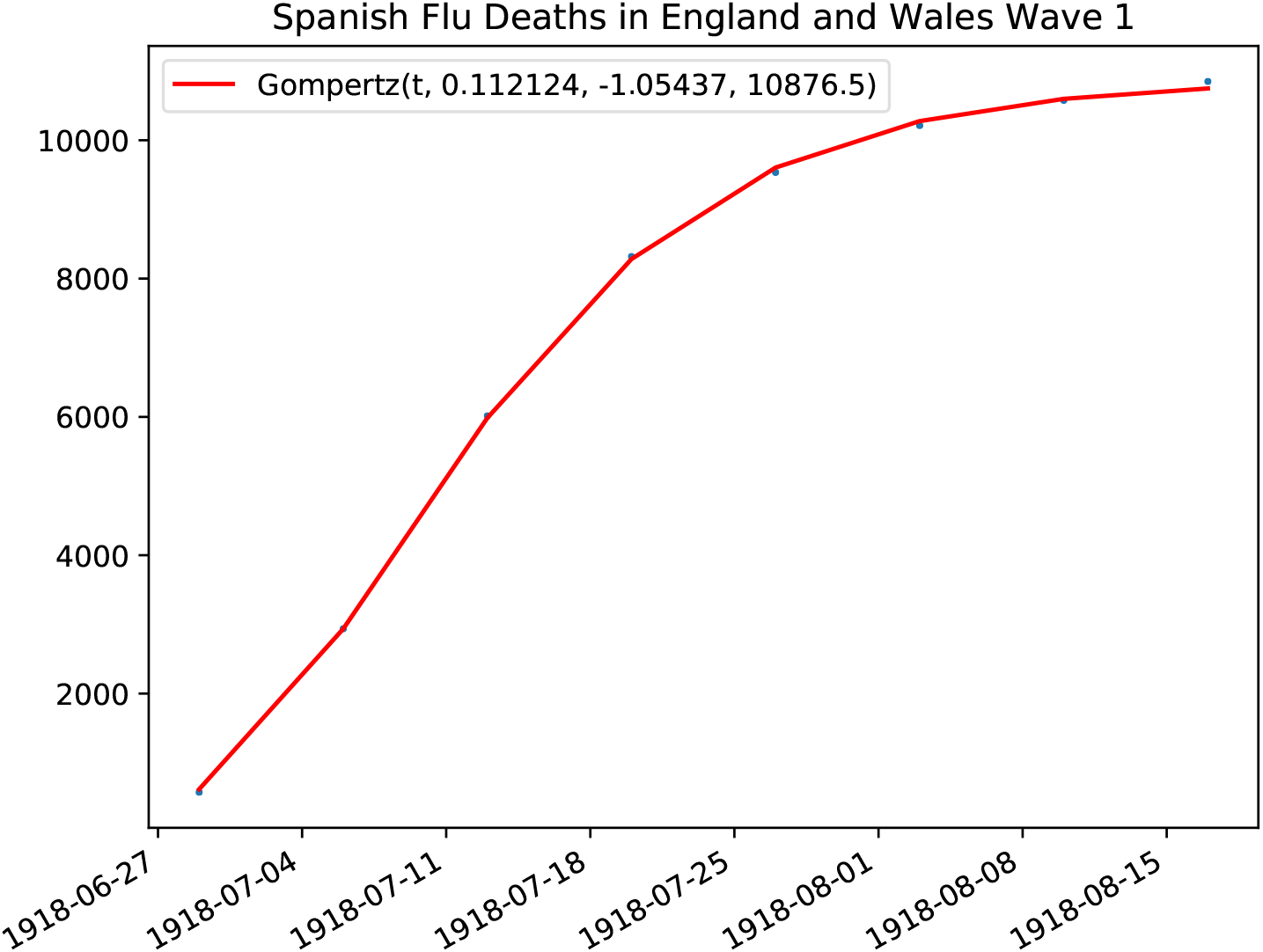
Spanish Flu deaths in England and Wales 29 Jun to 17 Aug 1918.

**Figure 72:**
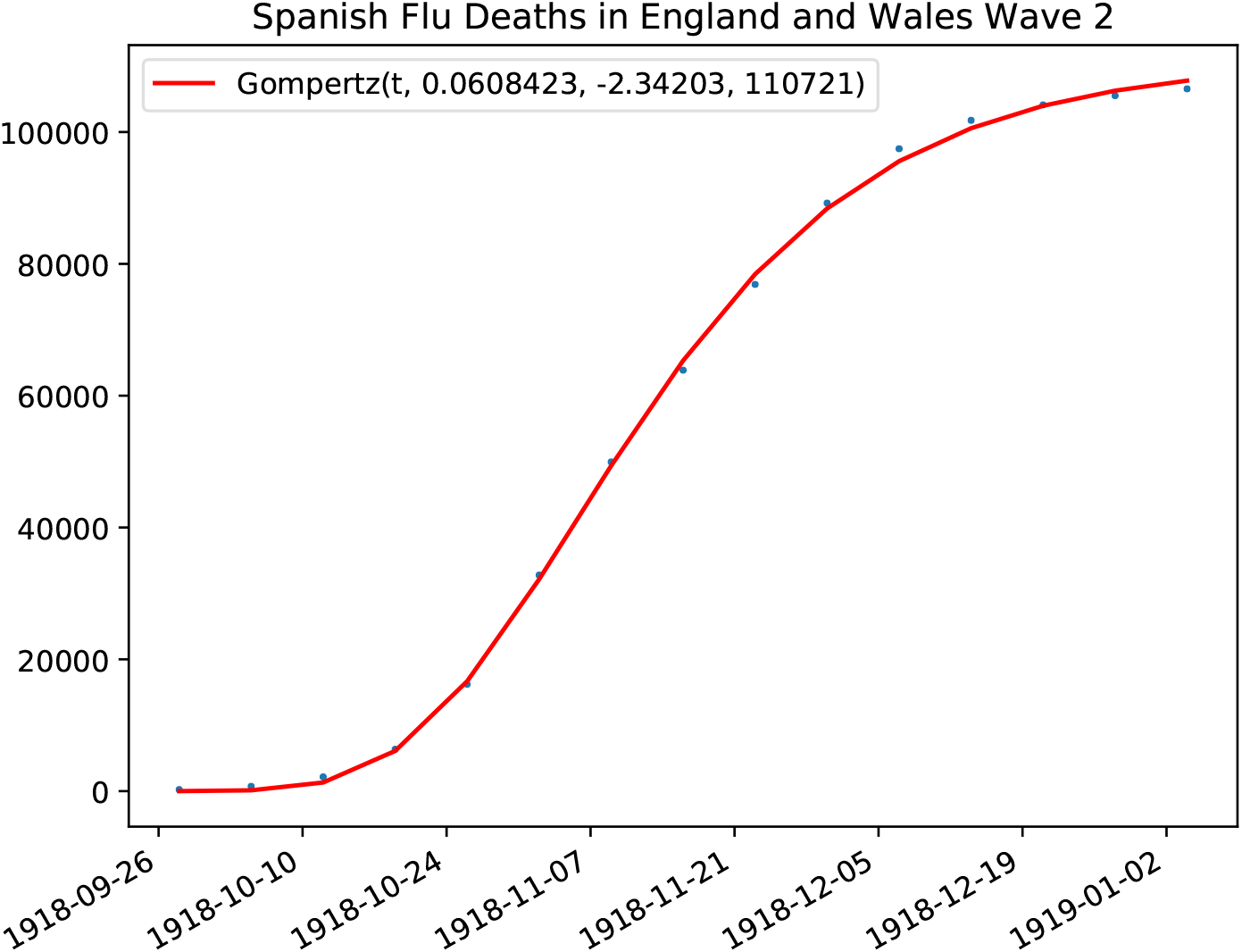
Spanish Flu deaths in England and Wales 28 Sep 1918 to 4 Jan 1919.

**Figure 73:**
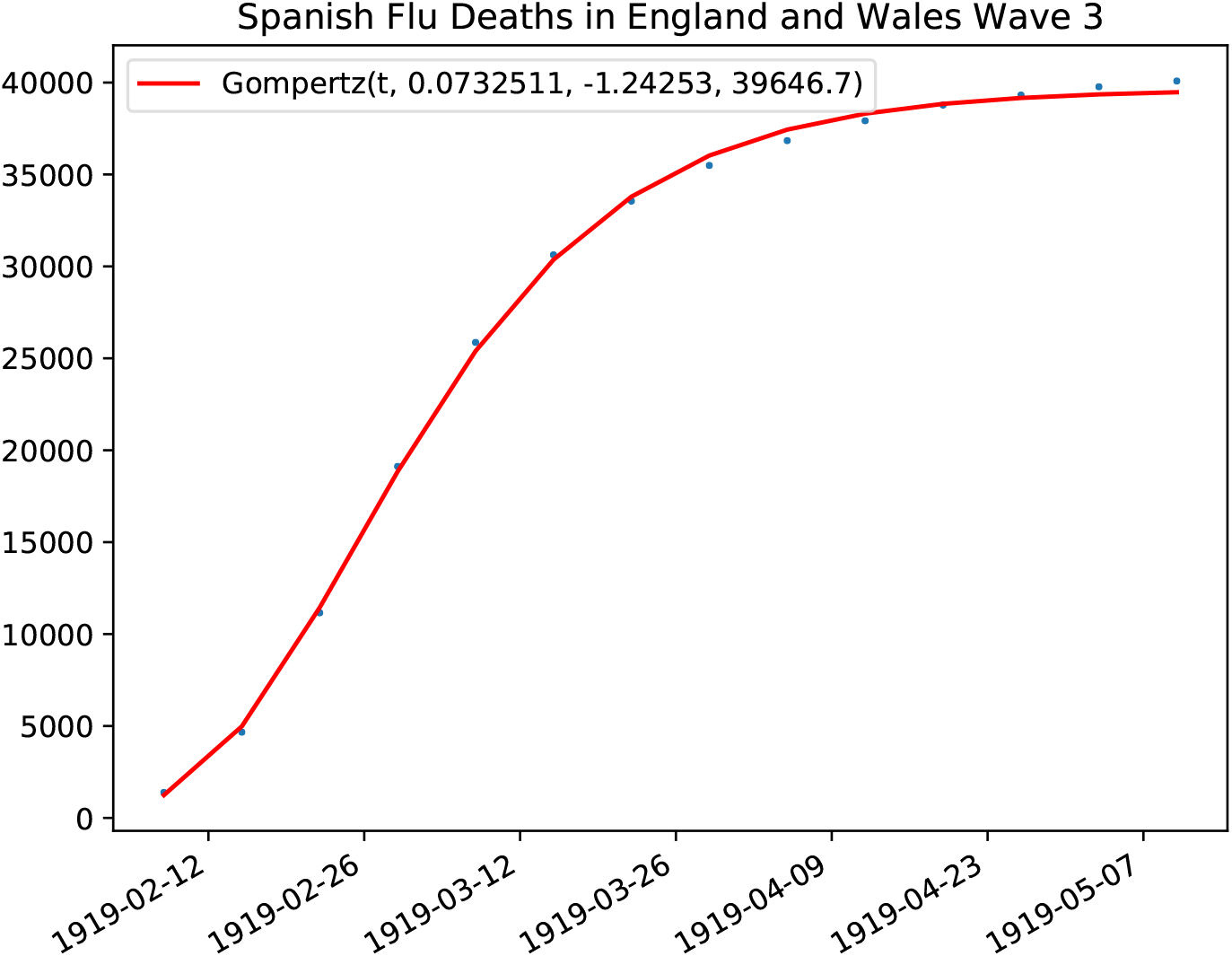
Spanish Flu deaths in England and Wales 8 Feb to 10 May 1919.

The cumulative deaths over all three waves are shown in Figure 74 with the Gompertz Function growth periods in red.

**Figure 74:**
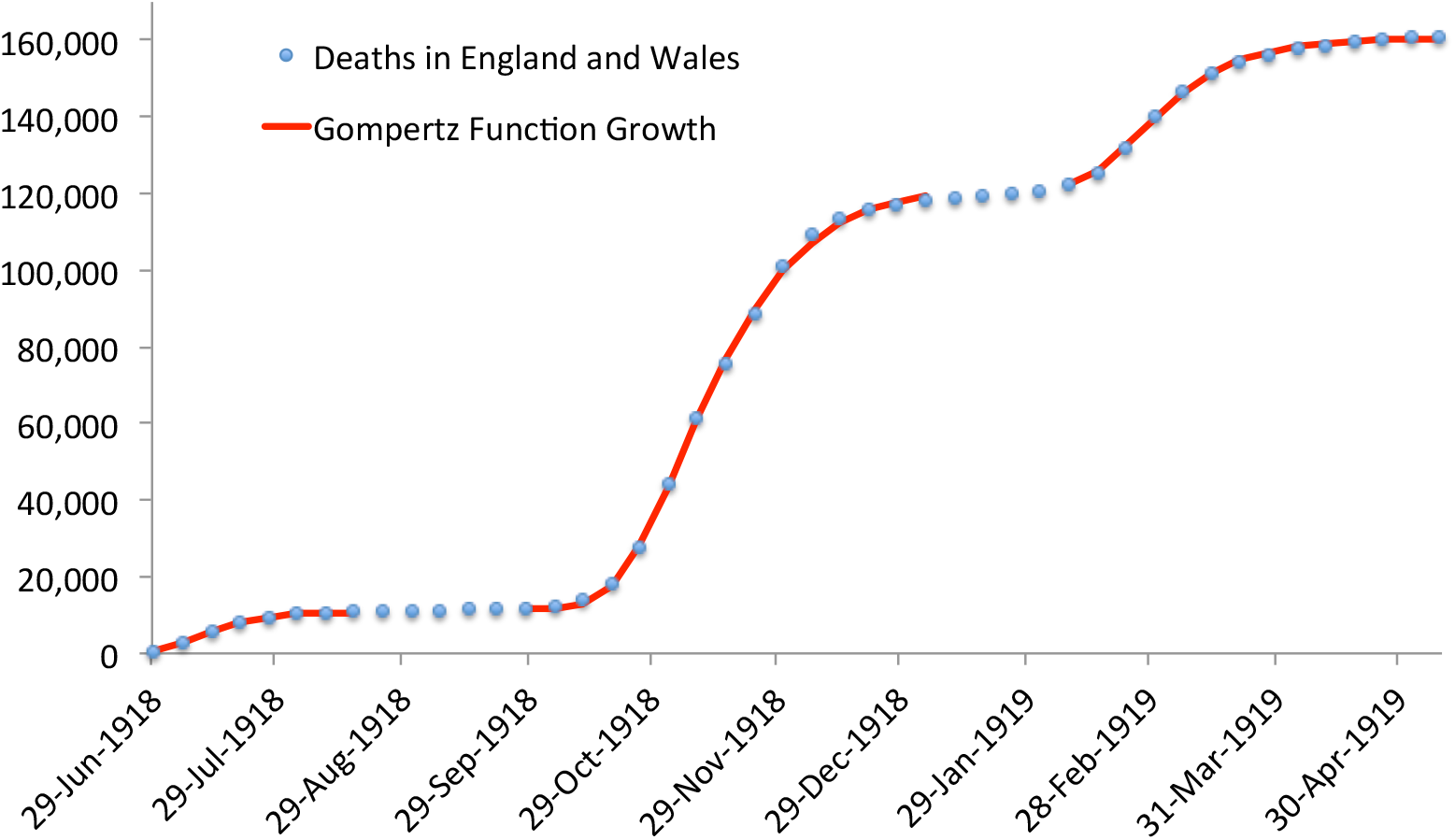
Three waves of Gompertz Function growth separated by linear growth in the ‘Spanish Flu’ Epidemic in England and Wales.

In linear growth periods, as we have already noted, the Reproduction Number must be very close to 1–which indicates that at the end of Waves 1 and 2 the dominant variant had become endemic. There appears to be no generally accepted mechanism that would explain the waves observed in the Spanish Flu,[20],[7] however the linear growth periods suggest that Waves 2 and 3 may have been driven by variants of the original virus. This is also consistent with increased virulence of the second wave relative to either Wave 1 or Wave 3.

In Figure 8 we illustrated the fit of the Gompertz Function to Spanish Flu deaths in Prussia from the beginning of the outbreak until 15 December 1918. From that date, cumulative deaths grew *linearly* as Figure 75 shows. The linear fit to the data shown is almost exact, with a maximum absolute error of only 0.23% and *r*^2^ > 0.99. We conclude from this that by mid-December of 2018 the Prussian Spanish Flu epidemic phase was over.

**Figure 75:**
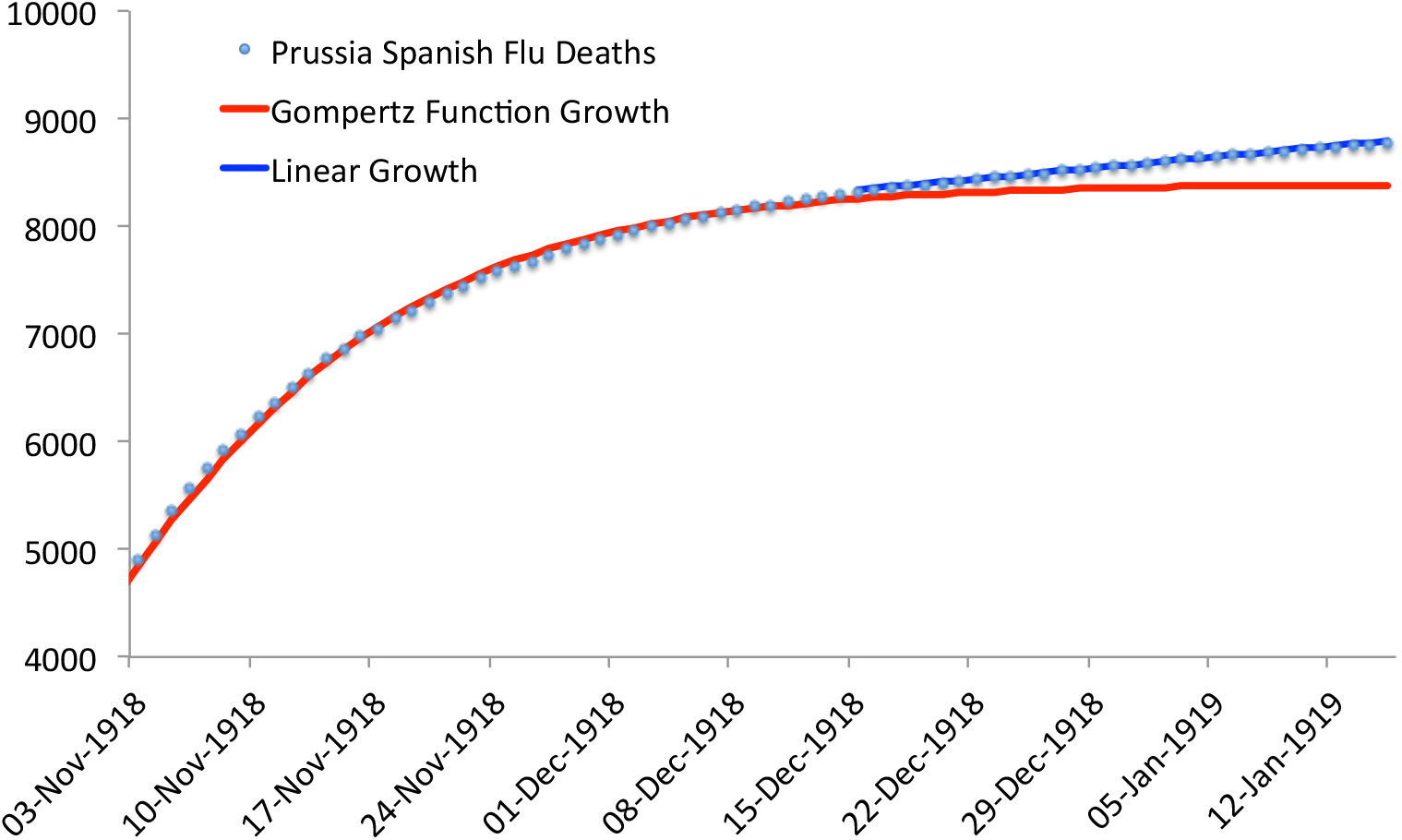
Transition from Gompertz Function growth to linear growth in cumulative Spanish Flu deaths in Prussia.

### 4.3 Covid-19: Endemic Cycles

We have already noted that the initial Covid-19 outbreaks in our examples all ended with a transition from Gompertz Function growth to linear growth.

Figure 51 showed this for Covid-19 Hospitalisations in London. After the linear exit from Gompertz Function growth phase in London Hospitalisations, there was piecewise linear growth with progressively lower slopes through the summer of 2020.

But, exactly as in Hope-Simpson’s influenza cycle, in September 2020 Covid-19 hospitalisations started rising again in Gompertz Function growth.(Figure 76)

**Figure 76:**
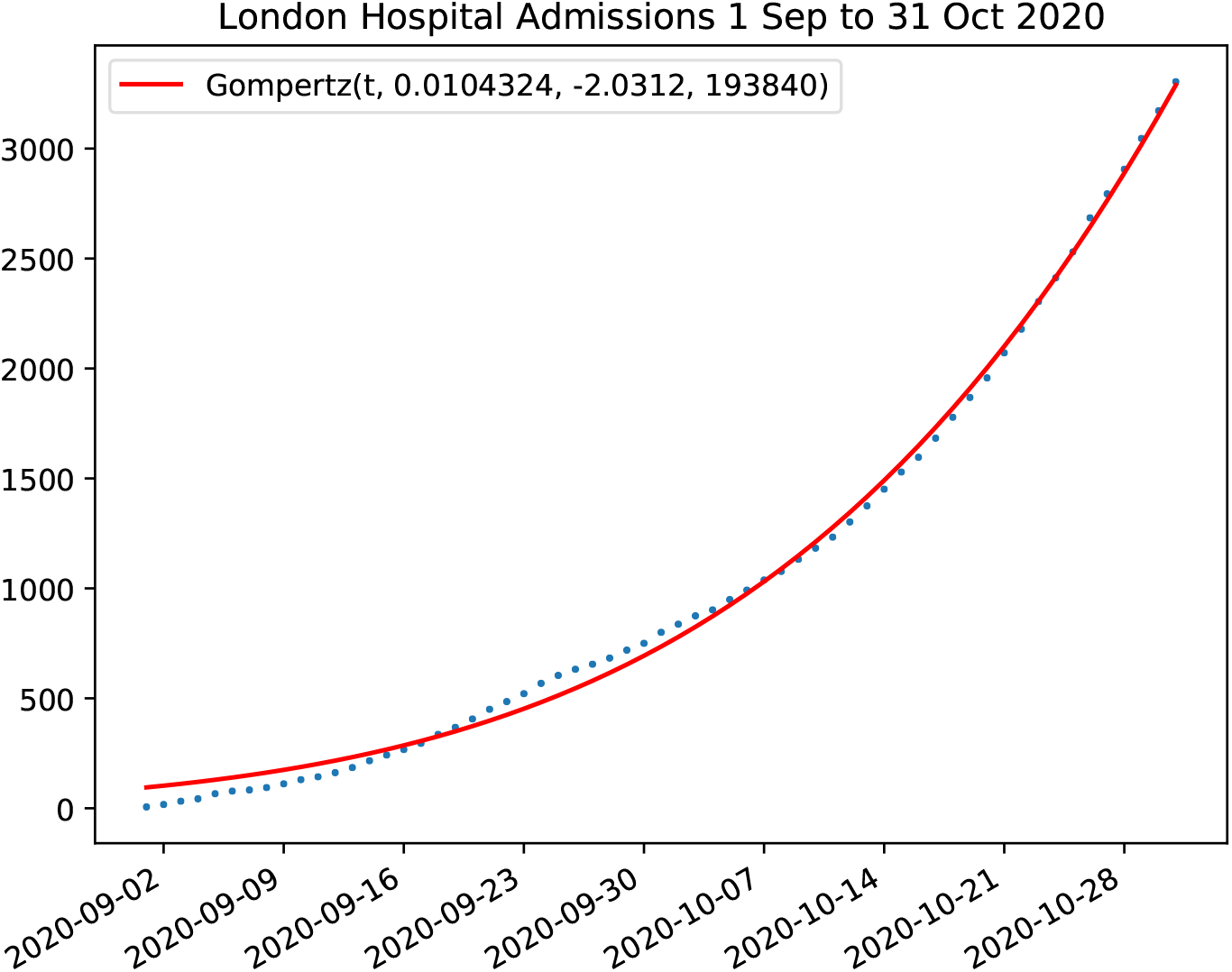
Gompertz Function growth London Hospitalisations 1 Sep to 31 Oct 2020.

This was followed by linear growth from 1 November to 7 December as Figure 77 illustrates.

**Figure 77:**
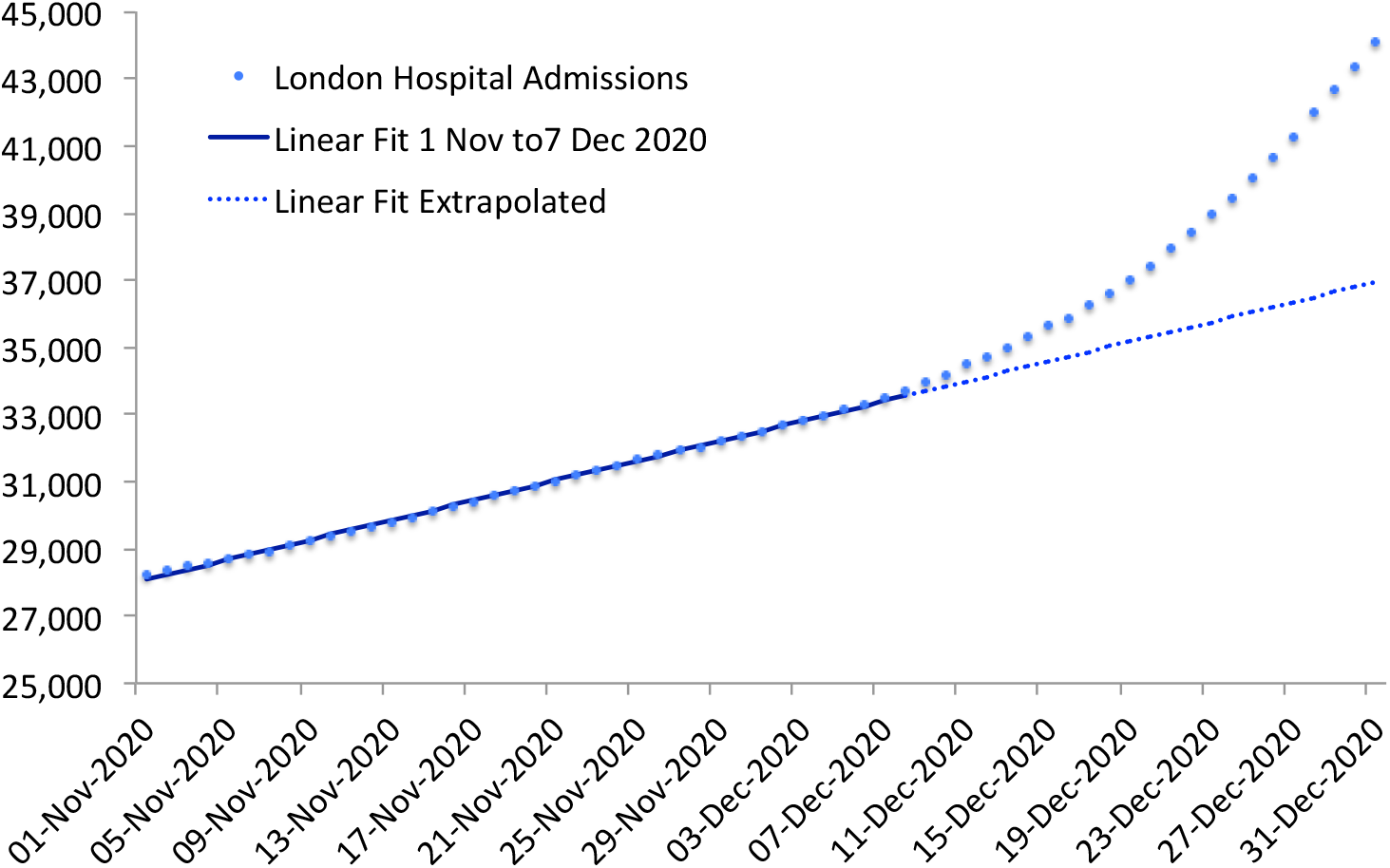
Transition from linear growth 1 Nov to 7 Dec 2020.

Then the main outbreak came at the height of the annual respiratory virus season (Figure 78). It ended with a transition to linear growth in March 2021. Covid-19 hospitalisations, ICU admissions and deaths in London repeated exactly the seasonal cycle of alternating Gompertz Function growth and linear growth we observed in Portuguese influenza cases.

**Figure 78:**
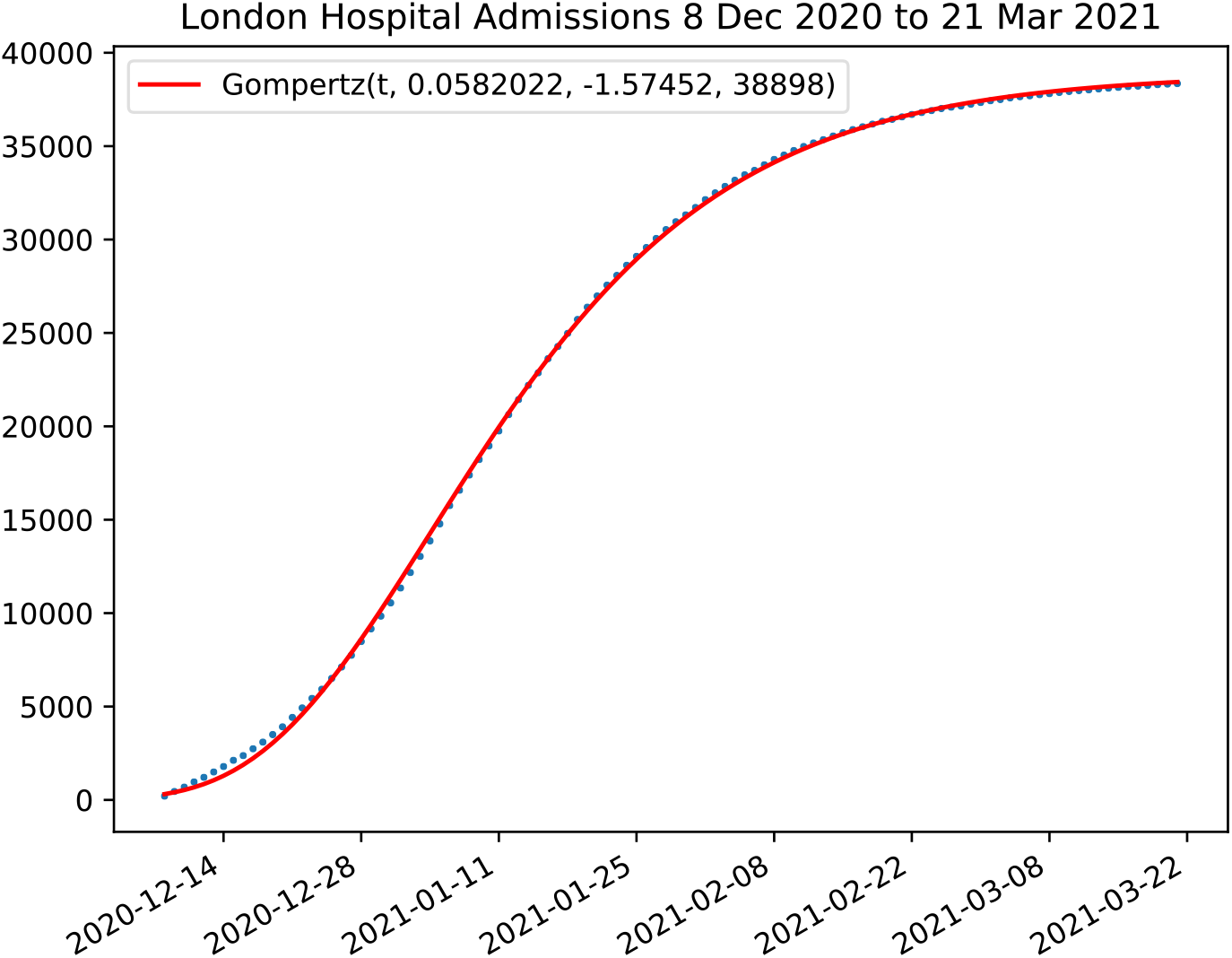
Covid-19 Hospital Admissions in London 8 Dec 2020-21 Mar 2021.

We have seen the same phenomenon repeated in multiple locations. We illustrate this next for Portugal’s ICU admissions. Like London, Portugal repeated the same pattern of linear and Gompertz function growth that we observed in the Portuguese influenza case records. Figure 79 shows the late summer outbreak of Gompertz Function growth followed by the main outbreak from January to March 2021 (Figure 80). This ended, as usual, with a transition to linear growth.

**Figure 79:**
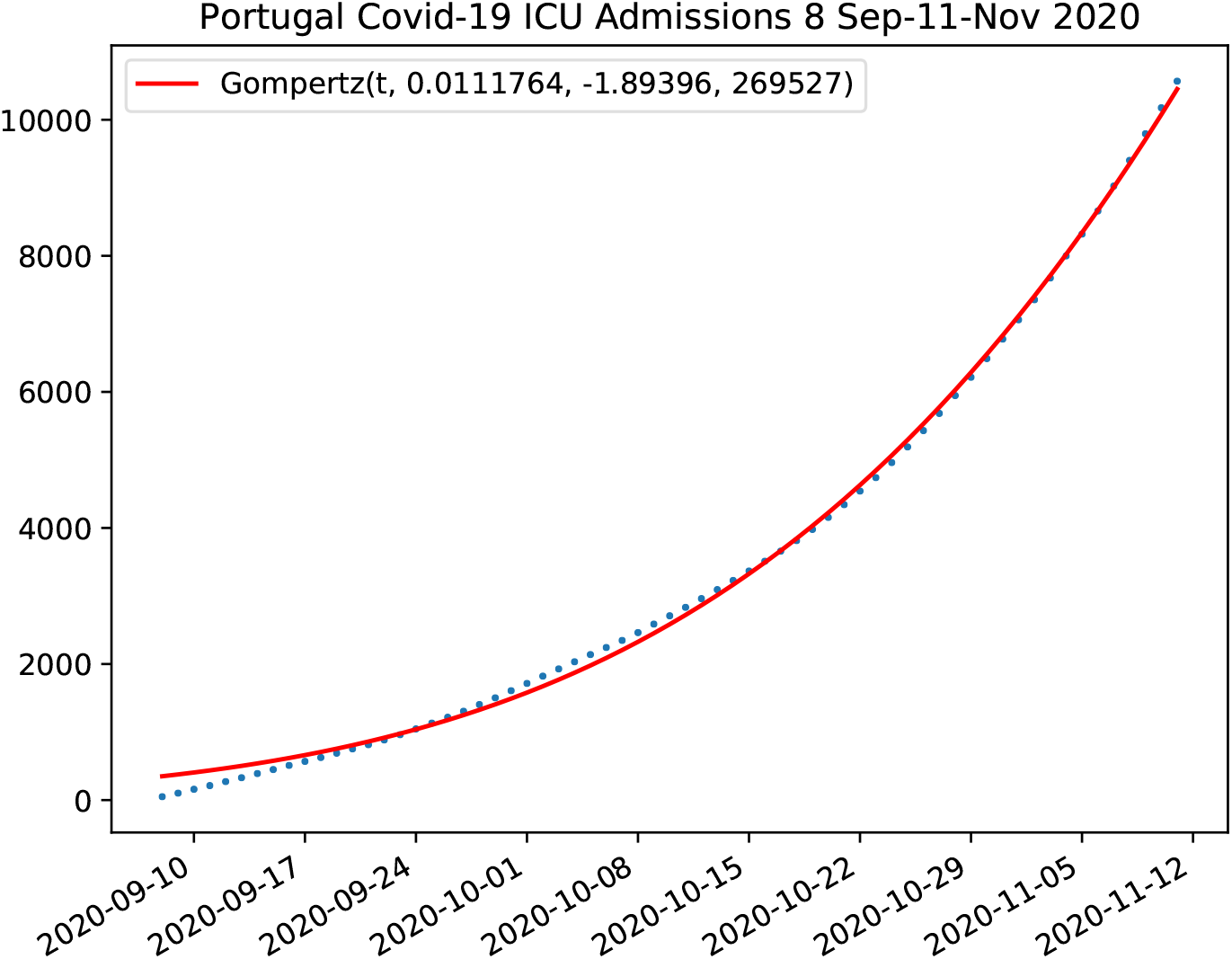
Portuguese Covid-19 ICU Admissions 8 Sep to 11 Nov 2020.

**Figure 80:**
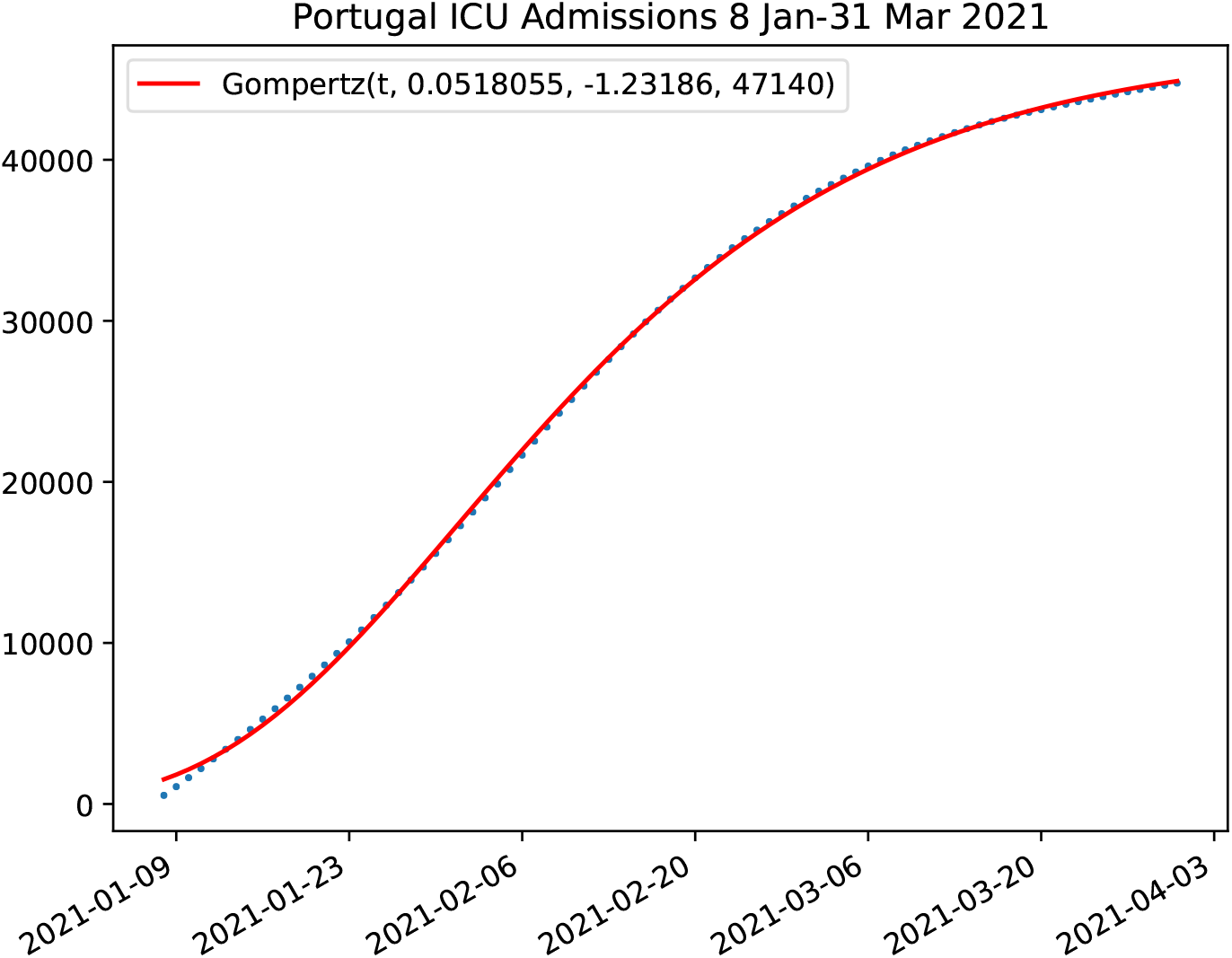
Portuguese Covid-19 ICU Admissions 8 Jan to 31 Mar 2021.

**Figure 81:**
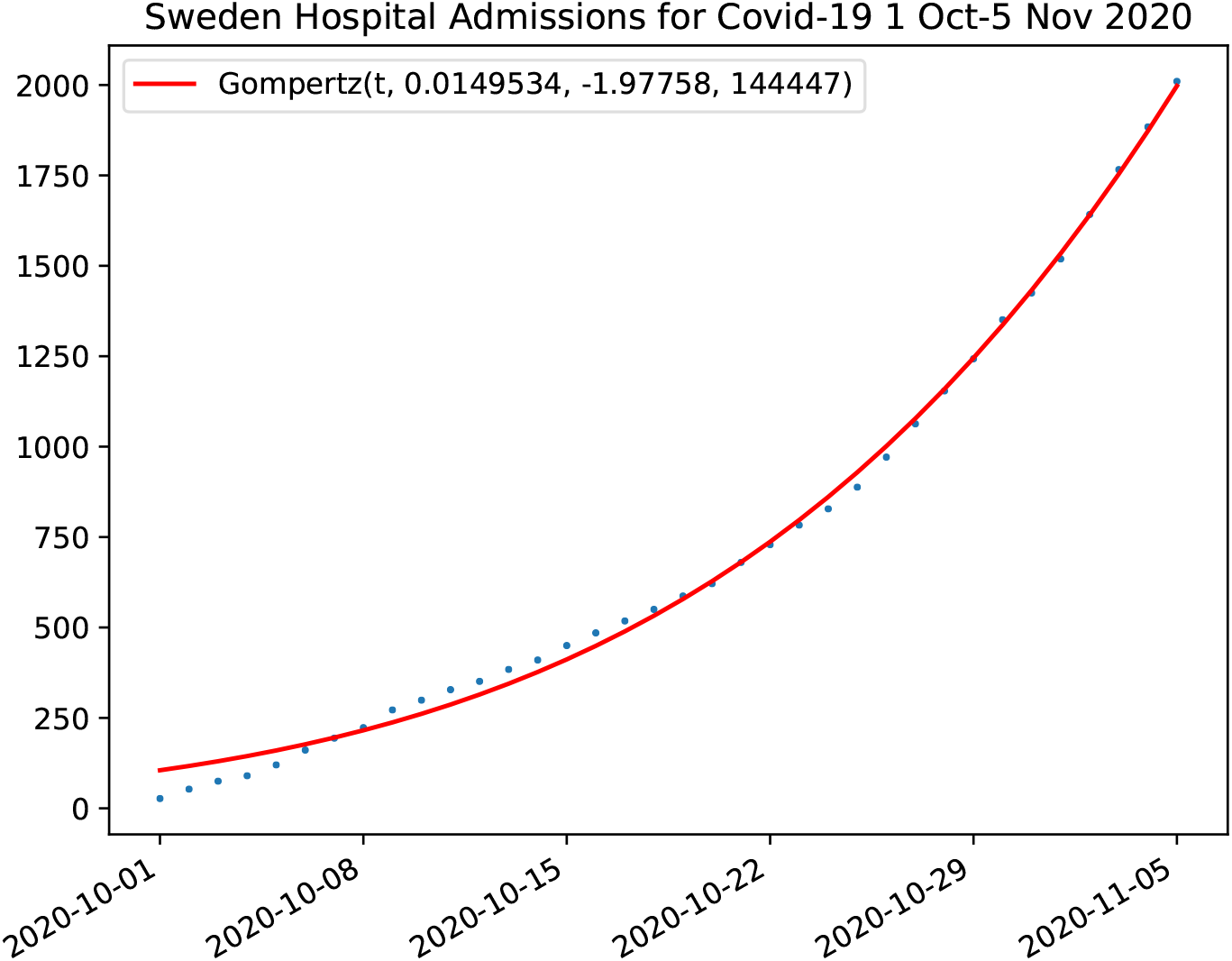
Sweden Covid-19 Hospital Admissions 1 Oct to 5 Nov 2020.

**Figure 82:**
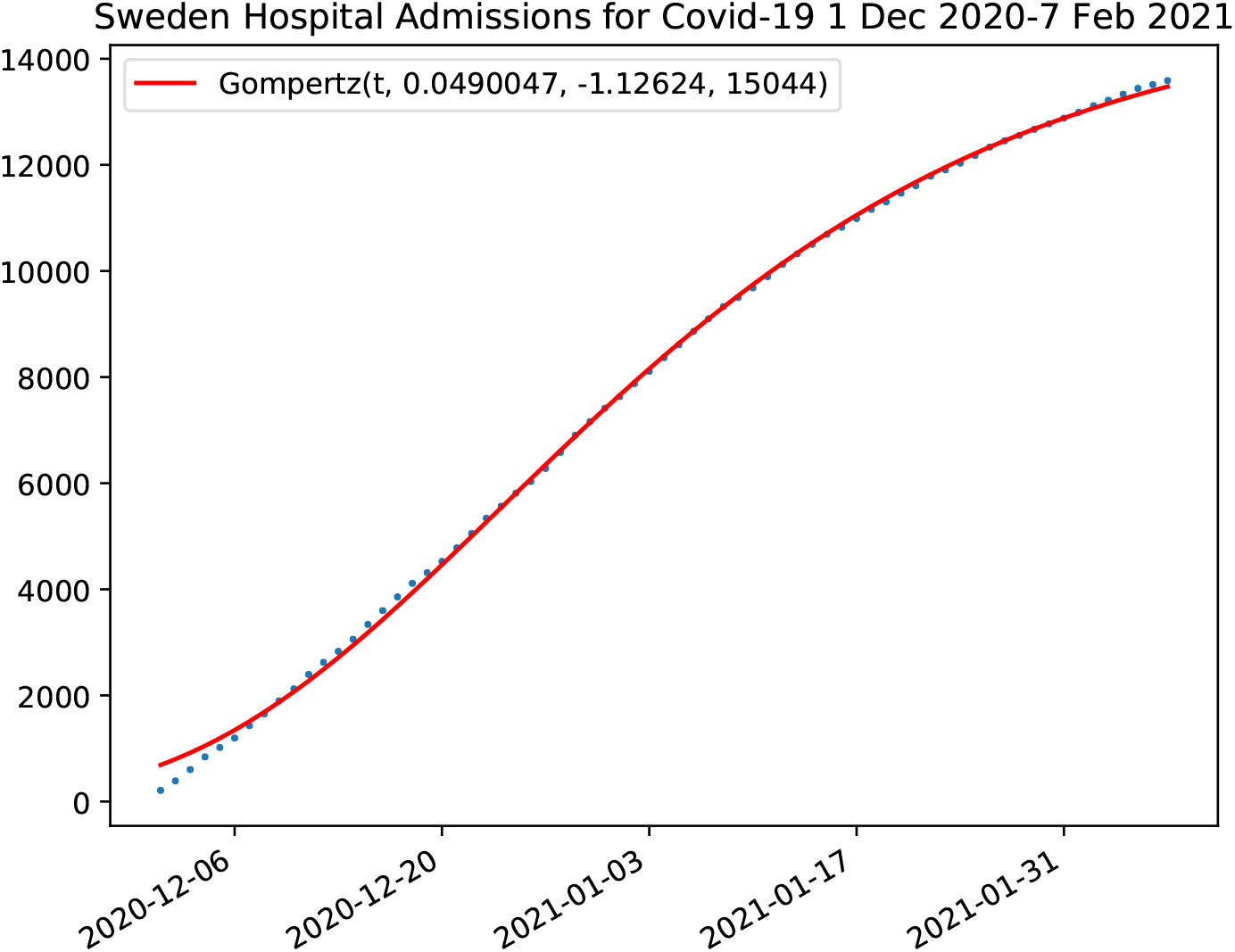
Sweden Covid-19 Hospital Admissions 1 Dec 2020 to 7 Feb 2021.

**Figure 83:**
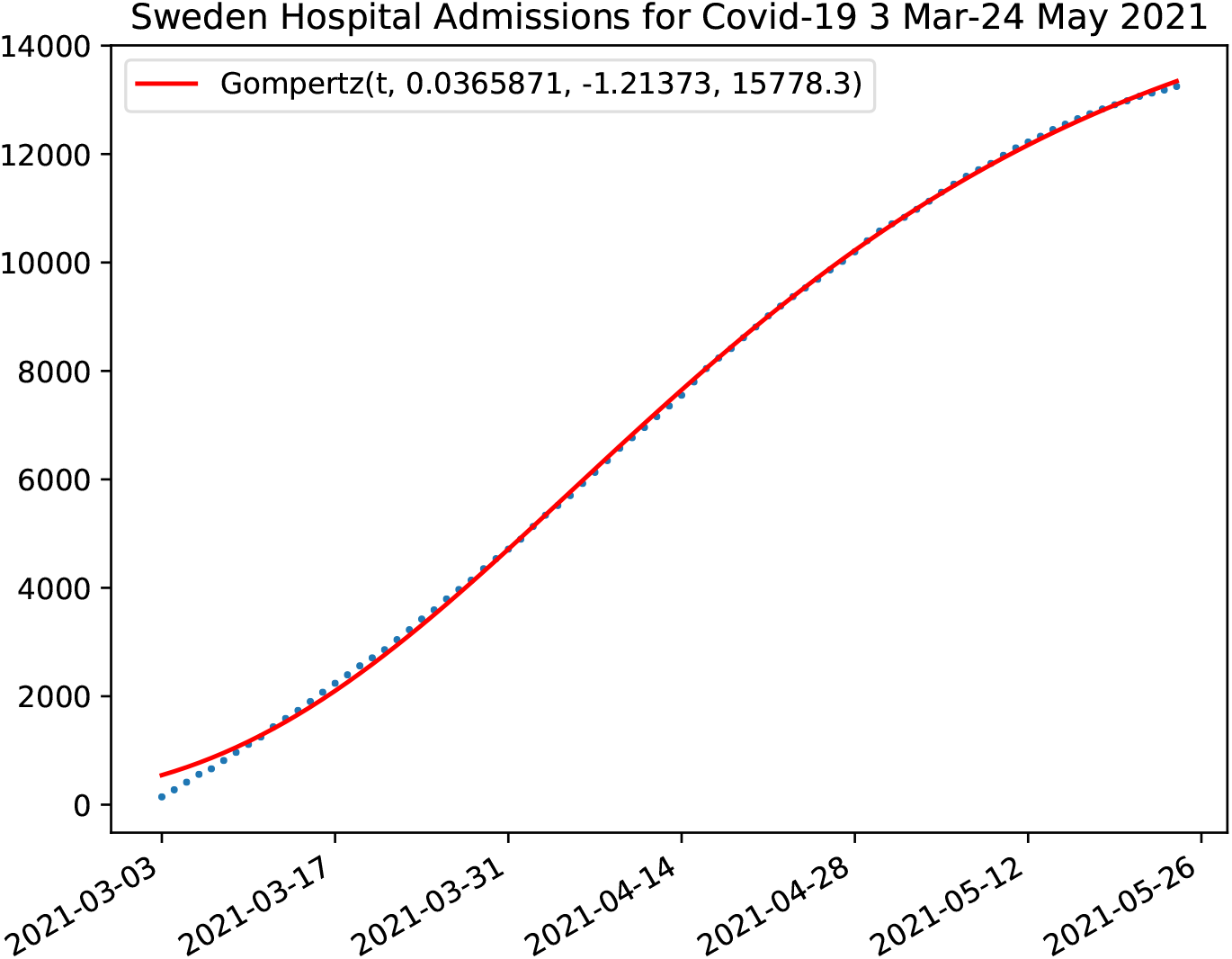
Sweden Covid-19 Hospital Admissions 3 Mar to 24 May 2021.

**Figure 84:**
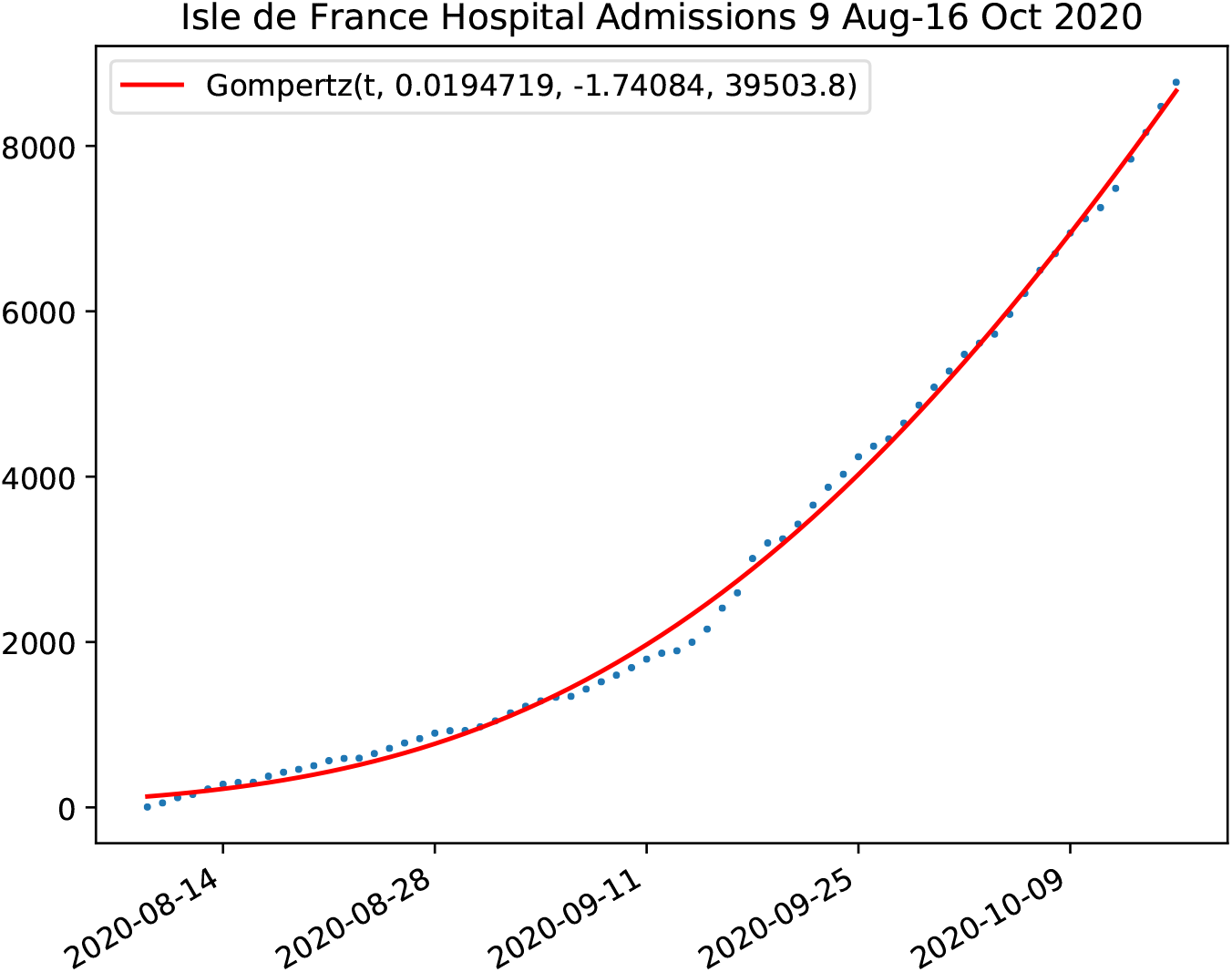
Isle de France Covid-19 Hospital Admissions 9 Aug to 16 Oct 2020.

**Figure 85:**
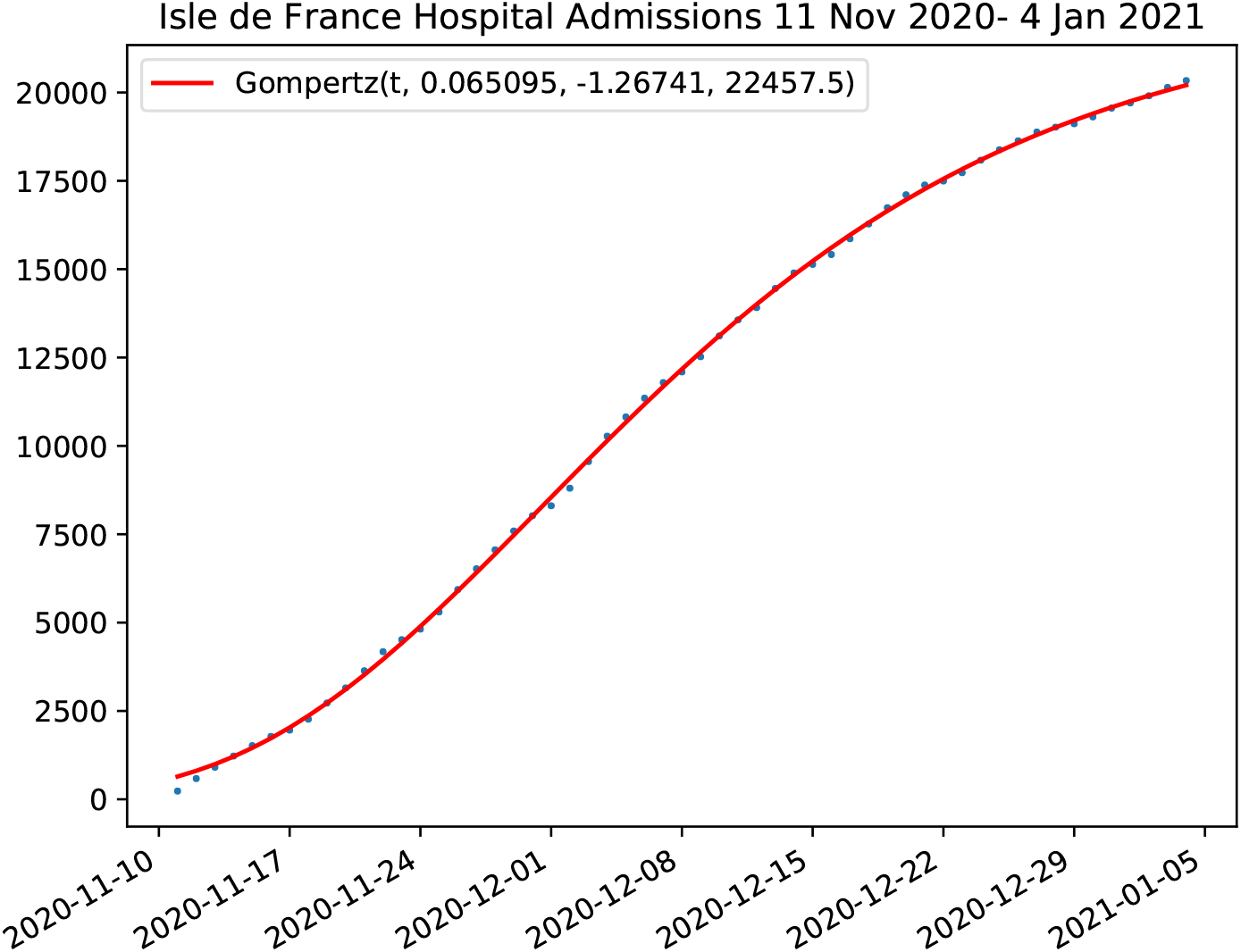
Isle de France Hospital Admissions 11 Nov 2020 to 4 Jan 2021.

**Figure 86:**
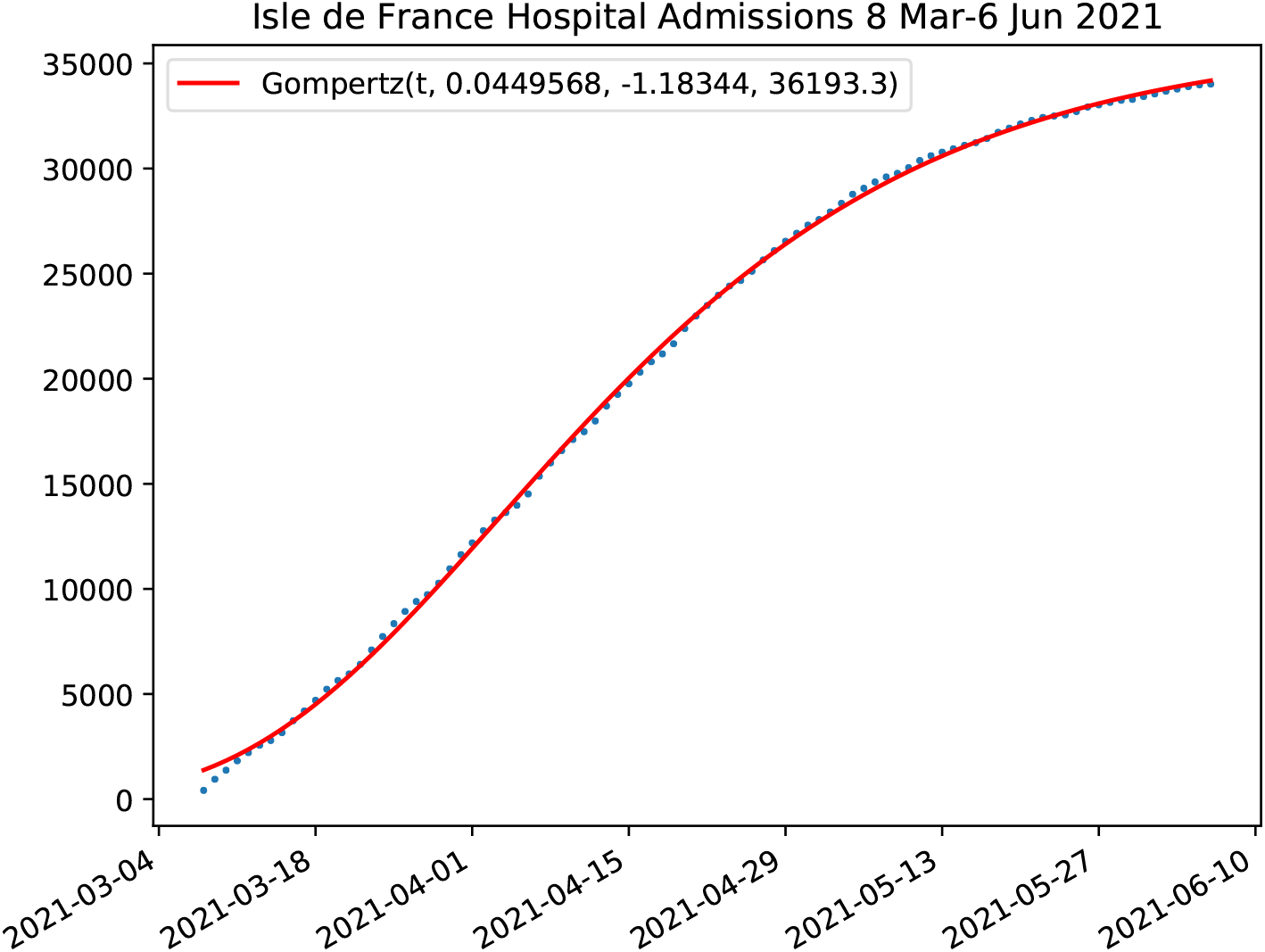
Isle de France Hospital Admissions 8 Mar 2021 to 6 Jun 2021.

**Figure 87:**
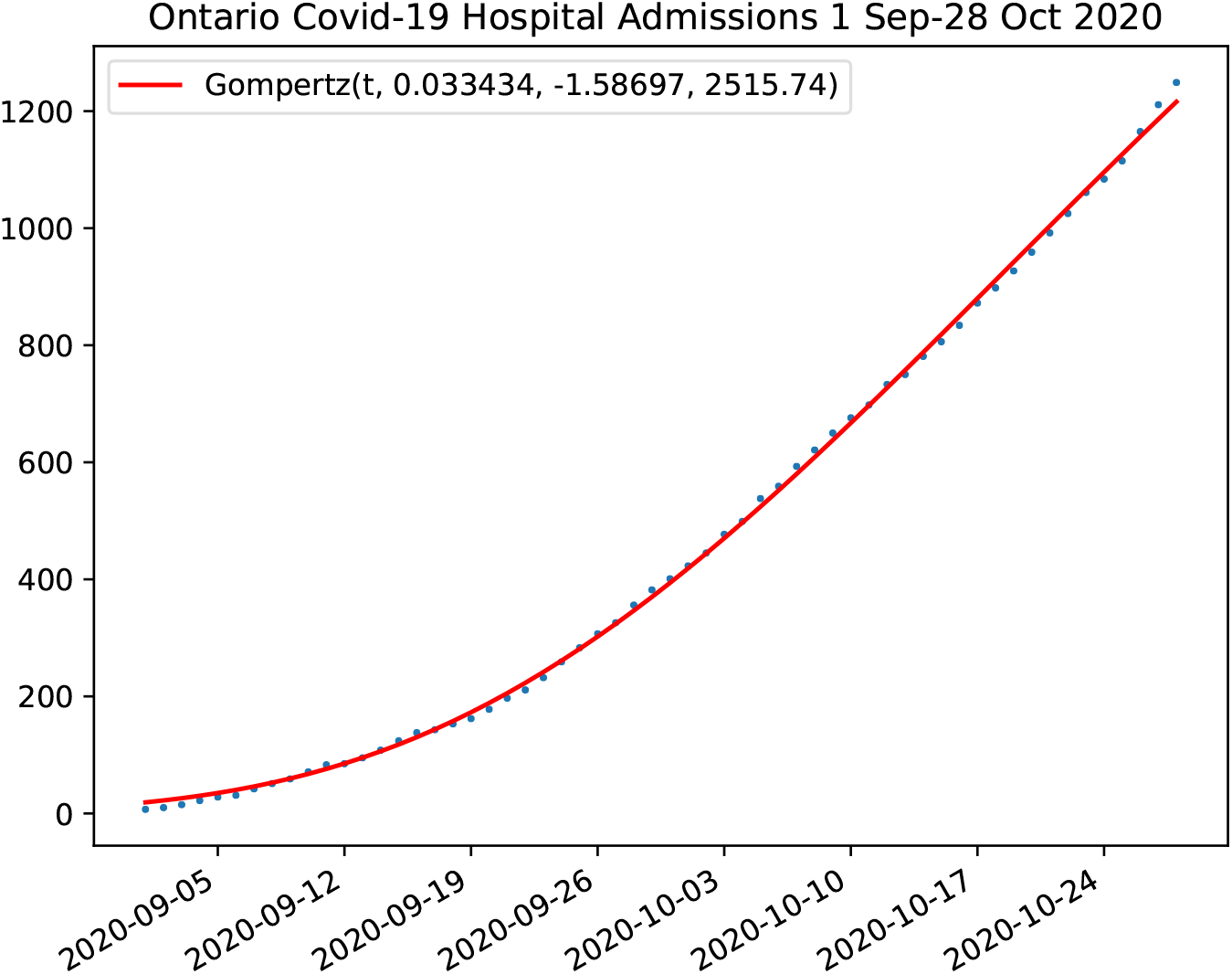
Ontario Covid-19 Hospital Admissions 1 Sep to 28 Oct 2020.

**Figure 88:**
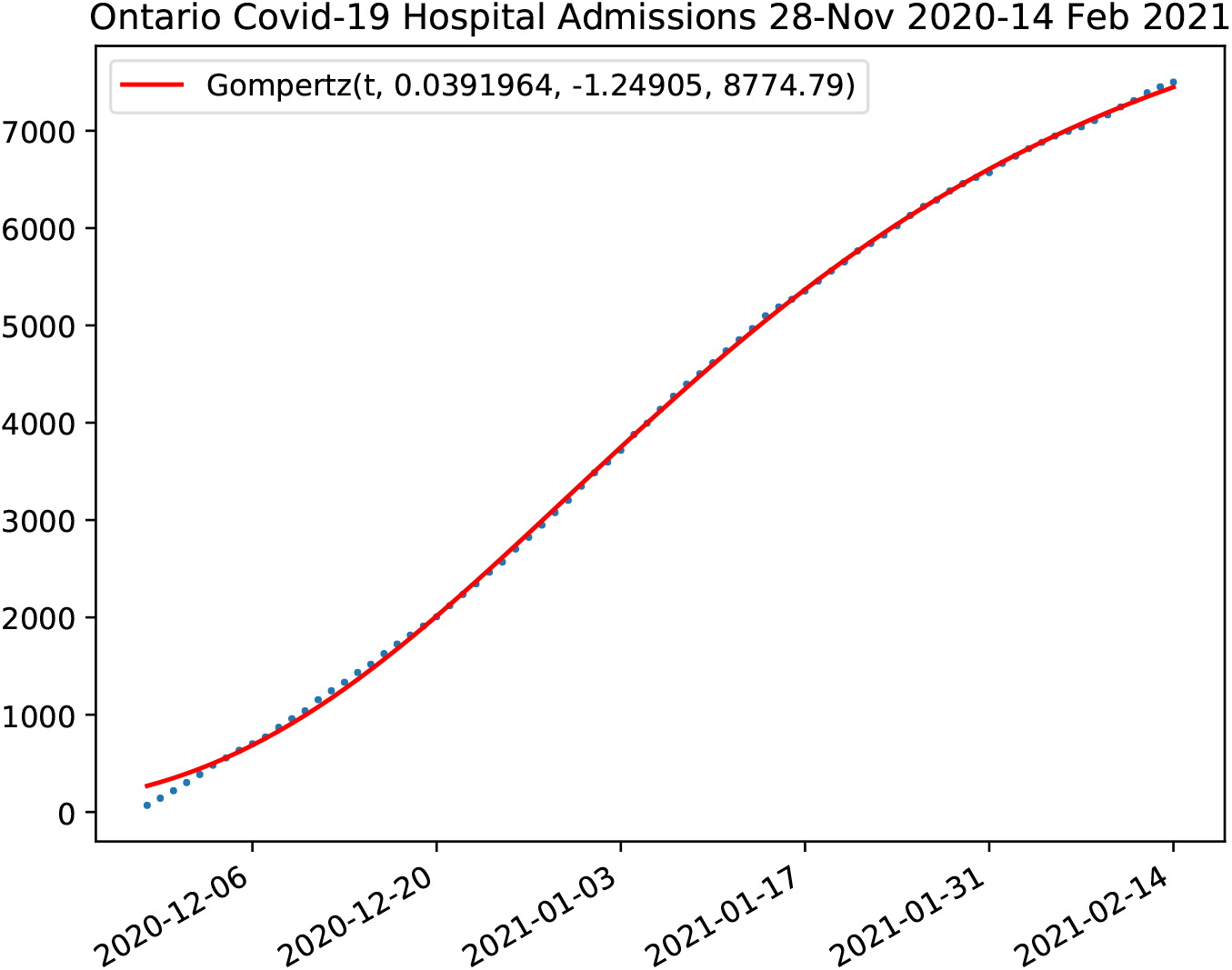
Ontario Covid-19 Hospital Admissions 28 Nov 2020 to 14 Feb 2021.

**Figure 89:**
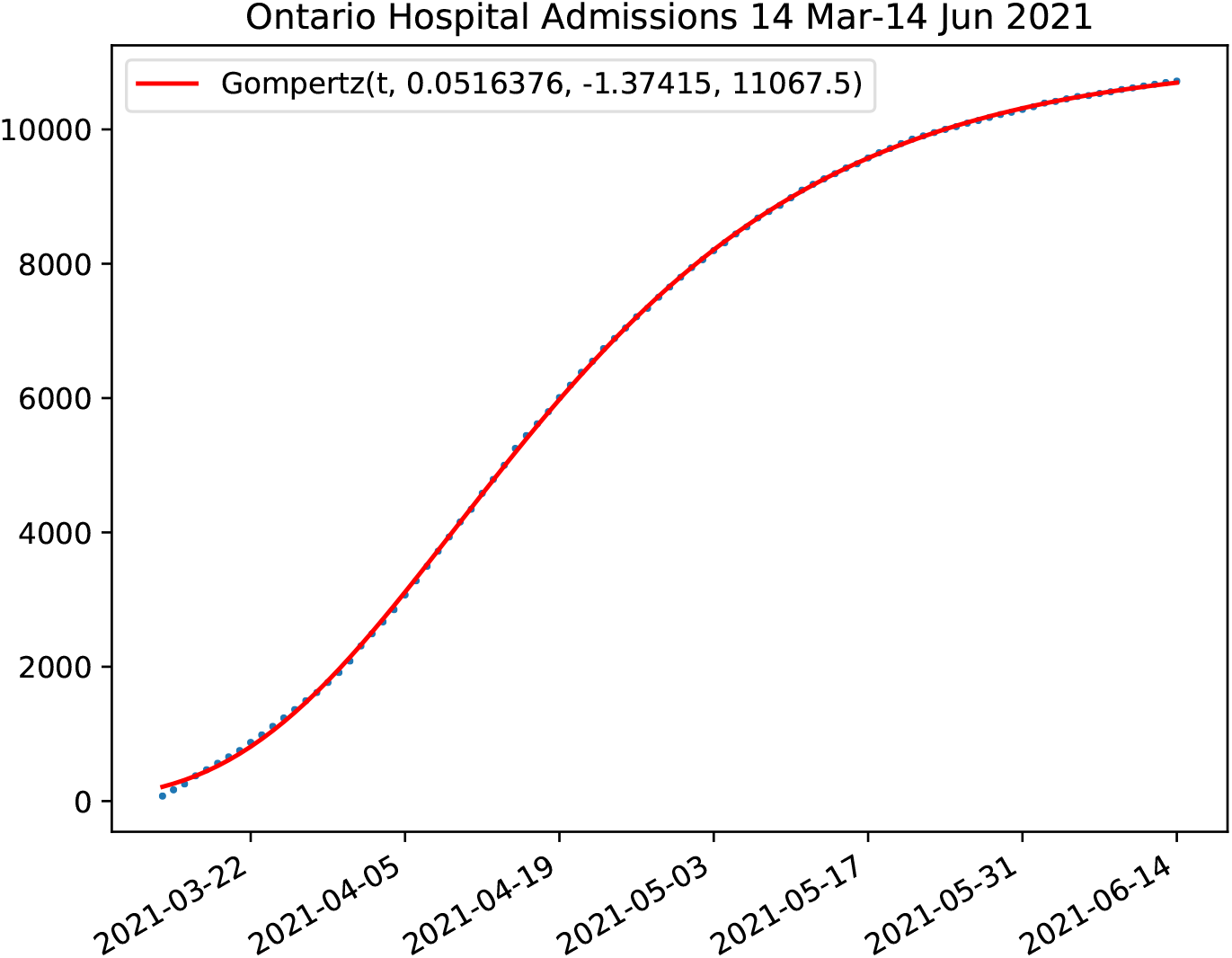
Ontario Covid-19 Hospital Admissions 14 Mar to 14 Jun 2021.

In data from Sweden, Isle de France and in Ontario, once again we find this influenza-like alternation of linear and Gompertz Function growth in the late summer and the autumn of 2020.

In each of these three locations there was an outbreak of Gompertz Function growth in the usual winter period, ending as usual with a period of linear growth.

But then, unlike London and Portugal, there was a further ‘wave’ beginning in March (late, but still within the seasonal period identified by Hope-Simpson). The two waves were separated by linear growth–which indicates that a different variant may have been responsible for the second one.

In Rio de Janeiro and Amazonas we have also seen the alternation of Gompertz Function growth and linear growth. The tropical region influenza cycle is more complicated than in the temperate zone so a comparison of the timing of Covid-19 outbreaks and the annual influenza cycle will be explored elsewhere. Figures 90-93 for hospitalisations in the State of Rio de Janeiro and deaths in Amazonas show how well the Gompertz Function fits the second round of Covid-19 outbreaks.

**Figure 90:**
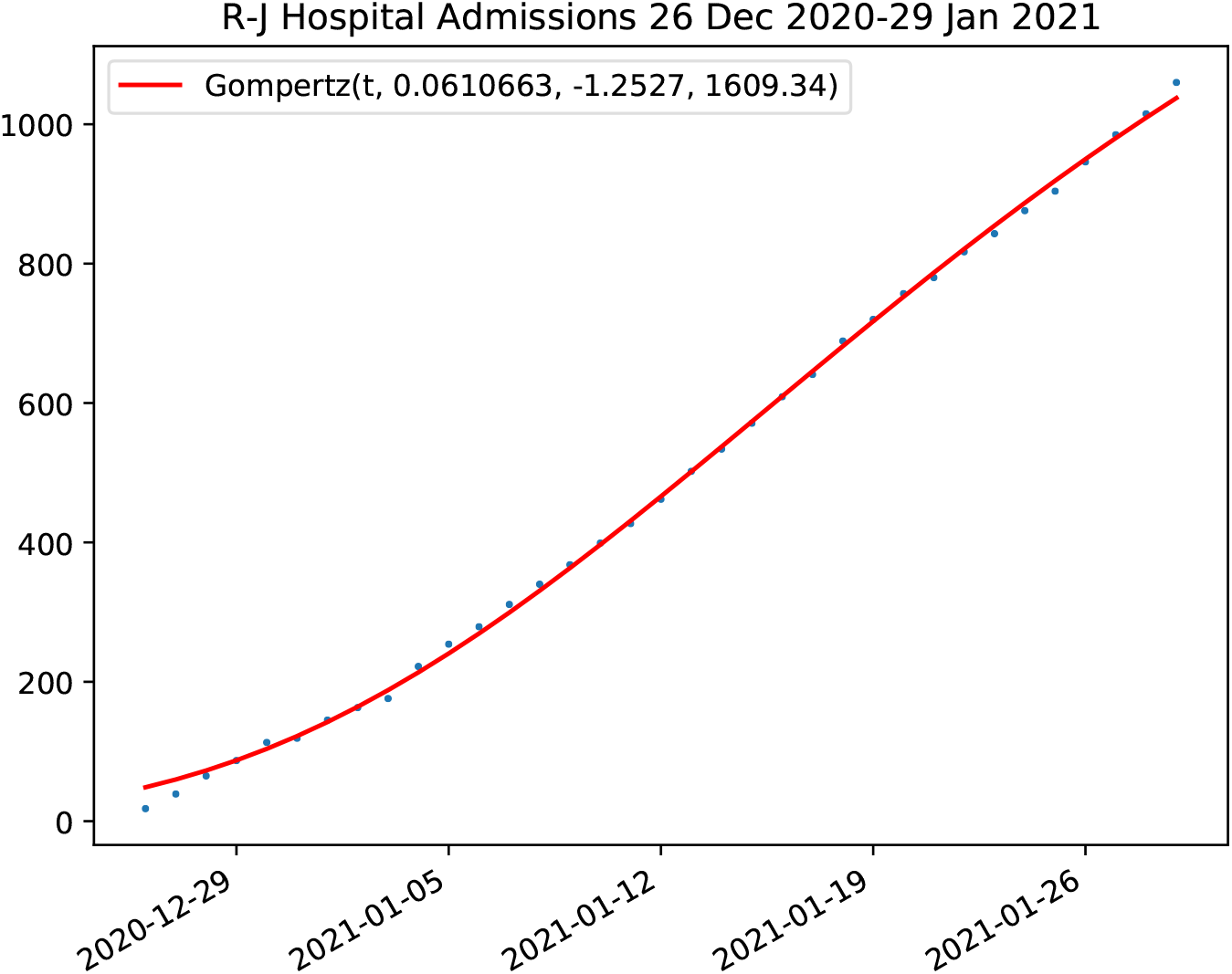
RJ Covid-19 Hospital Admissions 26 Dec 2020 to 29 Jan 2021.

**Figure 91:**
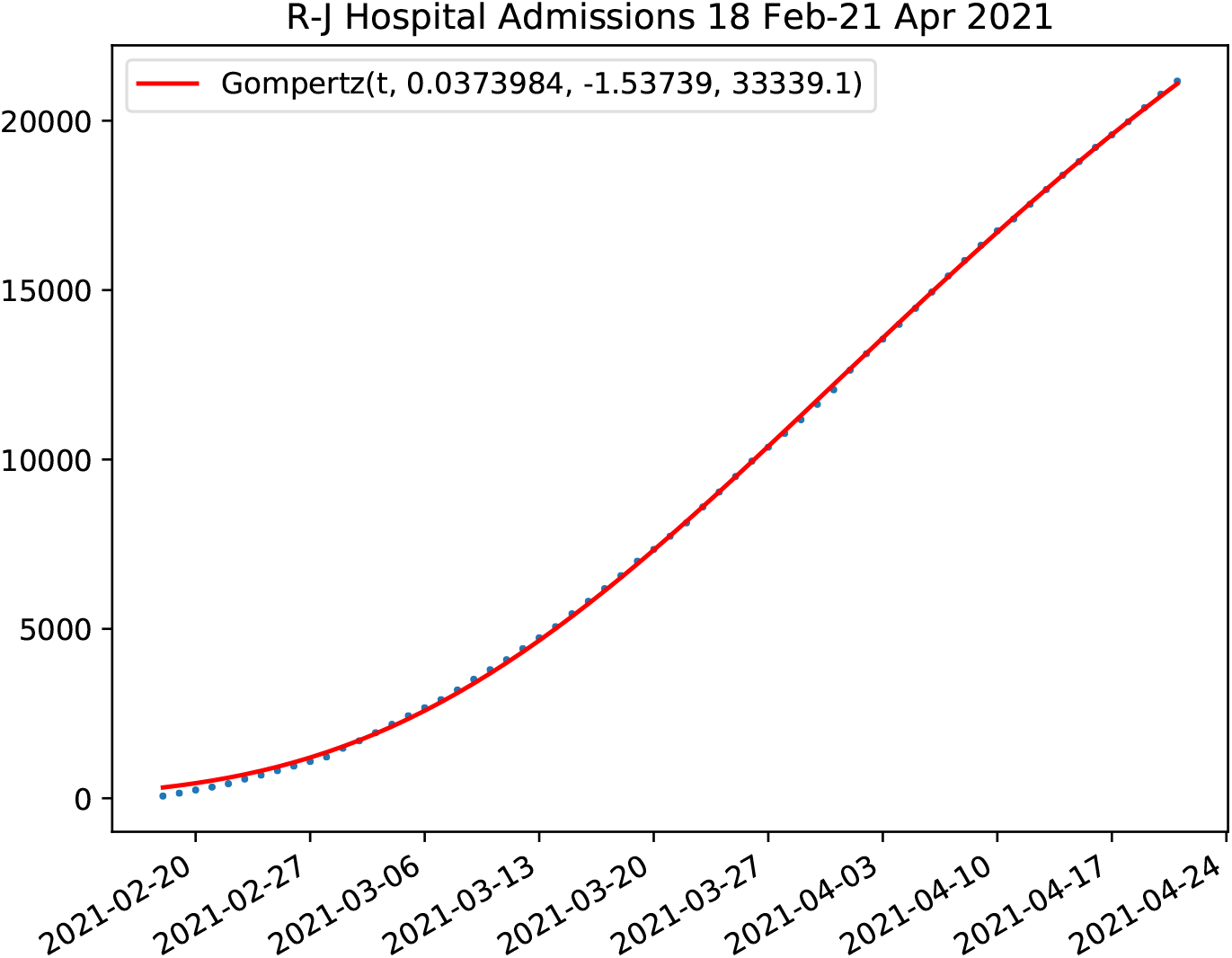
RJ Covid-19 Hospital Admissions 18 Feb to 21 Apr 2021.

**Figure 92:**
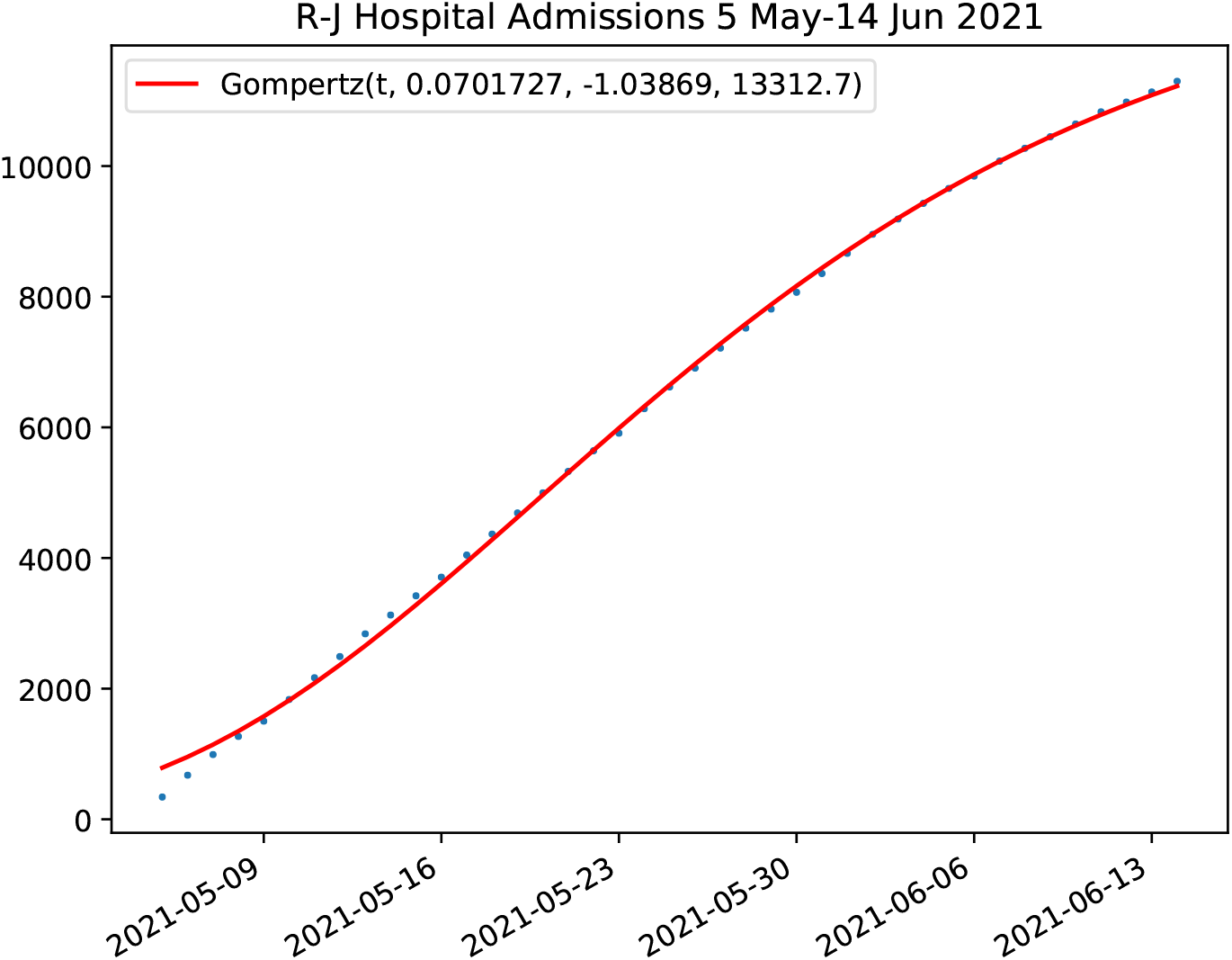
RJ Covid-19 Hospital Admissions 5 May to 14 Jun 2021.

**Figure 93:**
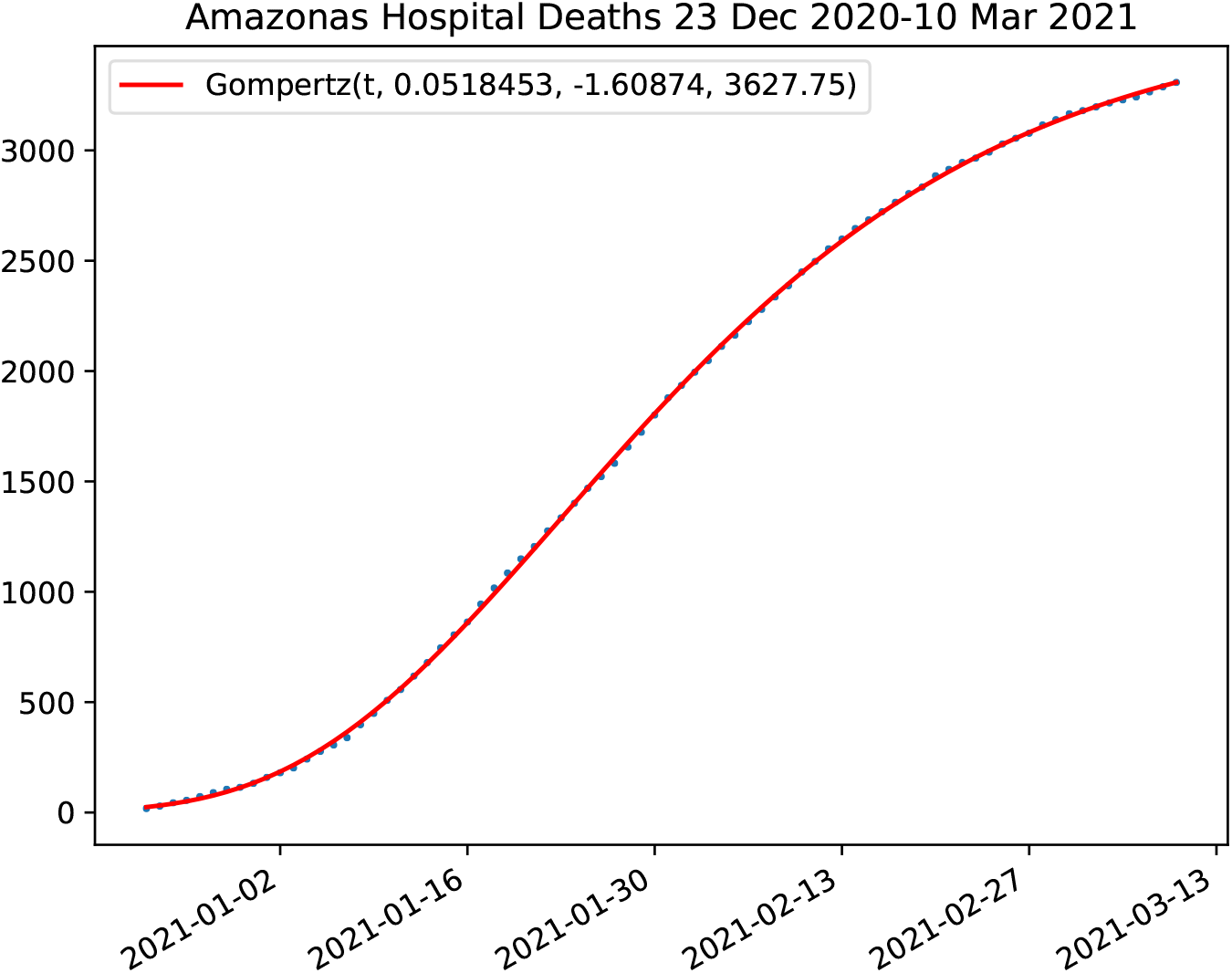
Amazonas Covid-19 Hospital Deaths 23 Dec 2020 to 10 Mar 2021.

Table 2 shows a sample of dates for alternating Gompertz Function and linear growth.

**Table 2:** Covid-19 Growth Cycle.

| Alternating Gompertz Function and Linear Growth for Covid-19 |  |  |
| --- | --- | --- |
| Area and Data type | Gompertz Growth | Linear Growth |
| Sweden Hosp | 3 Mar 20-17 May 20<br>1 Oct 20-5 Nov 20<br>1 Dec 20-7 Feb 21<br>19 Mar 21-24 May 21 | 18 May 20-30 Sep 20<br>6 Nov 20-30 Nov 20<br>8 Feb 21-18 Mar 21 |
| London Hosp | 19 Mar 20-23 Apr 20<br>1 Sep 20-31 Oct 20<br>8 Dec 20-21 Mar 21 | 24 Apr 20-31 Aug 20<br>1 Nov 20-7 Dec 20 |
| Isle de France Hosp | 18 Mar 20-11 May 20<br>9 Aug 20-16 Oct 20<br>11 Nov 20-4 Jan 21<br>8 Mar 21-6 Jun 21 | 12 May 20-8 Aug 20<br>17 Oct 20-10 Nov 20<br>5 Jan 21-7 Mar 21 |
| Ontario Hosp | 1 Mar 20-14 Jun 20<br>2 Sep 20-31 Oct 20<br>28 Nov 20-14 Feb 21<br>14 Mar 21-14 Jun 21 | 15 Jun 20-1 Sep 20<br>1 Nov 20-27 Nov 20<br>15 Feb 21-13 Mar 21 |
| Portugal ICU | 14 Mar 20-15 May 20<br>8 Sep 20-11 Nov 20<br>8 Jan 21-31 Mar 21 | 16 May 20-7 Sep 20<br>12 Nov 20-7 Jan 21 |
| RJ Hosp | 15 Mar 20-30 Jun 20<br>26 Dec 20-29 Jan 21<br>18 Feb 21-21 Apr 21<br>5 May 21-14 Jun 21 | 1 Jul 20-25 Dec 20<br>30 Jan 21-17 Feb 21<br>22 Apr 21-4 May 21 |
| Amazonas Deaths | 1 Apr 20 -14 Jun 20<br>23 Dec 20-10 Mar 21 | 15 Jun 20-22 Dec 20 |
| End of Table |  |  |

## 5 Notes on Gompertz Function Fits

### 5.1 The ‘Exponential Growth’ Phase

Early in the Covid-19 epidemic the doubling time was a matter of daily speculation and concern. Then, and subsequently, the phrase ‘exponential growth’ has been used to describe Covid-19 outbreaks by many people who clearly have only a tenuous grasp of its meaning.

Epidemics do indeed grow rapidly, even extremely rapidly, at first. Epidemiologists often make use of the idea that epidemics have an initial exponential growth phase, but this is really an approximate feature of compartmental models^24^ not an empirical observation. In a study using a range of data sets from real epidemics, Chowell et al [5] show that subexponential growth is the rule rather than the exception in the early phase of an epidemic.

The utility of the ‘exponential growth’ approximation is that it allows modellers estimate model inputs. While there is often a reasonably good fit to an exponential function in the early phase in epidemic data, such fits rapidly lose any predictive power. Exponential growth is simply too explosive to be sustainable.

Figures 94 and 95 show the Gompertz Function fit and the best exponential function fit (using the same nonlinear regression algorithm) for Covid-19 hospitalisations in Portugal from 5 March to 9 April 2020. It’s (just) possible to see from the graphs that the Gompertz Function fit is closer to the data. But in fact the Gompertz Function fit is *much* better: the mean squared error of the exponential fit is 1.8 times as large as it is for the Gompertz Function over the sample period.

**Figure 94:**
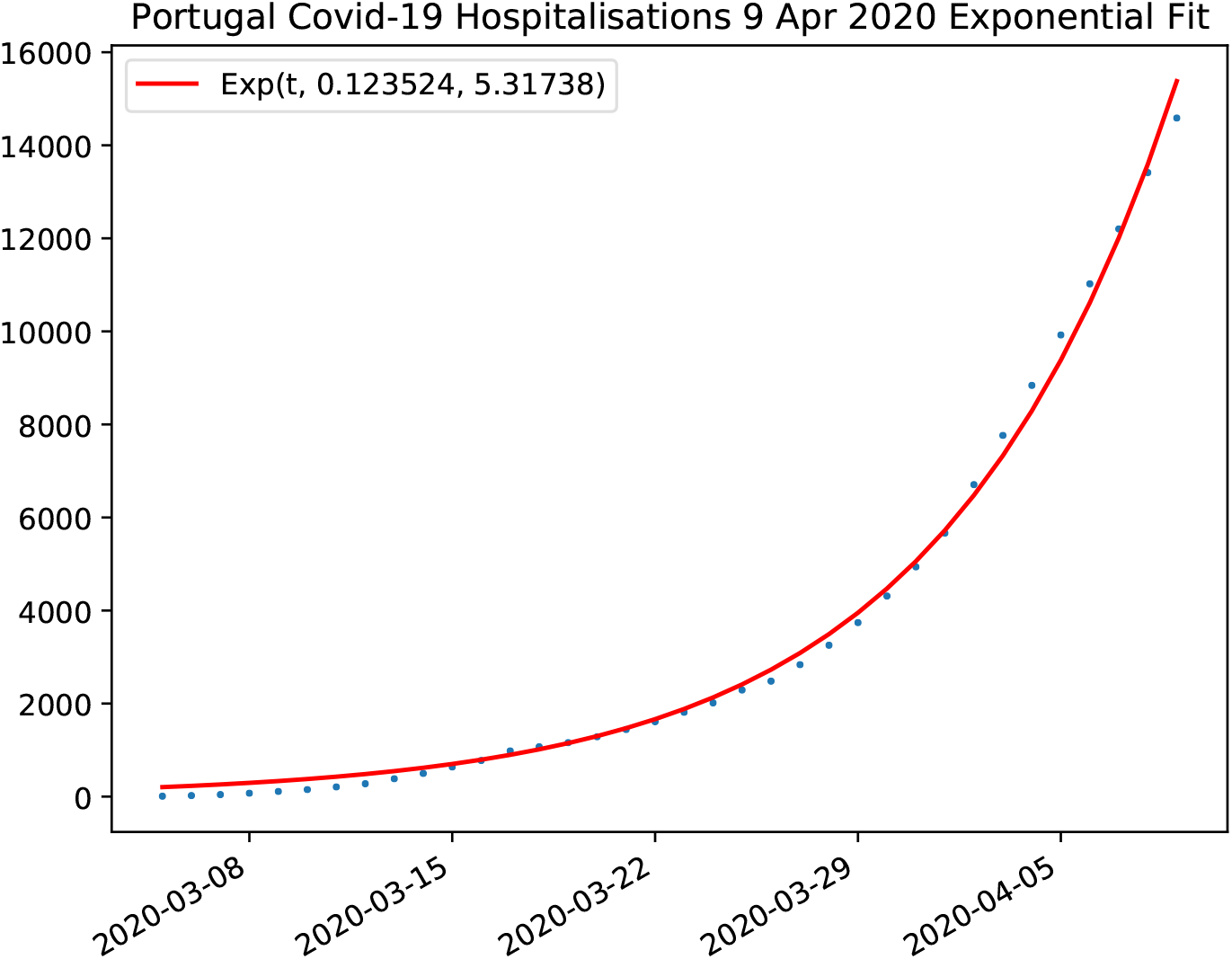
‘Exponential Growth’ in Portuguese Covid-19 Hospitalisations.

**Figure 95:**
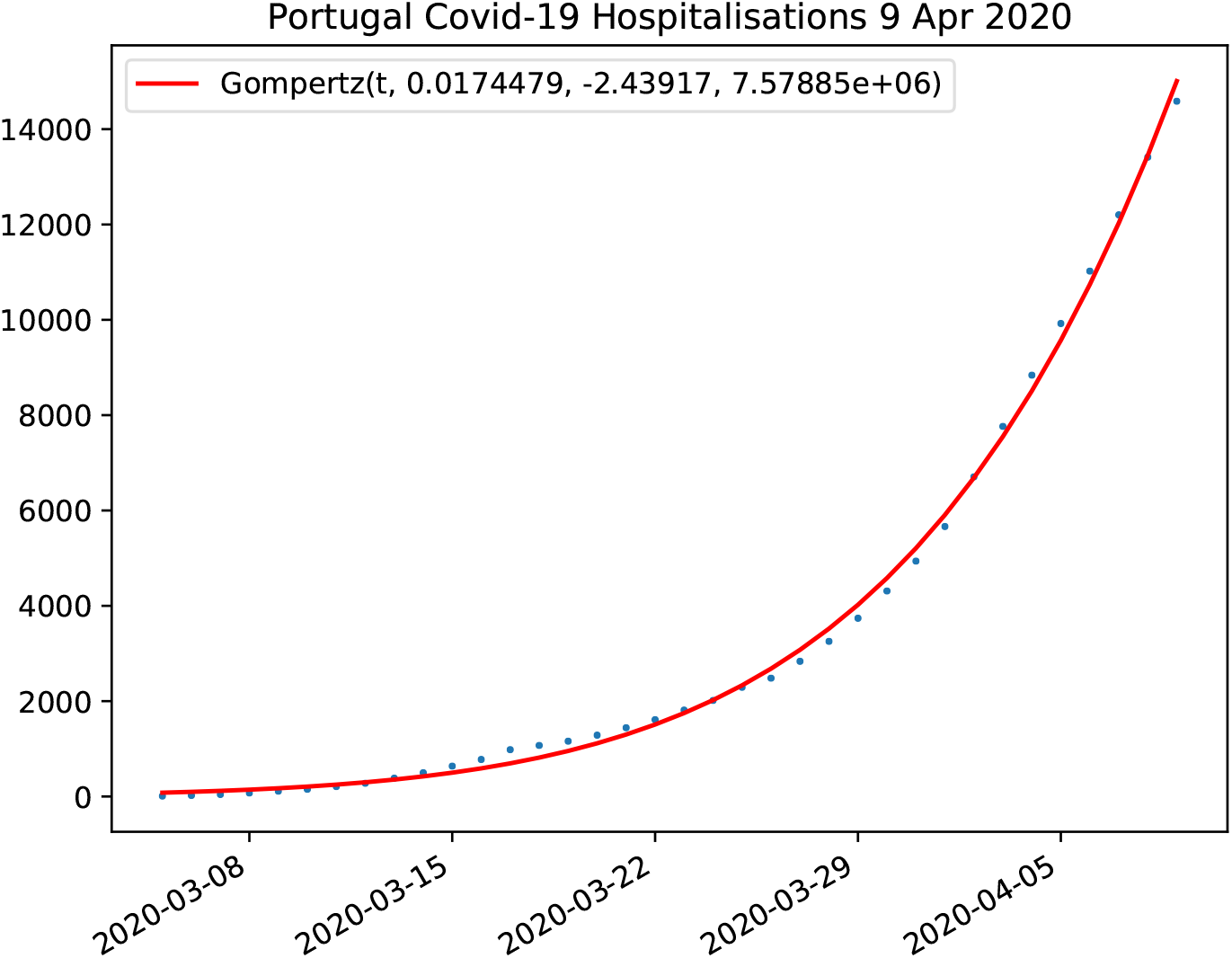
Gompertz Function Growth in Portuguese Covid-19 Hospitalisations.

There is also a very big difference in the loss of predictive power out of sample. Both fits produced over-estimates of future hospitalisations. On 9 April 14, 600 people had been hospitalised for Covid-19 in Portugal. By 15 May a total of just over 50, 000 hospitalisations were recorded. The exponential fit would have reached 1.3 million and the Gompertz Function fit 274, 000 if they were projected out to 15 May.

But it’s not the long term predictions that matter on 9 April. We’ve already seen that by 16 April the Gompertz Function fit (Figure 46) had absolute errors below 10% for more than a week and below 15% for three weeks.The important question is how useful the predictions made on 9 April would have been in the short term. Figure 96 shows the out of sample errors for the two fits.

**Figure 96:**
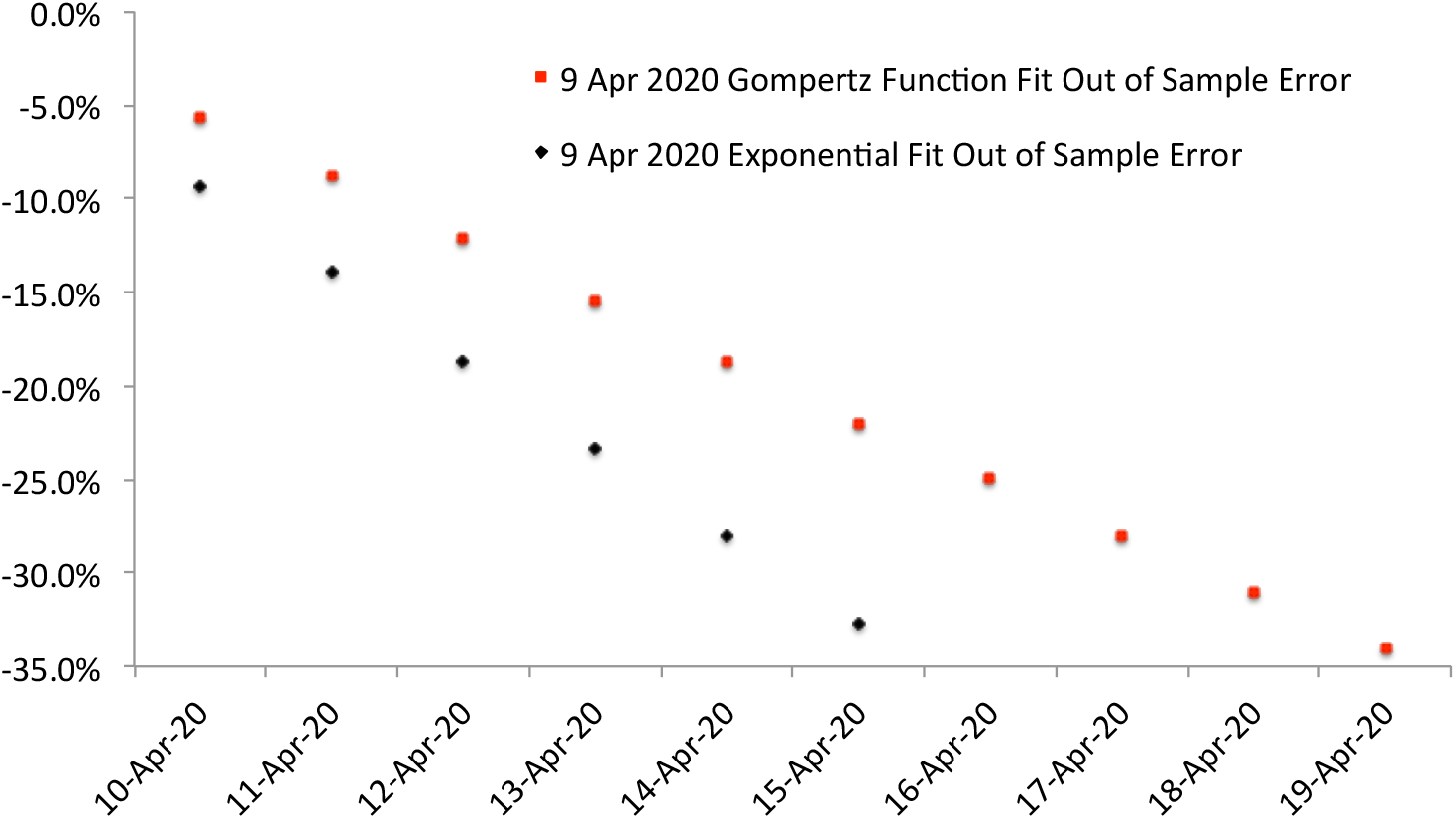
Out of sample errors Gompertz Function and Exponential fits.

Remember that hospitalisation data is typically delayed by two days, so that while the first out of sample day is 10 April, the calculations could not have been made until 11 April. Only on 12 April is the out of sample period in the future of the calculation. By this time the error in the exponential growth fit is already almost -18%, compared with -12% for the Gompertz Function. By 16 April the exponential fit error is -38% compared to -25% for the Gompertz Function.

We have seen that out of sample error levels for Gompertz Function fits drop rapidly to 10% or less for extended periods as the epidemic progresses. But in the early stages errors over a period of a week or more will generally exceed this level. This is because there are two distinct phases of Gompertz function growth.

### 5.2 Two Phases of Gompertz Function Growth

We have shown evidence for the Extended Gompertz Function Model of alternating periods of Gompertz Function Growth and linear growth. Within the former we generally observe two phases. While there are excellent Gompertz Function fits all through an epidemic, the Gompertz Function that fits the data for, say, the first 3 weeks is different from the one that fits them 3 weeks later. We illustrate this again with Portuguese Hospitalisations from the initial Covid-19 epidemic. Figure 33 shows how well the Gompertz Function fits the hospitalisation data from 5 Mar to 15 May 2020. But we can also see from Figure 97 that the Gompertz Function from 9 April is a much better fit to the data from 5 March to 9 April. The mean squared error over this part of the sample is almost four times as large for the 15 May fit as it is for the one on 9 April. This behaviour is generic. It is the reason that there are still large in-sample errors in the initial weeks for fits whose out of sample errors fall dramatically as in Figures 42 to 48.

**Figure 97:**
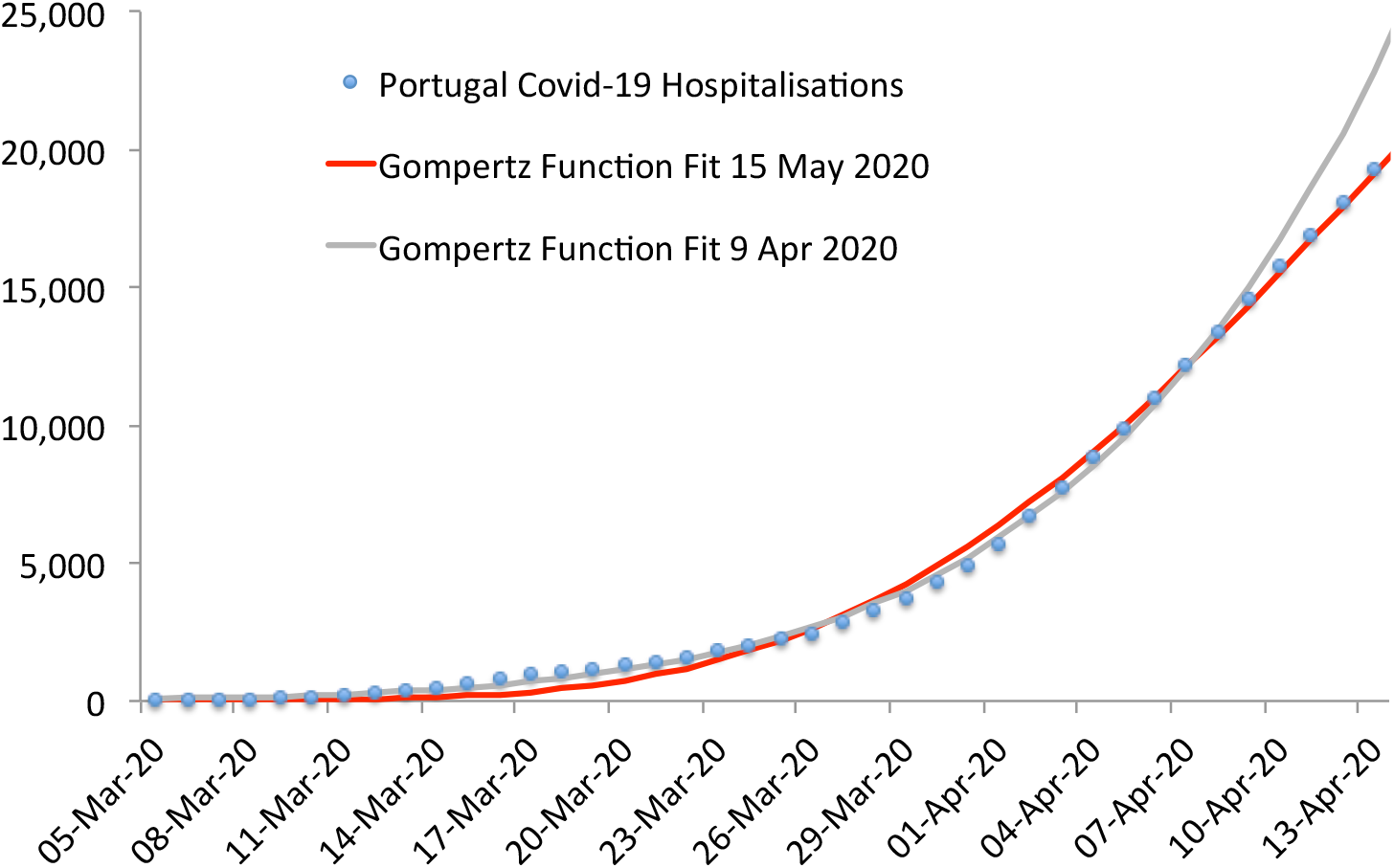
Gompertz Function Fits from 9 Apr and 15 May shown from 5 Mar to 9 Apr.

Note that this is inconsistent with the idea that early epidemic observations can be viewed as being a ‘noisy’ version of the values from the final Gompertz Function. The noise would have to be systematically biased to be much larger in the initial weeks of the epidemic than it is in the later stage. There is certainly no reason to imagine that ‘measurement error’ in the count of hospitalisations could behave in such a way.

We will return to the question of improving the predictive power of early stage fits and ways of using advanced statistical methods to produce accurate bounds on early stage surge levels in a separate publication.

The division between the ‘initial phase’ and the rapid convergence phase can be described in terms of the critical time *T*_1_ when the acceleration reaches its peak. Each successive fit produces a different pair (*a*_*i*_, *b*_*i*_) and hence a different value for 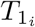. During the initial phase 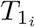 will be in the future of the calculation date *D*_*i*_. The threshold for the end of the initial phase is the time from which all 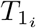 are in the past of date *D*_*i*_. In the case of the Portuguese Hospitalisations, 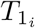 for the 15 May Gompertz Function fit is 29 March, the date it had converged to by 30 April. 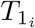 was consistently in the past of the calculation date by 16 April–which was effectively the end of the ‘initial phase’.

Of course we can’t know how long the initial phase will last. We just know that it will end. We know that the evolution will continue to follow the Gompertz Function Model or switch to linear growth–as happens every year in the initial influenza outbreak in Portugal and has happened in all of the Covid-19 outbreaks reported above.

It is often the case in a mathematical process that we know a certain event will occur if an iteration continues long enough–but we can’t know in advance how many iterations will be needed. In the meantime, we only know that we should continue the iteration.

We should continue to update the Gompertz Function Fits and monitor their out of sample predictive power. This can very easily be done every day.

### 5.3 Gompertz Function Fits in the Rapid Growth Initial Phase

During the initial phase it is common to find that the best fit has a very large value of the parameter *N*. In the Portuguese hospitalisation example on 9 April for example, *N* was over 7 million (which is 70% of the population of Portugal and not a believable long term prediction). The date of *T*_1_ given by the 9 April fit was 29 May. A week later the 16 April fit gave a *T*_1_ of 1 April–well in the past–and *N* had dropped below 82, 000.

It’s easy to see from the Gompertz Function dynamics why very large values of *N* will occur in the initial phase of very rapid growth. These dynamics are, as we’ve already discussed, completely determined by those of the Gumbel distribution. We can think of the parameters *a* and *b* obtained in a fit of data up to day *D* as defining a map from the time interval of days [0, *D*] to the real line by *x* = *ϕ*(*t*) = *at* + *b*. The number of positive derivatives of the Gompertz Function on [0, *D*] is the same as the number for the standard Gumbel distribution 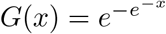 on the image interval *ϕ*([0, *D*]) = [*b, aD* + *b*].

Given any positive integer *k*, there’s a number *B*(*k*) < 0 such that whenever *x* < *B*(*k*) the first *k* derivatives of *G* are positive. When epidemic data is increasing very rapidly (say with growth like a polynomial of degree *k*), the nonlinear fit routine will find a Gompertz Function such that *b* + *aD* < *B*(*k*). The more negative the value of *B*(*k*), the larger the value of *N*.

In our example, on 9 April, *D* = 35 and the right hand endpoint of the image interval is *a*35 + *b* = − 1.8285. In the image interval the first four derivatives are positive and the first three are monotone increasing. Near the end of the interval the fourth derivative peaks and starts to decrease. This dampens the Gompertz Function fit’s growth relative to the exponential case, where derivatives of all orders are always positive.

The Gumbel distribution’s value at -1.8285 is approximately 0.00198. In order for the Gompertz Function value to approximate 14, 586, the number of cases on 9 April, we need to have a value of *N* such that 0.00198*N* ≈ 14, 586 or *N* ≈ 7.4 million.

### 5.4 Fitting to the Cumulative Rather than to Daily Data

In all of our examples we have shown fits to cumulative data. This is more directly aligned with our focus of predicting health care demands. The relevant issue is how many new patients will be admitted over some planning horizon rather than how many each day within that period.

But there is also a technical issue. We could have fit the daily data with the function *Y* (*t*) = *Ng*(*t, a, b*) where *g* is the probability density function of the Gumbel distribution. But the passage from daily data to the cumulative is equivalent to integration and has a significant smoothing effect. It is more efficient to first smooth the data then fit the curve and take first differences than it is to fit the daily data and then integrate.

The linear growth phases are also far more obvious in the cumulative than they are in the daily data.

Nevertheless, fitting the daily data to the derivative of the Gompertz function gives very similar results because of the very close approximation of 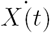 to the daily difference *X*(*t* + 1) − *X*(*t*). Note that this is a feature of the small values of *a* because a one-day grid on the interval [0, *D*] is equivalent to a grid of step size *a* on [*b, aD* + *b*].

## 6 Relation to Compartmental Models

Compartmental models such as the SEIR model where the linked dynamics of portions of a population that are susceptible (*S*) to an infectious disease, exposed to it but not yet able to transmit it (*E*), infectious (*I*) and removed (*R*) form the basis for much of the modelling of epidemics. The simpler SIR model (where exposure equates to infectiousness) was introduced by Kermack and McKendrick almost a century ago as an approximation (whose assumption of homogeneity in the population they acknowledged was unrealistic).

Like all mathematical models, these rely on a series of assumptions which are only approximately satisfied in reality. The hope is that they can still provide useful insight into the real world process.

One of the most restrictive assumptions is that the populations within the compartments are homogeneous. As Graham Medley, Chairman of the UK’s SPI-M pandemic modelling group, points out:^25^

> It is intriguing that, currently, most models of infectious disease transmission dynamics assume that all hosts are identical, when we know that they are not. For some infections, such as measles, it is probably adequate to consider that everybody is average when predicting the impact of immunisation. However, such models result in policy decisions that have assuming that “everybody is equal” as an unwritten assumption.For other infections, such as HIV, assuming that everybody is average is known to be inadequate; the commonest model structures assume that the population is divided into discrete groups, where everybody within the group is average for that group. But how should the groups be chosen, and how do they interact?

In the case of Covid-19, removing this simplifying assumption can produce outcomes which are greatly at odds with those produced by the standard models. In particular this can significantly lower the Herd Immunity Threshold, in either the SIR model [17] or in the SEIR model [8].

In [8], Gabriela Gomes et al show this with heterogeneity introduced through individual variation in either susceptibility or exposure in an SEIR model of deaths where the Herd Immunity Threshold is lowered substantially.

The cumulative simulated deaths for England reported in [8] are very well approximated by Gompertz functions. This may simply reflect the fact that they are constructed to be in reasonable agreement with the observed deaths data. It is also possible that heterogeneity could be ‘calibrated’ by targeting Gompertz Function growth in the outputs.

## 7 Using the Extended Gompertz Function Model in Future

### 7.1 Ongoing Covid-19 Outbreaks

While Covid-19 outbreaks continue we can expect that the alternating pattern of Gompertz Function growth and linear growth will be maintained. Indeed, this is what has been observed since the end of the 2020-2021 episodes.

In England, for example, by the middle of March 2021, the Gompertz Function growth in hospitalisations had ended. Linear growth that began then continued until the end of May.

Then another Gompertz Function growth period began, this one ‘out of season’.This caused the Government to delay the planned ‘reopening’ for several weeks at tremendous economic and social cost. But by mid July the hospitalisation growth was linear once again.

In the autumn there was another seasonal Gompertz Function growth phase which, as in 2020 was replaced by linear growth in November. It seems likely that another period of Gompertz Function growth should have been expected in December. It appears to have begun in the first week of December with the Omicron variant.

We cannot know in advance when a transition from linear growth to Gompertz Function growth will occur. We should be ready for these at the usual respiratory virus intervals, but it is now clear that Covid-19 can appear ‘out of season’ as well.

To benefit most from the use of this model, it is essential to be able to detect the transitions, especially from linear to Gompertz Function Growth.

While we know that the latter will fairly rapidly become highly predictable, we must also deal with the initial period where the predictive power is much lower–and we cannot know in advance how long that phase will persist.

There are many ways of approaching these problems. Ours is to use a probabilistic approach to estimate, based on recent observed daily admissions what future peaks will look like on average. This has proved very effective both in detecting the onset of the Gompertz Function growth phase and in providing useful bounds for growth of admissions until the predictive power of the Gompertz Function fits becomes sufficient. We will discuss this approach in a separate publication.

### 7.2 Preparing for the Return of Seasonal Influenza

We have shown that the Extended Gompertz Function Model describes the Influenza cycle in Portugal and have shown indirect evidence that this is also the case elsewhere. It is important to test this conjecture in as many other countries as possible. If it is confirmed, the model can be an important addition to influenza surveillance systems and alleviate the frequent crises that occur when the main seasonal outbreaks are larger than usual.

### 7.3 Pandemic Preparedness

Finally, the Extended Gompertz Function Model has immediate applications to preparation for future pandemics. When such an event occurs we should expect cases, hospitalisations and so on to follow Gompertz Function Growth. During these periods the Gompertz Function Model has excellent predictive power.

We should expect waves to be separated by periods of linear growth indicating endemic status for the initial virus and the likelihood of a variant driving a subsequent wave.

It is likely that the next epidemic (of any sort) will also begin with Gompertz Function growth. We are hopeful that,when that happens, rather than panicked visions of exponential growth, this model will be applied to make accurate predictions.

## A The Geometry Behind the Gompertz Function

This appendix provides a quick sketch of the geometry behind the Gumbel distribution and the Gompertz Function. An introduction to this topic is contained in [4] where we described a new solution to the problem of characterising the domains of attraction of the Gumbel distribution and the other Extreme Value Distributions.

The ‘location-scale’ parameters in univariate probability distributions arise from the action of the affine group *t* →*at* + *b*. As Elie Cartan taught us [2], group actions produce geometry and geometry produces differential invariants such as curvature. The Gumbel distribution is part of an exceptional maximally symmetric constant ‘curvature’ family, the Extreme Value Theory attractors and is the one with zero curvature.

Cartan’s methods [19] show that for a smooth univariate probability distribution *F* the equivalence class of *F* under the affine group is determined by the functional relation between the invariant 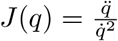 and *q* = *log*(*F*).

If we denote that function by *h* so that the relation is *J* (*q*) = *h*(*q*), then we may also view the equivalence class as determined by the second order ordinary

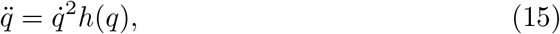

and identify the location scale parameters of *F* as the initial conditions for this differential equation.

For each *F* this equation is the Euler-Lagrange equation (equivalently Hamilton’s equations) for the variational problem of minimising the integral of 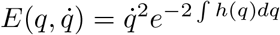.

*E* is the conserved quantity that Noether’s theorem associates to the time translation symmetry subgroup of the affine group so we can think of it as an ‘energy’.

The 1-parameter family of distributions with 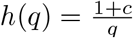 corresponds to the Extreme Value distributions. The Gumbel distribution is given by *c* = 0, the values *c* < 0 give the Weibull distributions and *c* > 0 the Fréchet distributions. If we parametrise these with 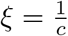 then as |*c* | → ∞ both the Weibull and Fréchet families tend to the Gumbel distribution.

Only for the Extreme Value distributions is there a second symmetry of the differential equation. It is obtained by simultaneously scaling *t* and *q* by the same factor. (This is not a Noether symmetry so there is no associated conservation law.)

This maximally symmetric case is diffeomorphic to the affine group itself, identifying the differential system for equation (15) with the Maurer Cartan structure equations for the affine group.[3]

A distribution *F* on the interval [*A, B*] (where either of *A* or *B* may be infinite) is in the domain of attraction of an Extreme Value distribution corresponding to the constant *c* if and only if its invariant *h*_*F*_ tends to 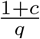 as *t*→*B*.

The Gumbel distribution’s domain of attraction is the *thin-tailed distributions*– the distributions defined on intervals of the form [*A*, ∞] which have finite moments of all orders (such as the normal, Laplace or logistic distributions). So if *F* is any thin-tailed distribution the defining equation for the equivalence class of *F* must approach that of the Gumbel distribution as the percentile level tends to 100%.

This means that any probability distribution *F* in the Gumbel domain of attraction will have a good fit by a Gumbel distribution beyond some quantile *Q* and hence a good fit by the Gompertz Function to a model *Y* = *NF* (*t, a, b*) above the value of *t* corresponding to *Q*.

In a preliminary investigation we have seen that the quality of any fit to epidemic data seems to indicate that, for practical purposes, *Q* ≈0.5. So the portion of the epidemic curve that occurs after about half of the susceptible population *N* is affected will be essentially indistinguishable from a Gompertz Function. If we add to this the empirical observation that the daily peak comes at approximately 0.37*N*, the epidemic curve is bound to resemble a Gompertz function very closely over its entirety.

The long evolutionary path to a stable equilibrium between virus and host seems to have led to the emergence of the Gompertz function as a stable equilibrium for epidemic evolution.

The Gumbel energy is 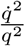, the square of the *relative* velocity so it would be interesting to see if there are known biological processes that minimise this.The ‘simpler’ Lagrangian with energy 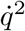 yields an exponential distribution (the *c* = −1 case of the Weibull family) for *F*. This could also be a stable equilibrium but presumably one with no people and no human viruses.

There is an intriguing bit of evidence for the idea that viruses themselves are following a Gompertz Function in an example of observations of bacteriophage viruses (A. G. McKendrick 1939). McKendrick’s model produces a good fit but a Gompertz density fits the data with only about one tenth of the mean squared error of the McKendrick model.

## B Covid-19 Data Sources

The Covid-19 data presented in our analysis were taken from:

**Sweden**: https://www.socialstyrelsen.se

**London**: https://coronavirus.data.gov.uk

**Isle de France**: https://www.data.gouv.fr

**Ontario**: https://covid-19.ontario.ca

**Portugal**: https://github.com/dssg-pt/covid19pt-data

**Amazonas and Rio de Janeiro**: https://datasus.saude.gov.br/

## Data Availability

The Covid-19 data presented in our analysis were taken from: Sweden: https://www.socialstyrelsen.se
London: https://coronavirus.data.gov.uk
Isle de France: https://www.data.gouv.fr
Ontario: https://covid-19.ontario.ca
Portugal: https://github.com/dssg-pt/covid19pt-data
Amazonas and Rio de Janeiro: https://datasus.saude.gov.br/

https://coronavirus.data.gov.uk

https://www.data.gouv.fr

https://covid-19.ontario.ca

https://github.com/dssg-pt/covid19pt-data

https://datasus.saude.gov.br/

## Footnotes

1 The rise in hospitalisations which prompted the UK Government to delay its ‘re-opening’ measures in England in June 2021 began when the linear growth that followed the winter 2020-2021 outbreak was succeeded by Gompertz Function growth from 29 May to 14 July. Linear growth in hospitalisations resumed on 15 July and continued until the end of September.

2 As Graham Medley, chairman of the UK’s SPI-M modelling group, pointed out [16] providing scenarios is the (often misunderstood) purpose of the Covid-19 modelling presented to the UK’s SAGE committee. Unfortunately, not all modellers are as clear about this as Medley.

3 The parameters are determined by the standard statistical process of non-linear regression which finds the Gompertz Function of ‘best fit’ to the data.

4 On day *n* we plot the average for days *n* −3 … *n* + 3 to remove the lag.

5 While it’s possible to fit an exponential to early outbreak data, the fit rapidly loses any predictive power. Exponential growth is simply not sustainable. For example, if the exponential growth that best fit the death data in Italy on 21 March 2020 was continued until 13 May, the death toll would have exceeded the population of the country. By contrast, Gompertz Function fits to this early data have much better predictive power out of sample. See Section 5.1

6 Although it is less visually obvious in the cumulative deaths graph than in the daily one, the same asymmetry is there–reflected in the fact that the concavity changes at the time when *X*(*t*) ≈ 0.37*N* rather than 0.5*N* as it would in the symmetric case.

7 We are grateful to Prof. B.A.Shadwick for the Python program which we have used for all of the fits produced in this paper.

8 Fits are done using with time in days starting from day 0.

9 The last severe ‘flu season’ in the Northern Hemisphere

10 This corresponds to putting a band around the prediction rather than the data.

11 For example,we recently found an example where the number of cases on a given day in the first Quarter of 2020 was revised in the third Quarter of 2021.

12 Alternatively we can fit 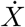 directly by non-linear regression with very similar results.

13 For example, a common ‘S-curve’ from population dynamics is the Logistic function *X*(*t*) = *NL*(*t*) where *L* is the Logistic distribution *L*(*t, a, b*) = (1 + *exp*(−(*at* + *b*)))^−1^.

14 As we explain in the Appendix, the Gumbel distribution is not just another ‘S-curve’ but is exceptional from the point of view of the geometry of the affine group on the line (the ‘Location-Scale’ transformations). This geometry is what explains the Gumbel distribution’s role as an ‘attractor’ in Extreme Value Theory and contributes to its apparent inevitability in epidemic behaviour.

15 The Gompertz function can fit growth with arbitrarily many positive derivatives. See Section 5 for a brief discussion of the ‘exponential growth phase’.

16 This underestimation by the Logistic function was observed in Ebola and Zika outbreaks. See for example [18]

17 The Gompertz Function approaches the ‘final value’ *N* asymptotically as time tends to ∞.

18 We are using the proportion of the population that needs to be immune in order for new infections to decline as our definition of the Herd Immunity Threshold.[14].

19 This approximation has excellent predictive power. If the linear fit to the week of data from 21-27 April 2020 is extrapolated forward for two weeks, its average absolute percentage error out of sample is only 0.8% and the maximum absolute percentage error is 2.7%.

20 https://transparencia.sns.gov.pt/explore/dataset/atendimentos-nos-csp-gripe/export/?disjunctive.ars&sort=dia)

21 This includes the 2020-2021 season, which differs from the rest only by a very large reduction in scale.

22 The data from the 2020-2021 season, which also follows Gompertz Function growth as we show below, is invisible on the scale of this graph.

23 We are grateful to D. Earn for providing this data.

24 For example in an SIR model, if we assume that the entire population is susceptible at the outset, so 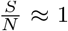, then the while that condition holds, *I* grows exponentially. But the SIR equations also show the evolution of *S* will rapidly violate the condition 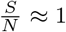.

25 https://www.lshtm.ac.uk/aboutus/people/medley.graham

